# A law of human glucose homeostasis for inferring glucose regulatory capacity from glucose time series

**DOI:** 10.1101/2025.09.30.25337012

**Authors:** Hikaru Sugimoto, Ken-ichi Hironaka, Tomoko Yamada, Kazuhiko Sakaguchi, Wataru Ogawa, Shinya Kuroda

## Abstract

Diabetes affects over 500 million people, underscoring the need for accurate and accessible screening tools. Common measures, including fasting glucose and continuous glucose monitoring (CGM)-derived indices, cannot directly quantify the body’s capacity to regulate glucose, partly because an equation linking glucose intake and observable glycemic trajectories to underlying regulatory capacity has not been established. Here, we derive an equation *F* **= *K***·***G***, connecting glucose input (*F*) and trajectory (***G***) to a regulatory-capacity vector (***K***) integrating insulin secretion, insulin sensitivity, and insulin-independent glucose disposal. ***K*** decomposes into proportional–integral–derivative (PID)-like components from control theory, while ***G*** integrates glucose-trajectory characteristics, including area under the curve, amplitude, and temporal distortion. Using clamp data and mathematical models of glucose-insulin-C-peptide interactions, we derive and evaluate the equation. Across oral glucose tolerance tests and CGM data from over 2,000 individuals, ***K*** can be inferred without direct measurement of insulin and stratifies diabetes-related phenotypes independently of standard clinical covariates. This framework further suggests four exploratory patterns of impaired glucose regulation, including one characterized by deficient insulin-independent glucose lowering. These results provide a mechanistic interpretation of how biological systems achieve robust glucose regulation and enable physiologically interpretable digital phenotyping from routinely acquired glucose data.

## Introduction

Glucose homeostasis is a fundamental component of human physiology, and its dysregulation underlies diabetes mellitus, which now affects over 500 million people worldwide^1,2^. Current indices of glycemic control, such as single-time-point blood glucose measurements and glycated hemoglobin (HbA1c), provide limited temporal resolution and little mechanistic insight into the physiological processes that maintain glucose homeostasis^3–9^. These limitations have prompted efforts to establish theoretical frameworks that better explain glucose regulation.

Over the past few decades, numerous mathematical models^10–15^ and machine learning approaches^16–19^ have been proposed to advance our understanding of glucose homeostasis. One example is the hyperbolic law^20–22^, which states that as insulin sensitivity declines, insulin secretion increases to maintain glucose homeostasis, resulting in a compensatory shift along the hyperbolic curve defined by these two variables. Consequently, the disposition index (DI), defined as the product of insulin sensitivity and insulin secretion, has been widely used as an indicator of glucose regulatory capacity^20^. However, this framework relies on invasive and labor-intensive testing, such as oral glucose tolerance tests (OGTTs) or glucose clamp procedures, and does not provide a complete picture of glucose trajectories. While machine learning models are increasingly applied to predict diabetes-related outcomes, they are often constrained by complexity, lack of interpretability, and limited methodological transparency^23^. These limitations reinforce the enduring value of simple quantitative equations that provide mechanistically interpretable and robust insights.

Here, we sought to develop a theoretical framework for human glucose homeostasis that is both physiologically interpretable and practically applicable. We first developed a reduced-order model of glucose, insulin, and C-peptide dynamics that retains key components represented in established physiological models, including glucose effectiveness, insulin sensitivity, insulin secretion, and insulin clearance. Despite its simplified structure, the model reproduced glucose– insulin dynamics across hyperglycemic and hyperinsulinemic–euglycemic clamps, OGTTs, mixed-meal tolerance tests (MMTTs), and intravenous glucose tolerance tests (IVGTTs). We then analytically integrated the model to derive a compact equation, *F* **= *K***·***G***, which relates time-integrated glucose input (*F*) and measurable features of the glucose trajectory (***G***) to a regulatory-capacity vector (***K***). The vector ***K*** generalizes the DI and aligns conceptually with proportional–integral–derivative (PID)-like control systems used in control engineering to achieve robust regulation^24^. As continuous glucose monitoring (CGM) and dietary records now enable glucose trajectories and meal-derived inputs to be measured minimally invasively, this framework can be used to infer latent glucose-regulatory capacity (***K***) from glucose (***G***) and input (*F*) data alone. Furthermore, we demonstrate that the inferred regulatory-capacity components stratify glucose tolerance and diabetes-related phenotypes independently of several clinical covariates.

## RESULTS

### Mathematical modeling of human glucose homeostasis

Our aim was not to construct a maximally detailed physiological simulator, but rather to identify a minimal dynamical structure that satisfies three requirements. First, the model should reproduce glucose, insulin, and C-peptide dynamics during glucose challenges, including clamp tests, OGTTs, MMTTs, and IVGTTs. Second, its components should remain interpretable in terms of established glucose–insulin physiology, including glucose effectiveness, insulin sensitivity, insulin secretion, insulin clearance, and C-peptide kinetics. Third, the model should be analytically reducible to a simple equation that relates glucose input, glucose waveform features, and regulatory capacity. Regulatory capacity is difficult to quantify without insulin, C-peptide, or clamp data, and this reduction allows it to be inferred from the more readily measurable glucose input and glucose trajectories. Although existing models have proven valuable for simulating glucose–insulin dynamics or estimating physiological parameters, a unified framework that is simultaneously reduced-order, physiologically interpretable, and analytically tractable for estimating regulatory capacity from glucose data alone has not yet been established.

We first analyzed hyperglycemic and hyperinsulinemic–euglycemic clamp data from 121 individuals in the *Kobe University Study*^25^, including 50 with normal glucose tolerance (NGT), 18 with impaired glucose tolerance (IGT), and 53 with type 2 diabetes mellitus (T2DM) (**Fig. 1a**). Clamp tests, which measure blood glucose, insulin, and C-peptide concentrations during controlled infusions of glucose and insulin, have been the gold standard for assessing glycemic control abilities^10^. Building on previous studies (**Supplementary Notes 1, 2, Extended Data Fig. 1a–o**), we formulated a compartment model capturing the interactions among six compartments: plasma glucose (*G*), remote glucose (*Q*), remote insulin (*X*), serum insulin (*I*), serum C-peptide (*C*), and remote C-peptide (*Y*) (**Fig. 1b, c**). The model was described by the following equations:

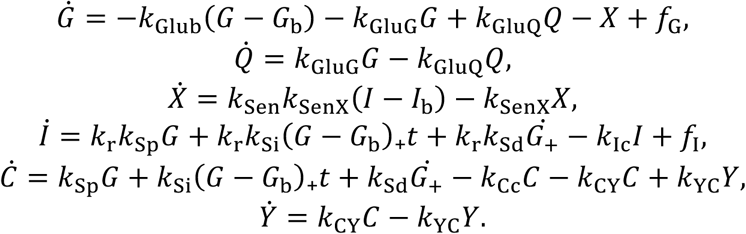

**Fig. 1.**
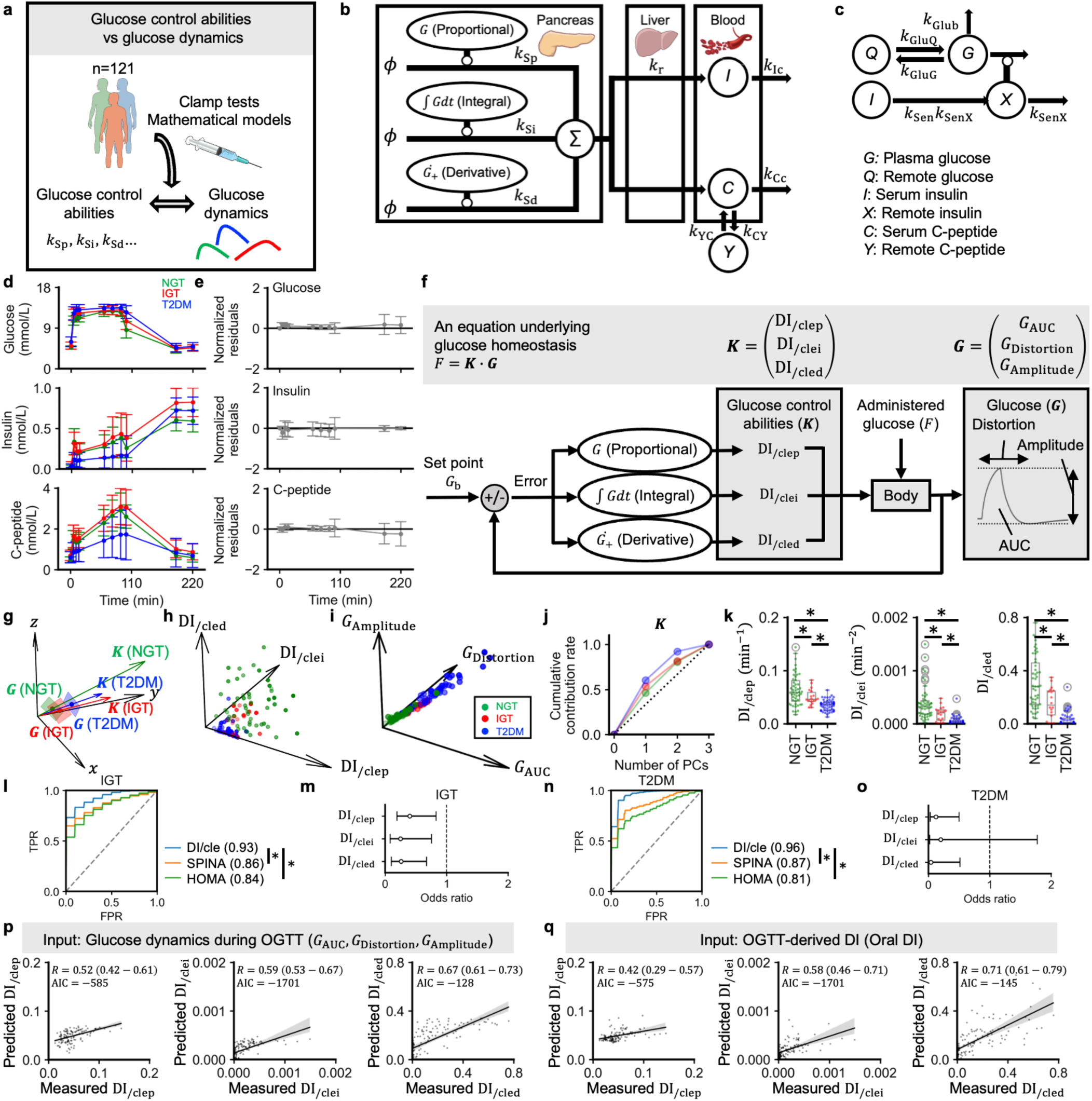
Inference of glucose control abilities from clamp data and their link to glucose dynamics. **a,** Study design. Glucose control abilities and glucose dynamics were quantified in 121 individuals from the *Kobe University Study* by combining clamp tests with mathematical modeling. **b, c,** Model diagrams used to infer glucose control abilities. **b,** the insulin-C-peptide regulatory model and **c,** the glucose regulatory model. Circular nodes denote variables, arrows denote fluxes, and lines ending in circles represent regulatory activation. k with subscripts denote rate constants. φ denotes a fixed value. **d,** Time courses of plasma glucose, serum insulin, and serum C-peptide during the sequential clamp protocol (0–220 min), consisting of a hyperglycemic clamp (0–90 min), a transition period (90–100 min), and a hyperinsulinemic– euglycemic clamp (100–220 min). Data are mean ± SD, stratified by glucose tolerance status (NGT, normal glucose tolerance, green; IGT, impaired glucose tolerance, red; T2DM, type 2 diabetes mellitus, blue). **e,** Time courses of the normalized residuals [(simulated − measured)/measured values] for glucose, insulin, and C-peptide. Data are mean ± SD. **f,** Schematic linking glucose control abilities and glucose dynamics features. The administered glucose *F* satisfies *F* = ***K*** · ***G***, where ***K*** = (DI_/clep_, DI_/clei_, DI_/cled_) comprises the proportional, integral-like, and derivative disposition indices and ***G*** = (*G*_AUC_, *G*_Distortion_, *G*_Amplitude_) summarizes the glucose response. **g,** Geometric representation of the *F* = ***K*** · ***G*** equation across glucose tolerance groups. For each group, the vector ***K*** (from the origin) and the plane {***G***: ***K*** · ***G*** = *F*} are shown. Axes correspond to *G*_AUC_, *G*_Distortion_, and *G*_Amplitude_ for ***G*** and to DI_/clep_, DI_/clei_, and DI_/cled_ for ***K***. As glucose tolerance worsens from NGT to IGT to T2DM, |***K***| decreases and the corresponding ***G*** plane recedes from the origin. **h, i,** Scatter plots of individual ***K*** (**h**) and ***G*** (**i**) values. **j,** Cumulative proportion of variance explained as a function of the number of principal components in a principal-component analysis of ***K***. **k,** Box plots of DI_/clep_, DI_/clei_, and DI_/cled_ in NGT, IGT, and T2DM groups. \**Q* < 0.05; Welch’s *t*-test with Benjamini–Hochberg correction. **l,** Receiver operating characteristic (ROC) curves for classifying glucose intolerance (IGT or T2DM versus NGT) from multivariable logistic-regression models built on the DI_/cle_ set (DI_/clep_, DI_/clei_, and DI_/cled_), the SPINA set (SPINA-GBeta, SPINA-GR, and SPINA-DI), or the HOMA/QUICKI set (HOMA-β, HOMA-IR, and QUICKI). AUC values are shown in parentheses. \**P* < 0.05. **m,** Odds ratios for glucose intolerance estimated from a multivariable logistic regression with standardized DI_/clep_, DI_/clei_, and DI_/cled_ as predictors. Points are odds ratios; horizontal bars indicate 95% confidence intervals. **n,** ROC curves for classifying T2DM versus non-T2DM using logistic regression models based on DI_/cle_, SPINA, or HOMA/QUICKI feature sets. AUC values are shown in parentheses. \**P* < 0.05. **o,** Odds ratios for T2DM estimated from a multivariable logistic regression with standardized DI_/clep_, DI_/clei_, and DI_/cled_ as simultaneous predictors. **p,** Predicted versus measured values of each ***K*** component (DI_/clep_, DI_/clei_, and DI_/cled_), from separate linear regressions using the ***G*** components (*G*_AUC_, *G*_Distortion_, and *G*_Amplitude_) obtained during an oral glucose tolerance test (OGTT) as predictors. “Measured” indices are values computed directly from clamp data, while “predicted” values are estimates obtained by regression. **q,** As in (p) but using the OGTT-derived oral disposition index (oral DI) as the predictor. Shaded areas denote 95% confidence intervals. *R*: Spearman’s correlation coefficient; AIC: Akaike information criterion. The AIC is reported so that models with different numbers of predictors can be compared based on penalties.

Here, *k*_Glub_ and *k*_GluG_⁄*k*_GluQ_ represent distinct aspects of glucose effectiveness. The parameter *k*_Glub_ captures an irreversible process in which glucose exits the bloodstream to extravascular compartments under hyperglycemia. By contrast, *k*_GluG_ and *k*_GluQ_ govern the reversible exchange of glucose between the plasma (*G*) and the remote compartment (*Q*): glucose flows from *G* to *Q* when *G* rises, and it returns from *Q* to *G* when *G* falls. Larger values of *k*_Glub_ and *k*_GluG_⁄*k*_GluQ_ indicate stronger *G*-lowering effects in response to hyperglycemia. The parameter *k*_Sen_ quantifies insulin sensitivity. The parameters *k*_Sp_, *k*_Si_, and *k*_Sd_ represent the basal, potentiated, and first-phase components of insulin secretion, respectively, conceptually corresponding to the proportional (P), integral-like (I), and derivative-like (D) elements of a PID controller, which is widely used in control systems engineering^26,27^. The parameters 1 − *k*_r_, *k*_Ic_, and *k*_Cc_ denote hepatic insulin clearance, peripheral insulin clearance, and C-peptide clearance, respectively. External infusion rates of glucose and insulin are denoted by *f*_G_ and *f*_I_, respectively (**Extended Data Fig. 1p**). *Ġ*_+_equals *Ġ* when *G* is increasing and zero otherwise. *G*_b_ denotes the fasting blood glucose concentration.

Although the model is formulated as a first-order approximation around the basal glucose– insulin equilibrium, this does not imply that the underlying glucose–insulin system is linear. Rather, our objective was to determine whether such a first-order approximation is sufficient to capture glucose dynamics over the experimentally relevant range. To this end, we evaluated the model under hyperglycemic and hyperinsulinemic–euglycemic clamp conditions, which impose large perturbations in glucose and insulin concentrations. This strategy parallels prior work showing that complex nonlinear processes, such as gene–gene interactions, can often be approximated effectively with linear models around equilibrium points^28^. Normalized residuals [(simulated − measured)/measured values] for the glucose, insulin, and C-peptide trajectories were close to zero, even across the upper physiological ranges of glucose and insulin imposed by the clamps (**Fig. 1d, e**). Correlations between simulated and observed data were 0.96 (95% CI, 0.95–0.96) for glucose, 0.97 (95% CI, 0.96–0.97) for insulin, and 0.95 (95% CI, 0.94–0.97) for C-peptide. Moreover, the insulin sensitivity index, derived from a nonlinear model^10,25^, was significantly correlated with *k*_Sen_ (R = 0.92, 95% CI: 0.89–0.95). These results indicate that this simple formulation remains valid across a wide physiological range, even in the presence of underlying nonlinearities.

### Reproducibility and generalizability of the mathematical model

We next asked whether the model remained accurate when glucose dynamics were measured at higher temporal resolution. We fitted the model to *Kobe University Study* clamp data in which plasma glucose was sampled at shorter intervals during the clamp protocol. The simulated glucose trajectories closely tracked the measured time courses across NGT, IGT, and T2DM participants, and normalized residuals, defined as (simulated−measured)/measured, were centered near zero throughout the experiment (**Extended Data Fig. 1q**). Across all participants and sampling times, simulated and observed glucose concentrations were highly correlated (R = 0.98; 95% CI, 0.98–0.99). In this model, the insulin-dependent glucose-lowering action is represented by a linear term, *X*. To test whether this formulation was appropriate, we compared it with nonlinear alternatives in which the action was modeled as *GX* or (*G* − *G*_b_)*X*. The linear *X* model performed at least as well as the *GX* model and yielded a significantly stronger positive correlation between simulated and observed glucose than the (*G* − *G*_b_)*X* model (**Extended Data Fig. 1r**), supporting the use of the linear form.

We then evaluated reproducibility in an independent clamp dataset comprising 64 participants from the *Kobe University Study2*^6^. After fitting participant-specific parameters, the model reproduced the measured time courses of plasma glucose, serum insulin, and serum C-peptide during both the hyperglycemic and hyperinsulinemic–euglycemic clamp phases (**Extended Data Fig. 1s–u**). Simulated and observed concentrations were strongly correlated for glucose (R = 0.97; 95% CI, 0.97–0.98), insulin (R = 0.98; 95% CI, 0.97–0.98), and C-peptide (R = 0.98; 95% CI, 0.98–0.99).

To further assess generalizability, we evaluated the model across independent datasets and reference-model simulations, including oral glucose tolerance tests, mixed-meal tolerance tests, intravenous glucose tolerance tests, and *in silico* physiological-model benchmarks (**Supplementary Notes 3-11, Extended Data Figs. 2 and 3**). Across these settings, the model reproduced glucose, insulin, and C-peptide dynamics despite differences in glucose dose, route of administration, nutritional context, species, and reference-model structure. Collectively, these results support the robustness of the model for capturing glucose–insulin–C-peptide dynamics across glucose tolerance states, sampling resolutions, and independent experimental and simulated datasets.

### A governing equation of human glucose homeostasis

Building on this model, we next sought to derive a unifying equation linking observable glucose trajectories to regulatory capacity. Previous studies have indicated that features of glucose dynamics, such as mean, variance, and temporal autocorrelation, are independently associated with glycemic control and diabetes complications^5–9^. Recent machine-learning analyses have also suggested that entire postprandial glucose trajectories can be reconstructed using these three statistics^9^. However, these links are merely correlative and lack a theoretical basis. We therefore aimed to formulate an equation that connects these three measurable features of glucose dynamics to the system’s underlying regulatory capacity.

By integrating the differential equations between two states in which plasma glucose and the insulin-related state variables have returned to their basal values and all state derivatives are zero (*e.g.,* a fasting condition in which a glucose bolus is administered over the first few minutes and, after sufficient time has elapsed, plasma glucose returns to its basal value and the system reaches steady state), we derived a single equation (**Fig. 1f, Supplementary Note 12**):

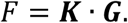

Here, *F* denotes the integrated glucose input over the analysis window. The ***K*** represents a composite regulatory capacity vector and is defined as: ***K*** = :*k*_Glub_ + *k*_Sen_*k*_r_*k*_Sp_⁄*k*_Ic_, *k*_Sen_*k*_r_*k*_Si_⁄*k*_Ic_, *k*_Sen_*k*_r_*k*_Sd_⁄*k*_Ic_;. The first component represents the glucose-lowering effect proportional to the current blood glucose level and is termed DI_/clep_. The second component captures glucose-lowering effects dependent on the integrated difference between current and basal glucose levels over time and is termed DI_/clei_. The third component reflects glucose-lowering effects related to the derivative of blood glucose and is termed DI_/cled_. Conceptually, these components align with the P, I-like, and D-like terms of a PID control system. The ***G*** was defined from plasma glucose dynamics: ***G*** = (∫(*G* − *G*_b_)*dt*, ∫(*G*− *G*_b_)h*tdt*, *G*_max_ − *G*_min_). These components reflect the area under the curve (*G*_AUC_), temporal distortion (*G*_Distortion_), and amplitude (*G*_Amplitude_), respectively. Conceptually, these components align with previously reported features, such as mean, temporal autocorrelation, and variance, that have been independently associated with the DI and diabetes complications^5,6,9^.

Simulation analyses showed that *G*_AUC_ alone was insufficient to infer either total glucose input (*F*) or glucose-regulatory capacity (***K***), because waveforms with similar *G*_AUC_ could differ in *G*_Amplitude_ and *G*_Distortion_ (**Supplementary Note 13, Extended Data Fig. 4a-n**). By contrast, the three-dimensional formulation *F* = ***K*** · ***G*** more accurately recovered administered glucose input and distinguished differences in regulatory capacity from glucose waveform structure. Thus, we modeled glucose homeostasis using the multidimensional glucose-dynamics vector (***G***) rather than a one-dimensional AUC-based representation.

### Geometric representation of *F* = *K* · *G* reveals inter-individual heterogeneity in *K*

We then applied the *F* = ***K*** · ***G*** framework to visualize regulatory capacity (***K***), thereby extending the classical DI into a multidimensional space. The classical DI is typically visualized as a hyperbolic curve defined by the trade-off between insulin sensitivity and secretion^20,29^. Even among individuals with NGT, considerable inter-individual variability exists along this curve^20^. To extend this concept, we developed a geometric representation of *F* = ***K*** · ***G*** (**Fig. 1g-i**). For fixed *F* and ***K***, ***G*** is constrained to a plane defined by *F* = ***K*** · ***G*** (**Fig. 1g**). As glucose tolerance declines, ***G*** is expected to increase, while ***K*** is expected to decrease. We estimated ***K*** from clamp tests and ***G*** from OGTT data (**Fig. 1h, i**). Although ***G*** is ideally calculated using glucose profiles between steady states (approximately 0-180 min during the OGTT), we observed a strong correlation between estimates derived from the 0-120 min window and the extended interval (**Extended Data Fig. 4o, p**). Accordingly, values derived from the 0-120 min OGTT data were used as proxy estimates in subsequent analyses. It should be noted that while the 0-120 min window largely preserves the relative magnitude of ***G*** across individuals, the absolute values are not preserved. We therefore used the 0-120 min estimates only as a reference in analyses for which only relative differences are relevant. These analyses included qualitative visualizations of interindividual variability and regression analyses in which we evaluated the predictive accuracy rather than the estimated regression coefficients.

Three-dimensional visualization of ***K*** revealed substantial heterogeneity in the P, I-like and D-like components of glycemic control even within the NGT group (**Fig. 1h**). Principal component analysis (PCA) supported this diversity: in NGT individuals, the first and second components accounted for only 46% and 34% of the variance, respectively (**Fig. 1j**). In contrast, individuals with T2DM showed reduced dimensionality, with variance concentrated more strongly along the first principal component. Comparison with a null distribution generated from randomly sampled data showed that the PC1 contribution rate was not greater than expected by chance in NGT individuals, whereas it was significantly increased in T2DM individuals (*P* < 0.01; **Methods; Extended Data Fig. 4q**). Thus, even among individuals with preserved glucose tolerance, glucose-regulatory capacity was distributed across multiple PID-like dimensions, whereas T2DM was associated with collapse toward a lower-dimensional regulatory state.

We next examined how variation in the PID-like components of ***K*** related to glucose tolerance. All three components (DI_/clep_, DI_/clei_, and DI_/cled_) were significantly reduced in individuals with IGT and T2DM compared with those with NGT (**Fig. 1k**). Logistic regression models incorporating these three components classified IGT and T2DM more accurately than models based on SPINA (SPINA-GBeta, SPINA-GR, and SPINA-DI) or HOMA/QUICKI indices (HOMA-β, HOMA-IR, and QUICKI) (**Fig. 1l-o**). Moreover, when the three components were entered simultaneously into multivariable logistic regression models, each independently contributed to the prediction of IGT. Correlation analysis with conventional metabolic indices further showed that DI_/clep_, DI_/clei_, and DI_/cled_ captured overlapping but non-identical physiological information (**Extended Data Fig. 4r**). These results indicate that deterioration of glucose tolerance can arise through different combinations of impaired proportional, integral-like, and derivative-like regulatory capacity across individuals. Thus, no single component fully explained glycemic dysregulation. Instead, simultaneous assessment of all three ***K*** components provides a more complete representation of the physiological heterogeneity underlying glucose tolerance.

We then compared ***K*** with the conventional clamp-derived DI (clamp DI), which primarily reflects first-phase insulin secretion and insulin sensitivity. While clamp DI showed a strong correlation with DI_/cled_ (R = 0.89, 95% CI: 0.84–0.92), its correlation with DI_/clep_was only moderate (R = 0.59, 95% CI: 0.46–0.70) (**Extended Data Fig. 4s**), suggesting that the conventional index does not fully capture the multidimensional nature of glycemic regulation. We assessed the predictive utility of ***K*** for post-OGTT glucose levels relative to clamp DI. Across all time points, linear regression models based on ***K*** consistently showed stronger correlations and lower Akaike Information Criterion (AIC) values compared to those using clamp DI (**Extended Data Fig. 4t, u**). Collectively, these results indicate that ***K*** provides a more comprehensive assessment of an individual’s ability to control blood glucose by capturing multiple dimensions of glucose regulation that are not fully captured by the traditional DI.

### Inferring clamp-derived *K* from OGTT profiles within the *F* = *K* · *G* framework

To assess the clinical applicability of *F* = ***K*** · ***G***, we tested whether clamp test-derived glucose regulatory capacity (***K***) could be predicted from glucose profiles (***G***) during OGTT. We compared two classes of OGTT-derived predictors. The first used glucose waveform features (***G***) alone, corresponding to glucose-only prediction. The second used the conventional OGTT-derived disposition index, oral DI, which is calculated from both glucose and insulin measurements during OGTT. Oral DI was included as a reference predictor because it is a widely used insulin-requiring benchmark for beta-cell function adjusted for insulin sensitivity. Thus, this comparison tested whether glucose-only waveform information could predict clamp-derived regulatory-capacity components with accuracy comparable to a conventional index requiring insulin sampling.

Given that the oral DI primarily reflects first-phase insulin secretion, we hypothesized that it would correlate most strongly with DI_/cled_, while providing limited information on DI_/clep_and DI_/clei_. We therefore evaluated prediction performance separately for each of the three clamp-derived components. Consistent with this hypothesis, models based solely on ***G*** during OGTT achieved prediction accuracies comparable to those obtained using the oral DI (**Fig. 1p, q**). The ***G***-based models outperformed oral DI in estimating DI_/clep_, suggesting that this regulatory dimension cannot be fully captured by oral DI alone. Although these findings constitute a proof of concept rather than a rigorous validation, these results suggest that under standardized conditions (fixed *F* and near-uniform ingestion rate), glucose dynamics features (***G***) alone can empirically predict the vector of glucose regulatory capacities (***K***), reducing the need for repeated insulin sampling.

### Inferring *K* from glucose dynamics using *F* = *K* · *G*

Because ***K*** has three components, it should, in principle, be estimable from ***G*** and *F* when data from at least three distinct glucose input conditions are available. Although the equation *F* = ***K*** · ***G*** was originally derived from clamp experiments, it is important to note that *F* represents the integrated glucose input rather than its temporal administration pattern. Furthermore, the DIs estimated from the OGTT profiles were significantly correlated with those derived from the clamp data (**Fig. 1p, q**). Building on this, we hypothesized that a similar structural relationship would hold under physiological conditions, such as during a meal when glucose enters the circulation via alternative routes. Specifically, despite differences in the route of administration and changes in the absolute values of the parameters^30^, the structure of the mathematical model and the relative ordering of the parameter values would remain consistent.

To test this hypothesis and to assess the consistency of our simplified formulation with an established, more complex mathematical model of daily postprandial glucose dynamics^31^, we performed large-scale *in silico* simulations (**Fig. 2a**). As previously described^31^, parameters for insulin sensitivity (V_mx_) and first-phase insulin secretion (K) were systematically varied to cover a physiological range including both NGT and T2DM. Each simulated individual underwent both rapid (bolus) and prolonged (continuous) glucose administration (**Fig. 2b, Extended Data Fig. 5a**). For each individual, we estimated the ***K*** vector from the blood glucose dynamics (***G***) and evaluated the correlation between the predicted ***K*** and V_mx_K values. Given that K mathematically corresponds to *k*_Sd_, we hypothesized that V_mx_K would exhibit the strongest correlation with DI_/cled_.

**Fig. 2.**
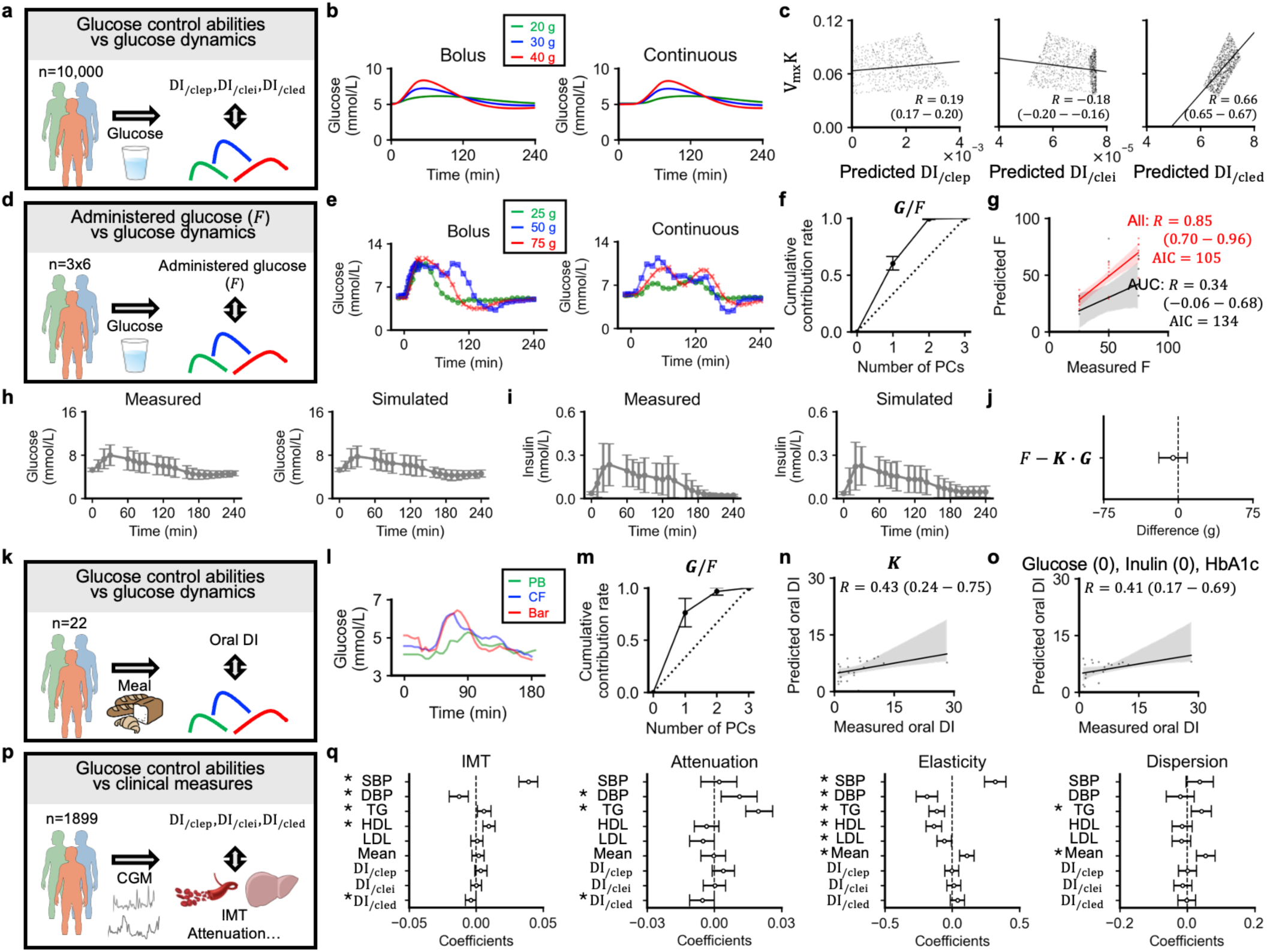
Relationship between glucose control abilities, administered glucose, and glucose dynamics. **a,** Schematic of *in silico* simulations evaluating the relationship between glucose control abilities and glucose dynamics across 10,000 human individuals. **b,** Examples of simulated blood glucose trajectories following the oral administration of 20 g, 30 g, or 40 g glucose, delivered either as a bolus or continuously. **c,** Correlations between predicted ***K*** components (DI_/clep_, DI_/clei_, and DI_/cled_) and ground-truth VₘₓK values (the product of insulin sensitivity and first-phase insulin secretion). Spearman’s correlation coefficients (*R*) with 95% confidence intervals are shown. For clarity, 1,000 individuals randomly subsampled from the full dataset are plotted; all statistics (correlation coefficients, 95% confidence intervals, and regression lines) were computed on all 10,000 individuals. **d,** Schematic of oral glucose tolerance test experiments with varying glucose loads and administration patterns. Data are from the *University of Tokyo Study*. **e,** Examples of plasma glucose trajectories following the oral ingestion of 25 g, 50 g, or 75 g glucose as a bolus or continuous input. **f,** Cumulative proportion of variance explained by the principal components of ***G***/*F* (the glucose-dynamics vector normalized by administered glucose). Data are mean ± SD across individuals. **g,** Relationship between the administered glucose dose (*F*) and the predicted *F* using either the AUC-only model (*F* = *K* · *G*_AUC_) or the full model (*F* = ***K*** · ***G***; “All”). Both axes are in grams; *F* is a concentration equivalent in the model and is converted for display. Shaded areas are 95% CIs. **h,** Mean measured and simulated plasma-glucose trajectories. Data are mean ± SD across 18 glucose-challenge profiles. **i,** Mean measured and simulated serum-insulin trajectories from the same fitted model as in (h). Data are mean ± SD across 18 glucose-challenge profiles. **j,** Difference between the administered glucose input *F* and the model-derived estimate ***K*** · ***G***. The residual was converted to grams using the glucose distribution volume and body weight. For this analysis, ***K*** was not estimated from *F* = ***K*** · ***G***. Instead, the full ODE model was fitted to the observed glucose and insulin trajectories for each challenge profile, and ***K*** was calculated from the fitted physiological parameters. ***G*** was calculated independently from the observed glucose trajectory, and the resulting ***K*** · ***G*** was then compared with the administered glucose load *F*. The point indicates the mean residual across 18 glucose challenge profiles, and the horizontal bar indicates the 95% confidence interval. The vertical dashed line indicates zero residual. **k,** Schematic of humans undergoing mixed meal tolerance tests. Data are from the *San Francisco Bay Area Study1*. **l,** Examples of plasma glucose trajectories following the ingestion of meals with different macronutrient compositions: PB (bread and peanut butter), CF (cornflakes and milk), and Bar (protein bar). **m,** Cumulative proportion of variance explained by the principal components of ***G***/*F*. Data are mean ± SD across individuals undergoing mixed meal tolerance tests. **n,** Predicted versus measured oral disposition index (oral DI) using the ***K*** components as predictors; each point is one participant. Shaded band, 95% CI. **o,** As in (n) but using fasting plasma glucose [Glucose (0)], fasting serum insulin [Insulin (0)], and HbA1c as predictors. **p,** Schematic of humans undergoing continuous glucose monitoring (CGM). Associations between estimated ***K*** (DI_/clep_, DI_/clei_, and DI_/cled_) and clinical measures were assessed. These measures included carotid intima-media thickness (IMT) and liver ultrasound-derived measures, such as attenuation, elasticity, and dispersion. Data are from the *HPP Cohort*. **q,** Multivariable linear regression of each vascular/hepatic outcome (IMT, attenuation, elasticity, dispersion) on a predictor set comprising DI_/clep_, DI_/clei_, DI_/cled_ and covariates (SBP, systolic blood pressure; DBP, diastolic blood pressure; TG, triglycerides; HDL and LDL cholesterol; Mean, mean glucose). Points are standardized regression coefficients; bars are 95% CIs.

As expected, DI_/cled_, derived from the *F* = ***K*** · ***G***, showed a strong correlation with the true V_mx_K values (**Fig. 2c**), supporting the accuracy of the model and its utility in inferring regulatory capacity from glucose dynamics and dosing. Furthermore, the temporal distortion feature of ***G*** was significantly correlated with the temporal autocorrelation-derived index (AC_Mean) (**Extended Data Fig. 5b**), suggesting that the three components of ***G*** (area under the curve, temporal distortion, and amplitude) are conceptually consistent with previously reported glucose dynamics-derived measures (mean, temporal autocorrelation, and variance), which have been independently associated with the DI and diabetes complications^5,6,9^.

### Inferring *F* from glucose dynamics using *F* = *K* · *G*

To further evaluate the equation, we analyzed a previously published human dataset (the *University of Tokyo Study*)^32^ reporting plasma glucose responses to three oral glucose loads (25 g, 50 g, and 75 g) delivered either as a bolus or continuously (**Fig. 2d, e**). Although ***G*** is defined in three dimensions, the geometric interpretation of *F* = ***K*** · ***G*** (**Fig. 1g**) predicts that ***G***/*F* for each individual should reside on a two-dimensional plane. Consistent with this prediction, PCA of ***G***/*F* showed that the first two components explained 99.5% of the total variance (**Fig. 2f**). This cumulative contribution was significantly greater than expected under a null model in which the three components of ***G***/*F* were statistically independent (*P* < 0.01; **Extended Data Fig. 5c**). Although this PCA result should be interpreted as supporting evidence rather than as definitive proof of the equation, it does support the geometric structure of the model.

We next tested whether the framework could be used to infer total glucose input (*F*) from observed glucose dynamics. Linear regression models using only *G*_AUC_ yielded weak and non-significant predictions (*R* = 0.34; 95% CI: –0.06 to 0.68), whereas models including the full ***G*** vector achieved much higher predictive accuracy (*R* = 0.85; 95% CI: 0.70 to 0.96) (**Fig. 2g**). These results suggest the necessity of considering multiple dimensions of glucose dynamics to accurately infer both regulatory capacity and time-integrated glucose input.

We then directly evaluated the equation by comparing the administered glucose load *F* with ***K*** · ***G***, where ***K*** and ***G*** were estimated from the observed glucose and insulin trajectories for each glucose challenge profile. The fitted mathematical model reproduced the overall glucose and insulin responses, including the post-ingestion rise and subsequent decline toward baseline (**Fig. 2h, i**). The mean residual *F* − ***K*** · ***G*** was −5.28 g and was not significantly different from zero (**Fig. 2j**). Together, these results support the approximate validity of *F* = ***K*** · ***G*** in human oral glucose challenge data.

### Inferring *K* from CGM data using *F* = *K* · *G*

We then investigated whether glucose regulatory capacity could be estimated from continuous glucose monitoring (CGM) data during mixed meal tolerance tests^33^ using the *F* = ***K*** · ***G*** equation. Although the equation was developed for glucose-specific inputs and does not explicitly account for lipid or amino acid effects, we tested whether the ***K*** vector derived from interstitial glucose profiles under real-life meal conditions could approximate the oral DI. Interstitial glucose trajectories (***G***) and carbohydrate intake (*F*) were measured after three meals with different macronutrient compositions (**Fig. 2k, l**).

PCA of ***G***/*F* for each individual showed that the first two components accounted for 96.7% of the variance (**Fig. 2m**). This cumulative contribution was significantly greater than expected under a null model in which the three components of ***G***/*F* were statistically independent (*P* < 0.01; **Extended Data Fig. 5d**), providing empirical support for the theoretical framework even under free-living conditions. In linear regression models, the CGM-derived ***K*** vector had comparable predictive performance to the model that used markers requiring blood sampling (fasting plasma glucose, fasting serum insulin, and HbA1c) in predicting oral DI (**Fig. 2n, o**). This comparison is conservative for the CGM-derived model, because oral DI itself is calculated using fasting glucose and insulin, creating partial overlap between the target variable and the blood-based predictors. Thus, the comparable performance of CGM-derived features, despite using no insulin measurements, supports the utility of the *F* = ***K*** · ***G*** framework for minimally invasive estimation of glucose-regulatory capacity from CGM data.

### Inferring diabetes-related phenotypes using CGM-derived ***K***

To examine the clinical significance of the *F* = ***K*** · ***G*** framework, we first tested whether glucose control capacity (***K***), estimated from free-living CGM data, was associated with vascular and hepatic health independently of conventional clinical measures by analyzing data from the *Human Phenotype Project (HPP) Cohort*^18,34,35^ (**Fig. 2p**). Using the equation *F* = ***K*** · ***G***, we estimated individual ***K*** values from CGM-recorded glucose dynamics and dietary carbohydrate intake. We then assessed the relationship between ***K*** values and vascular/hepatic measures, including intima-media thickness (IMT) and ultrasound-based hepatic attenuation, elasticity and dispersion. Systolic and diastolic blood pressure (SBP, DBP), triglycerides (TG), high-and low-density lipoprotein cholesterol (HDL, LDL) and mean glucose levels (Mean) were included as covariates.

Multiple linear regression analysis showed that DI_/cled_ was significantly associated with both IMT (β = –0.0040, 95% CI: –0.0080 to 0.0000) and liver attenuation (β = –0.0054, 95% CI: – 0.0110 to 0.0000) independently of other clinical measures (**Fig. 2q**). In contrast, mean glucose levels were independently associated with liver elasticity and dispersion. These results indicate that CGM-derived ***K*** captures aspects of vascular and hepatic phenotypes that are not fully explained by mean glycemia or standard clinical risk factors.

We next tested whether CGM-derived ***K***-related indices could predict metabolic and microbiome-associated phenotypes in an independent dataset. Using the *CGMacros Dataset*^36^, we evaluated the classification of glucose intolerance and microbial diversity status from CGM-derived ***K*** and compared their performance with that of conventional fasting blood–based indices. Of note, greater gut microbial diversity/richness has generally been associated with favorable metabolic and health-related phenotypes^37^. Logistic regression models based on ***K***-derived features predicted both glucose intolerance and microbial diversity with accuracy comparable to, or greater than, models based on SPINA-derived indices or HOMA/QUICKI-derived indices, which require fasting blood measurements (**Extended Data Fig. 5e**), suggesting that CGM-derived ***K***-related indices provide predictive information comparable to established blood-based measures of glucose–insulin regulation.

We then investigated whether simplified glucose-dynamics features corresponding to ***G*** could predict diabetes-related outcomes when standardized meal-input information was unavailable. Using CGM data from 100 individuals with diabetes^38^, we evaluated the prediction of two outcomes: the presence of diabetic complications and the two-hour post-load glucose concentration. We used mean glucose, glucose standard deviation, and AC_Mean as a glucose-only representation of overall glucose exposure, variability, and temporal structure, respectively. This choice was motivated by their conceptual correspondence to the components of ***G***, including the observed association between temporal distortion and AC_Mean (**Extended Data Fig. 5b**). Models based on these summary glucose-dynamics features achieved accuracy comparable to or greater than models that used GluFormer^39^ embeddings derived from the same CGM traces (**Extended Data Fig. 5f, g**). These results suggest that the proposed framework not only recapitulates glucose, insulin, and C-peptide dynamics, but also estimates glucose control capacity and various diabetes-related indicators without the need for blood sampling.

### A biphasic glucose pattern is associated with higher glucose effectiveness (*k*_GluG_⁄*k*_GluQ_)

The equation *F* = ***K*** · ***G*** is not affected by *k*_GluG_⁄*k*_GluQ_, indicating that this parameter cannot be inferred from the formulation alone. To address this limitation, we sought to identify an interpretable feature of the OGTT glucose trajectory that could serve as a surrogate marker for *k*_GluG_⁄*k*_GluQ_. Given that *k*_GluG_⁄*k*_GluQ_ reflects glucose flux from *G* to *Q* during the ascending phase, and from *Q* to *G* after the peak, we hypothesized that individuals with higher *k*_GluG_⁄*k*_GluQ_ would show more buffered glucose excursions during both the ascending and descending phases (**Extended Data Fig. 1n**), thereby producing a more prominent biphasic pattern^15^.

To test this hypothesis, we applied hierarchical clustering to standardized OGTT glucose trajectories from 121 individuals, aiming to group participants by the shape of their glucose response curves (**Fig. 3a**). Cluster quality was assessed by silhouette analysis, which identified a four-cluster solution as optimal (**Fig. 3b**). Among these, cluster 3 was characterized by a biphasic glucose profile (**Fig. 3c, d**). Individuals in cluster 3 had significantly higher values of *k*_GluG_⁄*k*_GluQ_ compared to other groups (**Fig. 3e**). We then tested this association more directly by classifying OGTT profiles as biphasic or nonbiphasic based on the presence of a secondary rise after an initial decline in glucose levels (**Methods**). Individuals with biphasic glucose dynamics showed higher *k*_GluG_⁄*k*_GluQ_ than those with non-biphasic dynamics (**Extended Data Fig. 5h**). Although these findings are exploratory and have not been validated in an external dataset, and although biphasic glucose dynamics may also be influenced by factors other than *k*_GluG_⁄*k*_GluQ_, these results suggest that biphasic glucose dynamics may serve as a less invasive phenotypic marker of increased *k*_GluG_⁄*k*_GluQ_.

**Fig. 3.**
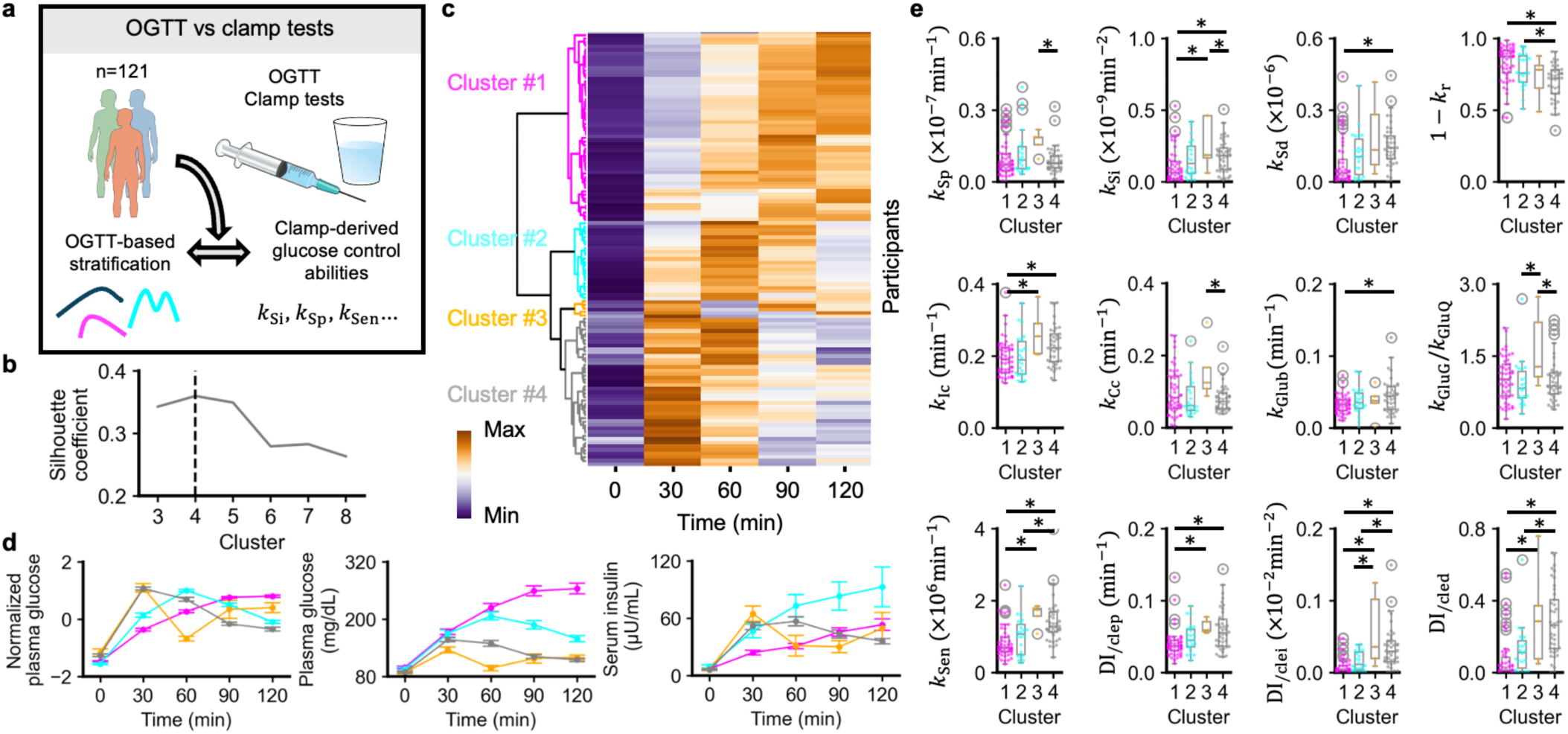
Associations between OGTT-derived glucose response profiles and clamp-derived glucose control abilities. **a,** Study design. Oral glucose tolerance tests (OGTT) and clamp data from 121 individuals (*Kobe University Study*) were used to relate OGTT response patterns to clamp-derived glucose control abilities. **b,** Selection of the number of clusters by silhouette analysis; the dashed line marks the number maximizing the silhouette coefficient. **c,** Heatmap showing the standardized plasma glucose responses during the OGTT (0–120 min) obtained by hierarchical clustering. Rows represent individual participants and columns represent time points. The color scale indicates z-scored glucose concentrations. **d,** Time courses of normalized plasma glucose (left), absolute plasma glucose (middle), and serum insulin (right) across the four identified clusters. Data are mean ± SEM. **e,** Box plots of *k*_Sp_, *k*_Si_, *k*_Sd_, 1 − *k*_r_, *k*_Ic_, *k*_Cc_, *k*_Glub_, *k*_GluG_⁄*k*_GluQ_, *k*_Sen_, DI_/clep_, DI_/clei_, and DI_/cled_ across clusters. Boxes show median and interquartile range; *Q* values were calculated using Welch’s *t*-tests with Benjamini-Hochberg correction. \**Q* < 0.05.

### Interrelationships among glucose-regulatory parameters

We next examined how glucose-regulatory parameters interrelate across different glucose-tolerance states to clarify potential compensatory mechanisms and their disruption. While both insulin sensitivity and secretion are known to decline with progression to T2DM, previous studies have shown that in individuals with NGT, reduced insulin sensitivity is often offset by increased insulin secretion^20,29,40,41^. Consequently, when analyzing a heterogeneous dataset, a positive correlation between insulin sensitivity and secretion would be expected, whereas within the NGT subgroup these parameters may instead display an inverse relationship. To date, however, these interrelationships, including those involving glucose effectiveness (*k*_GluG_⁄*k*_GluQ_), have not been systematically clarified. To systematically explore these interdependencies, we constructed correlation networks using clamp-derived estimates across the dataset and within stratified subgroups (**Fig. 4a, b**). In each network, a node represents an estimated glucose-regulatory parameter, and an edge represents a statistically significant pairwise Spearman correlation between two parameters (absolute coefficient > 0.21; *Q* < 0.05 in the full dataset).

**Fig. 4.**
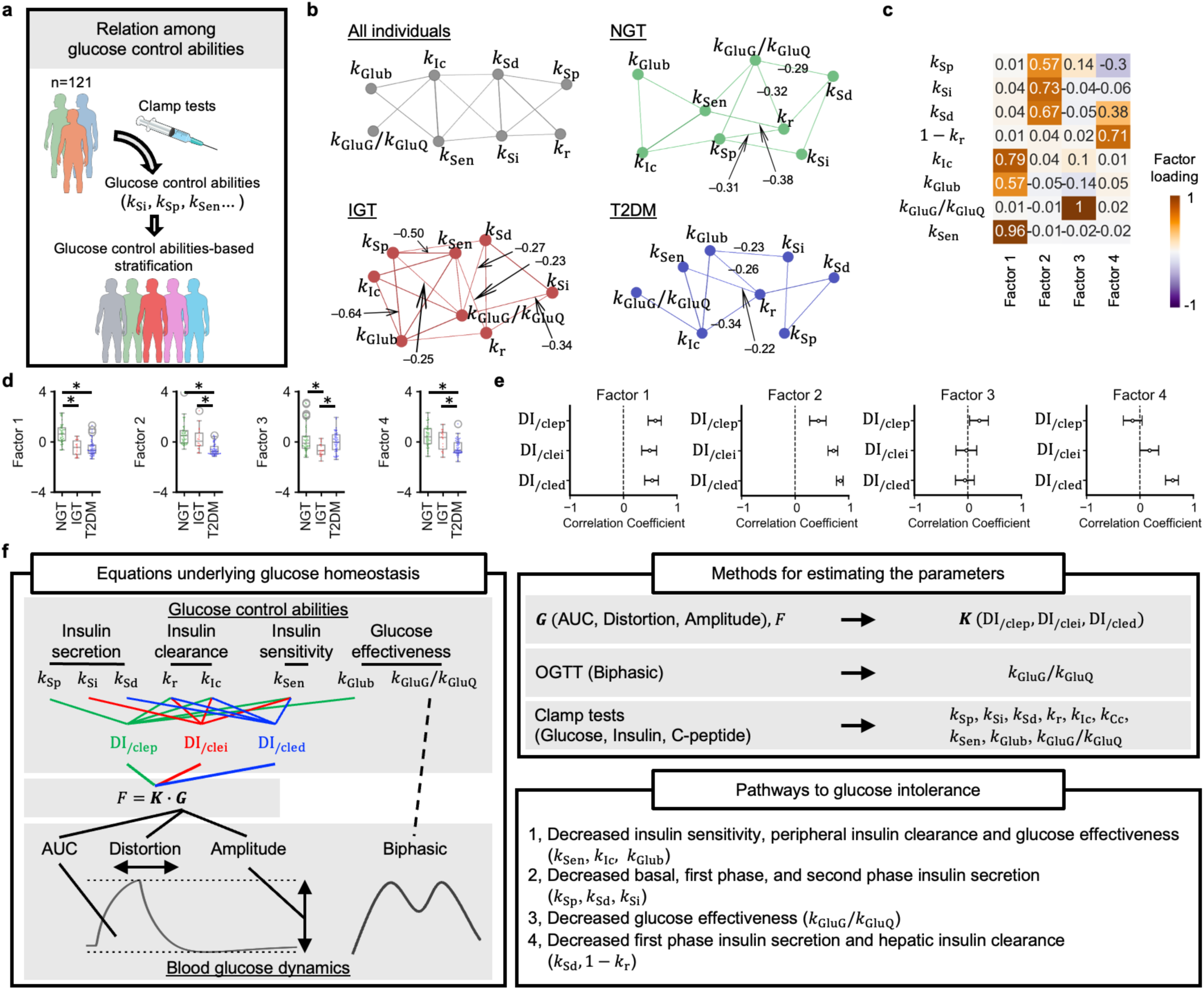
Interrelationships among glucose control abilities and pathways to glucose intolerance. **a,** Schematic overview of clamp-based assessments performed on 121 individuals from the *Kobe University Study* across a spectrum of glucose tolerance to examine interrelationships among components of glucose regulatory capacity. **b,** Correlation networks of regulatory parameters visualized using a spring layout. Edges connect parameter pairs with absolute Spearman’s correlation > 0.21 (*Q* < 0.05 for the entire dataset), with edge width reflecting correlation strength. Networks are shown for all participants (gray) and stratified by glucose tolerance group. For clarity, only negative correlations are annotated in subgroup networks. **c,** Heatmap of factor loadings from exploratory factor analysis. Rows represent individual parameters, and columns represent latent factors. Color encodes the magnitude and sign of each parameter’s contribution. **d,** Box plots of factor scores in the NGT, IGT, and T2DM groups. Boxes denote the median and interquartile range. *Q* values were determined using Welch’s *t*-tests with Benjamini–Hochberg correction. \**Q* < 0.05. **e,** Correlations of DI_/clep_, DI_/clei_, and DI_/cled_ with factors 1-4. Bars represent 95% confidence intervals of the correlation coefficients. **f,** A summary diagram showing the governing equation of glucose homeostasis, methods for parameter estimation, and major pathways leading to glucose intolerance.

In the combined dataset, most correlations were positive, reflecting coordinated dysregulation with glucose intolerance. These positive associations included a correlation between insulin sensitivity (*k*_Sen_) and peripheral insulin clearance (*k*_Ic_), consistent with previous reports showing that insulin clearance is positively associated with insulin sensitivity^42,43^. In contrast, stratified analyses revealed several negative correlations, particularly in the NGT and IGT subgroups. In NGT individuals, insulin sensitivity (*k*_Sen_) was negatively correlated with 1-hepatic insulin clearance (*k*_r_), suggesting that reduced insulin sensitivity may be partially compensated by suppressed hepatic insulin clearance to maintain systemic insulin levels. Furthermore, *k*_r_ showed negative correlations with both basal insulin secretion (*k*_Sp_) and glucose effectiveness (*k*_GluG_⁄*k*_GluQ_), suggesting additional possible compensatory adjustments involving hepatic handling of insulin. In IGT, negative correlations became more widespread, with *k*_Sen_ inversely correlated not only with *k*_r_ but also with *k*_Sp_. In contrast, individuals with T2DM showed fewer and weaker negative associations, particularly between insulin sensitivity and secretion parameters. Collectively, these results suggest that in IGT, multiple compensatory mechanisms involving insulin secretion, sensitivity, and clearance may exist, whereas in T2DM, such relationships appear to be disrupted, potentially contributing to progressive hyperglycemia.

### Multiple regulatory patterns associated with glucose intolerance

Group-wise comparisons confirmed that insulin sensitivity (*k*_Sen_), insulin secretion (*k*_Sp_, *k*_Si_, and *k*_Sd_), and glucose effectiveness (*k*_Glub_, *k*_GluG_⁄*k*_GluQ_) were significantly reduced in IGT or T2DM compared to NGT (**Extended Data Fig. 5i**). Insulin-dependent glucose-lowering effects were particularly decreased in these groups (**Extended Data Fig. 5j, Supplementary Note 14**). Despite these average trends, previous studies^20,29,40^ have indicated heterogeneity within each group, with some individuals showing disproportionately impaired insulin secretion, while others show more severe deficits in insulin sensitivity. For example, data-driven classifications of adult-onset diabetes, including the subgroups proposed by Ahlqvist et al., have used HOMA-derived indices to capture insulin secretion and insulin resistance^44^. However, HOMA-derived indices do not directly quantify insulin-independent glucose effectiveness and therefore may not distinguish hyperglycemia driven primarily by impaired glucose effectiveness from hyperglycemia driven by reduced insulin sensitivity. This limitation raises the possibility that a subtype characterized by reduced glucose effectiveness has been underrecognized in conventional classification frameworks. To investigate whether the *F* = ***K*** · ***G*** framework could identify latent patterns underlying this heterogeneity, we performed exploratory factor analysis on glucose-regulatory parameters that separately quantify insulin sensitivity, insulin secretion, hepatic and peripheral insulin clearance, and insulin-independent glucose effectiveness (**Fig. 4c**). Whereas the PCA in **Figure 1j** assessed the dimensionality of the three-component ***K*** vector itself, this factor analysis examined a broader set of model-derived physiological parameters to identify interpretable modules of covarying regulatory mechanisms.

The Very Simple Structure complexity criterion indicated that a four-factor solution best captured the structure of the data. Factor 1 primarily loaded on *k*_Sen_, *k*_Ic_, and *k*_Glub_. Factor 2 captured insulin secretion capacities (*k*_Sp_, *k*_Si_, and *k*_Sd_). Factor 3 was affected by glucose effectiveness (*k*_GluG_⁄*k*_GluQ_), while factor 4 was characterized by hepatic insulin clearance and first-phase secretion (1 − *k*_r_, *k*_Sd_). The Kaiser-Meyer-Olkin measure of sampling adequacy was 0.61, and Bartlett’s test of sphericity indicated statistically significant intercorrelations among the variables (*P* < 0.01), confirming the suitability of the dataset for factor analysis. All four factor scores were significantly reduced in IGT or T2DM compared to NGT (**Fig. 4d**), suggesting four latent dimensions of glucose-regulatory variation associated with glucose intolerance.

We next examined the correlations between the factors and DI_/clep_, DI_/clei_, and DI_/cled_ (**Fig. 4e**). Factor 1, reflecting insulin sensitivity (*k*_Sen_) and glucose effectiveness (*k*_Glub_), was significantly correlated with all three indices (DI_/clep_, DI_/clei_, and DI_/cled_), most strongly with DI_/clep_ (*R* = 0.60; 95% CI: 0.46 to 0.71), which is consistent with its strong loading on *k*_Glub_. Factor 2, representing *k*_Si_ and *k*_Sd_, showed strong associations with DI_/clei_ (*R* = 0.72; 95% CI: 0.62 to 0.80) and DI_/cled_(*R* = 0.85; 95% CI: 0.77 to 0.90). Factor 3, reflecting glucose effectiveness (*k*_GluG_⁄*k*_GluQ_), showed minimal correlation with any DI indices, as expected since it is not directly included in their calculation. Factor 4, characterized by high loadings on *k*_Sd_, was associated with DI_/cled_ (*R* = 0.62; 95% CI: 0.48 to 0.72). These distinct correlation patterns suggest that the impairment in DI_/clep_, DI_/clei_, and DI_/cled_ may reflect mechanistically divergent pathways of metabolic deterioration across individuals.

## DISCUSSION

Despite decades of efforts in mathematical modeling^10–14^ and machine learning^16,17,45^, a unified, mechanistically interpretable equation linking observable blood glucose dynamics to the underlying physiological processes governing glucose homeostasis has remained elusive. Here, we propose the relations underlying glucose homeostasis (**Fig. 4f**): (i) glucose dynamics and their regulatory parameters satisfy the equation *F* = ***K*** · ***G***, and (ii) individual differences exist within the range that satisfies this equation. ***K*** consists of three components reflecting the proportional (P), integral-like (I) and derivative-like (D) elements in control theory, while ***G*** is a three-dimensional vector consisting of the area under the curve, amplitude, and temporal distortion of glucose profiles. Although simple in form, this equation was derived from a system of six differential equations with thirteen parameters that reproduced the dynamics of glucose-insulin interactions during clamp tests and OGTTs.

This model theoretically enables minimally invasive estimation of regulatory capacity (***K***) from CGM data (***G***) and glucose input (*F*), without requiring blood sampling or insulin measurements. Previous studies have shown that conventional indices, such as fasting blood glucose levels, HbA1c or 2-hour OGTT values, primarily reflect mean glucose levels and are less predictive of glucose tolerance and risks of diabetes complications than CGM-derived indices including glucose variability and temporal autocorrelation^5,6,9^. However, these approaches have been correlative in nature and have not accounted for the administered glucose load. In contrast, the *F* = ***K*** · ***G*** equation provides a theoretically grounded method to estimate model-derived, physiologically interpretable regulatory-capacity indices by integrating both glucose dynamics and dosing.

From a clinical perspective, the framework has translational potential because it relies on CGM data, which are already widely used in daily diabetes care. It may complement existing diagnostic criteria and enable earlier detection of impaired glucose tolerance or candidate glucose-regulatory patterns not captured by conventional measures. It may also improve risk stratification for vascular and hepatic complications and enhance the predictive accuracy of clinical outcomes. Furthermore, by distinguishing mechanistic subtypes, such as secretion-deficient, resistance-dominant, or glucose-effectiveness-deficient phenotypes, the model may inform the choice of individualized therapeutic strategies and facilitate the identification of novel treatment targets. Because CGM can be repeatedly and relatively easily performed in daily clinical practice, changes in ***K*** components could, in principle, be monitored before and after lifestyle interventions or pharmacological therapies, although the within-individual reproducibility of ***K*** and its responsiveness to treatment were not assessed in this study. These applications suggest that the framework may provide a basis for future evaluation of precision-medicine applications in diabetes.

We evaluated the framework using multiple independent datasets, including clamp studies, OGTT data, mixed-meal tests, and CGM profiles. Nonetheless, several datasets had modest sample sizes, underscoring the need for larger, prospective studies to establish generalizability and clinical utility. Although the proposed framework can be applied to CGM data, estimation of ***K*** from free-living CGM profiles requires information on glucose input *F*, such as meal timing and nutrient intake, and should not be interpreted as a purely passive CGM-only method. Recent personalized-nutrition studies increasingly collect dietary records together with CGM data, including meal timing, meal composition, or food images^18,36^, demonstrating that joint monitoring of glucose trajectories and dietary inputs is becoming feasible in research settings. However, there is still no established physiological framework for integrating meal-derived inputs with CGM-derived glucose waveforms to infer glucose-regulatory capacity. The equation *F* = ***K*** · ***G*** provides one possible approach to this integration by relating estimated glucose input, glucose trajectory features, and regulatory capacity within a single interpretable model. When reliable meal information is unavailable, simpler descriptive CGM metrics, such as mean glucose, glucose variability, or temporal autocorrelation, may remain more appropriate for clinical or epidemiological analyses. Additional validation will also be required for the exploratory finding that biphasic OGTT dynamics may reflect *k*_GluG_⁄*k*_GluQ_. Although the observed correlations in the *HPP Cohort* were significant, they were modest. This is likely because the study mainly included healthy individuals with minimal inter-individual differences. Including individuals with diabetes may produce stronger correlations, and further research is needed. Building on this foundation, the model generates several testable predictions. For example, simultaneous measurement of glucose trajectories and clamp-derived parameters under variable glucose loads could provide empirical validation of the relationship. Moreover, while genetic and omics studies have traditionally focused on insulin sensitivity or secretion, the identification of individuals with impaired glucose effectiveness (*k*_GluG_⁄*k*_GluQ_) points to previously unexplored physiological mechanisms. Although previous studies have shown that glucose effectiveness decreases as glucose tolerance worsens^46^, most studies have quantified glucose effectiveness using the minimal model^47^, which mathematically approximates *k*_Glub_. While *k*_Glub_ correlated strongly with insulin sensitivity, *k*_GluG_⁄*k*_GluQ_ was largely independent of insulin sensitivity and secretion. Additionally, compensatory pathways, including insulin clearance, may also warrant further investigation. Genetic and omics investigations stratified by these factors could deepen our understanding of glucose homeostasis. Since PID-like feedback structures are effective at maintaining regulated variables near a set point, similar control architectures may underlie other homeostatic systems besides glucose homeostasis. If so, the perturbation inputs, regulatory capacities, and output waveforms of these systems may be related by equations analogous to *F* = ***K*** · ***G***. We emphasize, however, that this remains a possibility for future research.

In conclusion, the *F* = ***K*** · ***G*** equation provides a compact and interpretable representation of glucose homeostasis, linking glucose input, glycemic trajectories, and latent regulatory capacity. By integrating dosing information with waveform features, this framework moves beyond descriptive glycemic indices and offers a route toward risk stratification and precision medicine in diabetes.

## METHODS

### Study design and ethical approval

This study includes exploratory analyses of 11 independent studies. In this paper, these studies are referred to as the *Kobe University Study*^25^, the *Kobe University Study2*^6^, the *HuMet Repository*^48^, the *PREDICT1 Study*^49^, the *porcine intravenous glucose tolerance test (IVGTT) Study*^50^, the *University of Tokyo Study*^32^, the *San Francisco Bay Area Study1*^33^, the *San Francisco Bay Area Study2*^16^, the *CGMacros Dataset*^36^, the *Human Phenotype Project (HPP) Cohort*^18,34,35^, and the *Shanghai Study*^38^. Human studies were conducted in accordance with the Declaration of Helsinki and approved by the relevant institutional ethics committees. The animal study was approved by the appropriate animal experimentation committee and conducted in accordance with applicable animal-care guidelines. Written informed consent was obtained from all participants before enrollment.

The *Kobe University Study*^25^ is an observational study investigating the associations between glucose regulatory capacity and glucose dynamics. The primary objective of the analysis was to develop a mathematical model linking characteristics of glucose dynamics to underlying physiological mechanisms, such as insulin sensitivity, insulin secretion and clearance. The study was approved by the ethics committee of Kobe University Graduate School of Medicine and registered with the University Hospital Medical Information Network (UMIN000002359).

The *Kobe University Study2*^6^ is an observational study investigating glucose handling capacities and glucose dynamics in individuals without prior diagnosis of diabetes. The primary objective of the analysis in the present study was to evaluate the reproducibility and generalizability of the mathematical model in an independent clamp dataset. The study was approved by the ethics committee of Kobe University Hospital (Approval No. 1834; Kobe, Japan).

The *HuMet Repository*^48^ is a publicly available human dataset capturing temporal responses to multiple metabolic challenges, including a 75 g oral glucose tolerance test (OGTT). The primary objective of the analysis in the present study was to assess whether the mathematical model could reproduce plasma glucose and serum insulin dynamics during an OGTT in an external dataset. The underlying HuMet study was conducted at the Human Study Center of the Else-Kröner-Fresenius Center of Nutritional Medicine, Technische Universität München, and was approved by the ethics committee of the Technische Universität München (Approval No. 2087/08).

The *PREDICT1 Study*^49^ is a multinational, interventional clinical trial investigating postprandial metabolic responses to standardized mixed meals. The primary objective of the analysis in the present study was to assess whether the mathematical model could reproduce plasma glucose and serum insulin dynamics during a mixed-meal tolerance test (MMTT). The study was approved by the UK Research Ethics Committee (IRAS 236407). The current study analyzed glucose and insulin levels up to 4 h after the MMTT, which can be accessed at https://www.nature.com/articles/s41591-020-0934-0.

The *porcine IVGTT Study*^50^ is an experimental animal study characterizing glucose and insulin responses in pigs. The primary objective of the analysis in the present study was to assess whether the mathematical model could reproduce glucose and insulin dynamics under intravenous glucose perturbation beyond human datasets. The study was approved by the Ethics Committee for Animal Experimentation, Uppsala, Sweden (Permit C99/14), and followed the National Research Council Guide for the Care and Use of Laboratory Animals.

The *University of Tokyo Study*^32^ is an observational study investigating the relationship between administered glucose load and glucose dynamics. The primary objective of the analysis was to validate the mathematical model developed in this study by predicting the administered glucose amount from glucose dynamics data. The study was approved by the ethics committees of the Life-Science Committee of the University of Tokyo (16–265).

The *San Francisco Bay Area Study1*^33^ is an observational study investigating the relationship between continuous glucose monitoring (CGM)-derived measures and glucose control ability quantified by an oral glucose tolerance test (OGTT). The primary objective of the analysis was to predict the OGTT-derived disposition index from CGM-measured glucose dynamics. The study was approved by the Stanford Institutional Review Board (IRB 37141).

The *San Francisco Bay Area Study2*^16^ is an observational study investigating the relationship between glucose levels during OGTT and glucose control abilities. The primary objective was to investigate the consistency of glucose-derived indices calculated from OGTT data within the time frames of 0-120 minutes and 0-180 minutes. The study was approved by the Stanford University School of Medicine Human Research Protection Office (Institutional Review Board #43883).

The *CGMacros Dataset*^36^ is a publicly available observational study designed for personalized nutrition and diet monitoring in free-living conditions. The primary objective of the analysis in the present study was to evaluate whether CGM-derived glucose-response indices could classify glucose intolerance and gut microbial diversity status. Participants in the study were recruited at Sansum Diabetes Research Institute, Santa Barbara, CA. The study was approved by Advarra IRB (Pro00049227) and registered at ClinicalTrials.gov (NCT04991142). The present study analyzed de-identified, publicly available data (https://physionet.org/content/cgmacros/1.0.0/).

The *HPP Cohort*^18,34,35^ is an observational study investigating glucose fluctuations in non-diabetic individuals using CGM. The primary objective of the analysis was to assess the associations between CGM-predicted glucose control ability and carotid/liver ultrasound-derived measures. The study was approved by the Institutional Review Board of the Weizmann Institute of Science.

The *Shanghai Study*^38^ is an observational study investigating glucose fluctuations in diabetic individuals using CGM. The primary objective of the analysis was to assess the associations between CGM-derived glucose dynamics and diabetes-related measures. The dataset was collected as part of the Diabetes Data Registry and Individualized Lifestyle Intervention (DiaDRIL) project in Shanghai, China.

### Study population and inclusion criteria

Detailed inclusion and exclusion criteria, as well as comprehensive data acquisition protocols, have been reported previously^16,25,32–36,38,48–51^. A summary of the main criteria for each dataset is provided below.

The *Kobe University Study*^25^ enrolled individuals aged 20-80 years between October 2008 and December 2011. Exclusion criteria included renal impairment (creatinine clearance < 60 mL/min/1.73 m²), liver dysfunction (aspartate aminotransferase or alanine aminotransferase >50 IU/L), and ongoing insulin therapy. Of the 121 participants recruited, 50, 18, and 53 individuals met the criteria of normal glucose tolerance (NGT), impaired glucose tolerance (IGT), and type 2 diabetes mellitus (T2DM), respectively.

The *Kobe University Study2*^6^ enrolled adults aged 20 years or older without prior diagnosis of diabetes who were recruited at Kobe University Hospital between January 2016 and March 2018. Exclusion criteria included use of medications known to affect glucose metabolism (*e.g.*, steroids and β-blockers), psychiatric disorders, pregnancy or breast-feeding, and conditions deemed unsuitable for participation by the attending physicians. Of the 70 initially enrolled individuals, 6 were excluded because of β-blocker use (n = 1), missing continuous glucose monitoring data (n = 2), protocol deviations (n = 2), or missing data (n = 1), yielding a final analytic sample of 64 participants.

The *HuMet Repository*^48^ included 15 healthy male volunteers (mean age 27.8 ± 2.9 years; mean body mass index 23.1 ± 1.8 kg/m²) who were free of medications and metabolic abnormalities. All 15 participants completed the OGTT challenge and were included in the present analysis.

The *PREDICT1 Study*^49^ recruited generally healthy adults aged 18–65 years between 5 June 2018 and 8 May 2019. Exclusion criteria included ongoing inflammatory disease; cancer within the previous three years (excluding skin cancer); long-term gastrointestinal disorders including inflammatory bowel disease or celiac disease (but not irritable bowel syndrome); daily use of immunosuppressants or antibiotics within the prior three months; a capillary glucose >12 mmol/L (>216 mg/dL), known type 1 diabetes, or current use of medications for type 2 diabetes; acute clinically diagnosed depression; myocardial infarction or stroke within the previous six months; pregnancy; veganism; and eating disorders. Of the participants who completed the in-clinic MMTT, plasma glucose and serum insulin data from 903 participants were used in the present analysis.

The *porcine IVGTT Study*^50^ used 18 high-health pigs from two litters obtained from the Swedish Livestock Research Centre (Lövsta). Litter 1 (n = 8; 4 females and 4 males; Yorkshire × Swedish Landrace) had a mean body weight of 17.1 ± 0.4 kg at approximately 2 months of age, and Litter 2 (n = 10; 5 females and 5 males; Yorkshire × Swedish Landrace × Hampshire) had a mean body weight of 32.7 ± 0.8 kg at approximately 3 months of age. The pigs were housed individually in 3-m² pens with straw and wood-shavings bedding within sight and sound of one another under a 14:10 light/dark cycle, and were fed a commercial finisher diet (SOLO 330; Lantmännen, Sweden) without growth promoters or antimicrobials twice daily, with water available *ad libitum*. All 18 pigs underwent the IVGTT and were included in the present analysis.

The *University of Tokyo Study*^32^ enrolled individuals aged 20-50 years with HbA1c levels between 4.9% and 5.4% who had not been diagnosed with diabetes. A total of three participants were included in the study.

The *San Francisco Bay Area Study1*^33^ included individuals aged 25-76 years with no prior diagnosis of diabetes. Exclusion criteria included major organ diseases, chronic inflammatory diseases, malignancy, uncontrolled hypertension, eating disorders, a history of bariatric surgery, current use of weight-loss or diabetogenic medications. Of the 57 participants screened, 22 who completed both CGM during three standardized meals and an OGTT were included in the analysis.

The *San Francisco Bay Area Study2*^16^ included individuals aged 30-70 years with a body mass index of 23-40 kg/m^2^. Exclusion criteria included a history of diabetes, major organ diseases, uncontrolled hypertension, malignancy, chronic inflammatory diseases, use of any medications known to alter blood glucose or insulin sensitivity, hematocrit < 30%, serum creatinine above the normal reference range, and alanine aminotransferase levels more than twice the upper limit of normal. Individuals with missing values were excluded from the analysis. Of the 56 participants screened, 51 met the inclusion criteria. After screening tests, 8, 27, and 16 individuals met the criteria of T2DM, IGT, and NGT, respectively.

The *CGMacros Dataset*^36^ included 45 participants aged 18-69 years with a body mass index of 21–46 kg/m². For the present classification analysis, glucose intolerance was defined as HbA1c ≥ 6.0%, and participants with HbA1c < 6.0% were classified as not having glucose intolerance. Based on this definition, 22 participants were classified as having glucose intolerance and 23 participants as not having glucose intolerance. Exclusion criteria for participants with type 2 diabetes included treatment with oral medications other than metformin or treatment with injectable GLP-1 receptor agonists or insulin. All participants who completed the study and had available CGM, meal macronutrient, blood analytics, and gut health profile data were considered for the present analysis. Participants with missing values in the variables required for each prediction task were excluded from the corresponding analysis.

The *HPP Cohort*^18,34,35^ enrolled individuals aged 40-70 years between January 2019 and May 2023. Participants who underwent an ultrasound examination and had a food record were included in the analysis. Exclusion criteria included self-reported diabetes and use of diabetes-related medications. Of the 9,529 participants at baseline, 1,899 met the inclusion criteria.

The *Shanghai Study*^38^ enrolled individuals diagnosed with diabetes according to World Health Organization criteria. The participants were recruited at Shanghai East Hospital (September 2019–March 2021) and Shanghai Fourth People’s Hospital (June 2021–November 2021). This study excluded individuals with type 1 diabetes, leaving 100 participants.

### Clinical measures and assessments

Detailed descriptions of the clinical assessments have been published previously^16,25,32–36,38,48–51^. The main methods used for each dataset are summarized below.

In the *Kobe University Study*^25^, a hyperglycemic clamp was performed by intravenous infusion of a glucose bolus for 15 minutes, followed by a variable-rate glucose infusion to maintain a plasma glucose concentration of 200 mg/dL. This was followed, after a 10-minute interval, by a hyperinsulinemic-euglycemic clamp in which insulin was infused to achieve a plasma glucose concentration of 90 mg/dL or a fasting plasma glucose level in participants with NGT or IGT whose baseline glucose was below 90 mg/dL. Plasma glucose, serum insulin, and serum C-peptide were sampled before and at 5, 10, 15, 60, 75, 90, 100, 190, and 220 minutes after the onset of the clamp protocol. Plasma glucose was additionally measured every minute during the clamp and averaged over 5-minute periods. Both clamp procedures and a 75 g OGTT were performed within a 10-day window. Missing values were imputed using the k-nearest neighbor algorithm (k=2). Insulin secretion was quantified as the incremental area under the insulin curve during the first 10 minutes of the hyperglycemic clamp. The insulin sensitivity index ^25^ was calculated as the average glucose infusion rate during the last 30 minutes of the hyperinsulinemic-euglycemic clamp, normalized to plasma glucose and serum insulin concentrations. The clamp-derived disposition index (Clamp DI) was defined as the product of insulin secretion and sensitivity. The OGTT-derived disposition index (Oral DI) was calculated as the product of the insulinogenic index and the Matsuda index, as previously described^6,52,53^. SPINA-GR, SPINA-GBeta, SPINA-DI, HOMA-β, HOMA-IR, and QUICKI were calculated as previously described^54,55^.

In the *Kobe University Study2*^6^, consecutive hyperglycemic and hyperinsulinemic–euglycemic clamp tests were performed using an artificial endocrine pancreas (STG-55; Nikkiso Co., Ltd, Tokyo, Japan). The hyperglycemic clamp consisted of an intravenous glucose bolus of 9622 mg/m² infused over 15 minutes, followed by a variable-rate glucose infusion to maintain plasma glucose at 200 mg/dL for a total of 90 minutes. After a 10-minute washout interval, a hyperinsulinemic–euglycemic clamp was performed by intravenous infusion of human regular insulin (Humulin R; Eli Lilly Japan K.K., Kobe, Japan) at 40 mU·m⁻²·min⁻¹ for 120 minutes, with the variable-rate glucose infusion adjusted to maintain plasma glucose at the fasting level (target serum insulin concentration of 100 μU/mL). Plasma glucose was measured every minute during the clamp and averaged over 5-minute periods. Serum insulin and serum C-peptide were sampled before and at 5, 10, 15, 60, 75, 90, 100, 190, and 220 minutes after the onset of the clamp protocol.

In the *HuMet Repository*^48^, the OGTT challenge was performed on day 3 of the study blocks. After an overnight fast, participants ingested 300 mL of a glucose solution containing mono- and oligosaccharides equivalent to 75 g of glucose after enzymatic cleavage, consumed within 5 minutes. Plasma was sampled at 0, 15, 30, 45, 60, 90, 120, 180, and 240 minutes post-ingestion. Plasma glucose was measured by an enzymatic amperometric technique, and serum insulin was measured by enzyme-linked immunosorbent assay. Time-course data of plasma glucose and serum insulin during the 75 g OGTT were downloaded from the publicly available HuMet Repository (https://humet.org) and used for the present analysis.

In the *PREDICT1 Study*^49^, participants attended an in-clinic visit at 08:30 after an overnight fast (from 21:00 the previous night) and after refraining from exercise and limiting fat, fiber, and alcohol intake for 24 hours and from caffeine after 18:00 the previous evening. After cannulation of a forearm antecubital vein and collection of a baseline fasting sample, participants consumed a standardized breakfast meal of muffins and a milkshake providing 86 g of carbohydrate and 53 g of fat within 10 minutes. Venous blood was sampled at 0 (fasting), 15, 30, 60, 120, 180, and 240 minutes after breakfast. Plasma glucose was measured by the glucose oxidase method on a Siemens ADVIA 1800 analyzer, and serum insulin and C-peptide were measured by two-site sandwich immunoassay on an ADVIA Centaur XP. Publicly released time-course data of plasma glucose and serum insulin during the post-breakfast window of the standardized mixed-meal tolerance test were used for the present analysis.

In the *porcine IVGTT Study*^50^, each animal underwent a two-week acclimatization period that included training to step onto an electronic scale and to bottle-feed glucose solution. Under aseptic conditions and general anesthesia, an indwelling silicone catheter (SIL-C70 with rounded tip; Instec Solomon, Plymouth Meeting, PA) was implanted in the jugular vein and tunneled subcutaneously to an exit point between the scapulae. For the IVGTT, glucose (Glucose Fresenius Kabi, 500 mg/mL) was administered as an intravenous infusion of 0.5 g per kg body weight over 1 minute. Blood samples were collected before the glucose load and at 5, 10, 20, 30, 45, 60, 90, 120, and 180 minutes after the infusion. Blood glucose was measured with Accu-Chek test strips (Roche) validated for porcine blood, and plasma insulin was measured by porcine-specific enzyme-linked immunosorbent assay (10-1200-01; Mercodia, Uppsala, Sweden). Publicly released time-course data of plasma glucose and serum insulin during the IVGTT were used for the present analysis.

In the *University of Tokyo Study*^32^, a glucose solution containing 25 g, 50 g, or 75 g of glucose was administered orally after a 10-hour fast. Blood samples were collected from a superficial forearm vein at 0 minutes and every 10 minutes up to 240 minutes after ingestion. Two ingestion patterns were used: rapid bolus ingestion (completed within 1 minute) and continuous ingestion over two hours. For the latter, a non-contact micro dispensing robot delivered the glucose solution through tubes held in the participants’ mouths and ingested continuously over the two-hour period. All glucose solutions were diluted with distilled water to a final volume of 225 mL to standardize ingestion. Each participant completed all three glucose doses and two ingestion methods, with test sessions separated by at least one month. To characterize the dynamics of blood glucose following administration, three distinct indices (*G*_AUC_, *G*_Distortion_, and *G*_Amplitude_) were computed for each condition. *G*_AUC_ was calculated by integrating the linearly interpolated, baseline-corrected glucose curve. *G*_Distortion_ reflects temporal distribution of glucose exposure. *G*_Amplitude_ represents the amplitude of the glucose response.

In the *San Francisco Bay Area Study1*^33^, participants consumed three standardized breakfast meals on separate occasions: (i) bread with peanut butter, (ii) an energy bar, and (iii) cereal with milk and raisins. Each meal was eaten twice on non-consecutive days, and participants wore continuous glucose monitors throughout. Data from those with incomplete records or who consumed non-standardized items were excluded. The Oral DI was calculated as the product of the insulinogenic index and the Matsuda index, as previously described^6,52,53^.

In the *San Francisco Bay Area Study2*^16^, participants were instructed to maintain their usual diet and physical activity for three days prior to the OGTT, to fast for 10-12 hours, to abstain from strenuous physical activity after 17:00 the previous evening, and to abstain from smoking or nicotine use on the morning of the test. Blood samples were taken at 16 time points (−10, 0, 10, 15, 20, 30, 40, 50, 60, 75, 90, 105, 120, 135, 150 and 180 minutes) via an antecubital intravenous catheter. Plasma glucose concentrations were measured by the oxidase method. Analyses in this study were limited to 15 of these time points (0-180 minutes, excluding the -10-minute baseline).

In the *CGMacros Dataset*^36^, glucose values from the Abbott FreeStyle Libre Pro CGM were analyzed. For each annotated meal, CGM values were extracted from the recorded mealtime to 180 min after meal onset at 15-min intervals. To focus on meals with sufficient carbohydrate exposure, meals with carbohydrate-derived energy of 60 kcal or less were excluded. SPINA-GR, SPINA-GBeta, SPINA-DI, HOMA-β, HOMA-IR, and QUICKI were calculated as previously described^54,55^. Gut microbial diversity status was assessed using the Active Microbial Diversity score obtained from the Viome-derived gut health profile (Viome Life Sciences, Inc.).

In the *HPP Cohort*^18,34,35^, ultrasound assessments were performed to measure carotid intima-media thickness (IMT), liver attenuation, elasticity, and dispersion. Systolic blood pressure (SBP), diastolic blood pressure (DBP), triglycerides (TG), low-density lipoprotein cholesterol (LDL-C), and high-density lipoprotein cholesterol (HDL-C) measured at the time of the CGM assessment were also included in the analyses.

In the *Shanghai Study*^38^, interstitial glucose levels were recorded every 15 minutes using a CGM device for at least three days. Glucose profile characteristics from the first three days of CGM data for each participant were extracted. Trained research staff collected demographic and clinical information, including diabetes diagnosis and complications, using questionnaires. The study also investigated two-hour postprandial plasma glucose levels. Missing values were excluded from the analysis.

### Mathematical modeling of plasma glucose, serum insulin and C-peptide dynamics

To characterize the temporal dynamics of plasma glucose (*G*), remote glucose (*Q*), remote insulin (*X*), serum insulin (*I*), serum C-peptide (*C*), and remote C-peptide (*Y*), the following system of differential equations, based on previously published models, was used (**Supplementary Notes 1, 2, Extended Data Fig. 1**):

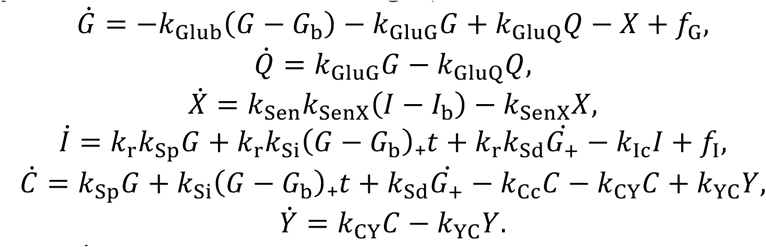

Here, *k*_Glub_ and *k*_GluG_⁄*k*_GluQ_ represent different aspects of glucose effectiveness. The parameters *k*_Sp_, *k*_Si_, and *k*_Sd_ represent the basal, potentiated, and first-phase components of insulin secretion, respectively. The parameters 1 − *k*_r_, *k*_Ic_, and *k*_Cc_ denote hepatic insulin clearance, peripheral insulin clearance, and C-peptide clearance, respectively. *Ġ*_+_ equals zero when *G* is decreasing and equals *Ġ* when *G* is increasing. The functions *f*_G_ and *f*_I_ represent exogenous glucose and insulin inputs, respectively. In clamp experiments, *f*_G_ and *f*_I_ were treated as known forcing functions and were directly specified from the administered glucose and insulin infusion profiles after linear interpolation between recorded time points. In contrast, during oral glucose administration, the systemic rate of glucose appearance was not directly observed; therefore, *f*_G_ was estimated from the glucose and insulin time-course data using a piecewise-linear function, following the framework of the oral minimal model^13^. In this setting, the time integral of *f*_G_ was constrained according to the administered oral glucose dose, while the parameters governing glucose and insulin dynamics were estimated simultaneously. When no exogenous insulin was administered, *f*_I_ was set to zero.

Parameter estimation was performed using a meta-evolutionary programming algorithm combined with non-linear least squares optimization, minimizing the normalized residual sum of squares between the measured data and the model predictions, as previously described^51,52,56^. This simulation was conducted using SciPy 1.10.1^57^.

The following equation was derived by integrating the differential equations between steady states (**Supplementary Note 12**):

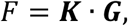

where *F* denotes the integrated glucose input over the analysis window, and ***K*** represents glucose regulatory capacity and is defined as: ***K*** = :*k*_Glub_ + *k*_Sen_*k*_r_*k*_Sp_⁄*k*_Ic_, *k*_Sen_*k*_r_*k*_Si_⁄*k*_Ic_, *k*_Sen_*k*_r_*k*_Sd_⁄*k*_Ic_;. The ***G*** is a composite vector summarizing blood glucose dynamics: ***G*** = (∫(*G* − *G*_b_)*dt*, ∫(*G* − *G*_b_)h*tdt*, *G*_max_ − *G*_min_). For each glucose challenge, the analysis window was defined from the start of glucose administration or meal ingestion. The derivation assumes that the cumulative exposure is reset at the start of each challenge and, when the equation is interpreted between steady states, that glucose and insulin-related state variables are close to their basal values at the beginning and end of the analysis window. To improve readability, integration limits are not shown in the equations presented in the main text. For readability, the third component is denoted as *G*_max_ − *G*_min_ in the main text. These quantities are identical when glucose has a single rising phase. In the actual calculations, the third component was evaluated in its general form, ∫ *Ġ*_+_*dt*, thereby accounting for additional rises in biphasic or multiphasic glucose trajectories.

### Mathematical model simulating postprandial plasma glucose dynamics

A previously validated meal simulation model of human glucose-insulin regulation^31^ was used to simulate glucose-insulin interactions under varying glucose intake profiles. This physiologically based model consists of modular subsystems representing glucose absorption, endogenous glucose production, insulin secretion and action, and peripheral glucose uptake. Parameters were estimated using data sets from individuals with NGT and T2DM undergoing triple tracer tolerance tests. The detailed formulation has been described previously^31^.

A dataset of 10,000 human individuals was generated by systematically varying parameters across their known physiological ranges. Specifically, insulin sensitivity (Vmx) and first-phase insulin secretion (K) were each assigned 100 evenly spaced values across their respective ranges, and the Cartesian product of these grids (100 × 100) yielded 10,000 unique parameter combinations representing phenotypes from NGT to T2DM^31,58^. Simulations were performed for both bolus ingestion (20, 30, or 40 g glucose ingested within 4 minutes) and continuous ingestion over 1 hour. Using glucose input (*F*), glucose dynamics (***G***), and the equation, *F* = ***K*** · ***G***, the ***K*** vector was estimated for each individual. The estimated ***K*** values were then compared with the product of Vmx and K. Additionally, the temporal distortion of glucose dynamics in ***G*** was compared with glucose autocorrelation-based index (AC_Mean^6^).

### Classification of OGTT glucose-response morphology

Plasma glucose concentrations during the OGTT were measured at 0, 30, 60, 90, and 120 min in the *Kobe University Study*. Glucose-response morphology was classified algorithmically using a prespecified threshold of 20 mg/dL to identify meaningful changes in glucose excursions and to minimize classification driven by minor fluctuations. For each participant, glucose differences between adjacent time points were calculated. An increase of >20 mg/dL was classified as a significant rise, a decrease of >20 mg/dL as a significant fall, and changes within ±20 mg/dL as not significant. A biphasic glucose response was defined as a curve showing: (i) an initial significant rise from fasting glucose to a peak occurring at 30, 60, or 90 min; (ii) a subsequent significant fall from this peak to a nadir before 120 min; and (iii) a significant re-rise from this nadir to the 120-min value. Triphasic responses, defined as curves showing a significant rise, fall, rise, and fall across consecutive OGTT intervals, were classified separately and were not included in the biphasic category. A monophasic glucose response was defined as a curve with a peak between 30 and 90 min, a significant rise from fasting glucose to the peak, no significant re-rise after the peak, and a fall of at least 20 mg/dL from 90 to 120 min. Curves that did not meet any of these definitions were classified as unclassified. The clamp-derived *k*_GluG_⁄*k*_GluQ_ values were then compared between the monophasic and biphasic groups.

### Significance testing of principal component cumulative contribution rates

To assess whether the observed explained variance ratios of the leading principal component(s) exceeded values expected under the null hypothesis of mutually independent input variables, we performed a Monte Carlo null-distribution test for each principal component analysis (PCA). For each test, a null reference distribution of the relevant PCA statistic was constructed by repeating the entire PCA procedure 1,000 times on synthetic datasets having the same shape (number of observations × number of features) as the empirical input matrix. Each synthetic dataset was generated as i.i.d. half-normal samples (the absolute value of a standard normal variate; numpy.random.default_rng with a fixed seed) to mimic the non-negative range of the empirical inputs ***K*** and ***G***/*F*. Each input matrix was z-score normalized column-wise before PCA, identically to the procedure used for the empirical data, and PCAs were performed with scikit-learn 1.0.2.

Two test statistics were used depending on the analysis. (i) For the PCA of ***K*** in the *Kobe University Study*, where PCA was performed at the group level on a matrix of individuals × ***K*** components (3 components: DI_/clep_, DI_/clei_, and DI_/cled_), the test statistic was the contribution rate of the first principal component (PC1). Separate null distributions were generated for the NGT and the T2DM groups using the corresponding empirical matrix dimensions. (ii) For the PCA of ***G***/*F* in the *University of Tokyo Study* and the *San Francisco Bay Area Study1*, where PCA was performed independently for each participant on a within-individual matrix of glucose-input conditions × ***G***/*F* components (6 conditions × 3 components), the test statistic was the cumulative explained variance ratio of PC1 and PC2 averaged across participants (n = 3 for the *University of Tokyo Study*; n = 22 for the *San Francisco Bay Area Study1*). For each Monte Carlo replicate, the same averaging across the corresponding number of synthetic participants was performed before storing the replicate statistic, so that the null distribution properly reflected the participant-level averaging used for the empirical statistic. The one-sided empirical *P* value was computed as *P* = (B + 1)/(N + 1), where N = 1,000 is the number of Monte Carlo replicates and B is the number of replicates yielding a null test statistic greater than or equal to the empirical statistic. Statistical significance was defined as *P* < 0.05.

### Predictive modeling of glucose tolerance and related phenotypes

For logistic regression analysis of glucose intolerance in the *Kobe University Study*, three feature sets were evaluated: DI_/cle_ (DI_/clep_, DI_/clei_, and DI_/cled_), SPINA (SPINA-GR, SPINA-GBeta, and SPINA-DI), and HOMA (HOMA-β, HOMA-IR, and QUICKI). For each feature set, a logistic regression classifier was fitted after z-score standardization. Classification performance was evaluated by repeated stratified 5-fold cross-validation. The same cross-validation splits were used for all feature sets to enable paired comparisons. For each test fold, predicted probabilities were used to calculate receiver operating characteristic (ROC) curves and area under the curve (AUC) values. Pairwise differences in AUC between feature sets were assessed using paired bootstrap resampling of participants with 2,000 bootstrap iterations. Two-sided P values were calculated as twice the smaller tail probability of the bootstrap distribution of AUC differences.

For predictive modeling in the *CGMacros Dataset*, two binary classification tasks were evaluated using the same framework: classification of glucose intolerance and classification of gut microbial diversity status. Glucose intolerance was defined based on HbA1c measured in the blood analytics file, with participants with HbA1c ≥ 6.0% classified as glucose intolerant and those with HbA1c < 6.0% classified as not glucose intolerant. Gut microbial diversity status was defined using the Active Microbial Diversity score from the gut health profile. Participants with an Active Microbial Diversity score of 3, corresponding to the “Good” category, were classified as having high microbial diversity, whereas participants with scores below 3 were classified as not having high microbial diversity.

To evaluate the predictive power of the blood glucose response indices (***G***) on the administered glucose dose (*F*), two linear regression models (*F* = *K* · *G*_AUC_ or *F* = ***K*** · ***G***) were evaluated using data from the *University of Tokyo Study*. For each participant, the ***K*** was assumed to remain constant throughout all observations. The predictive accuracy was evaluated by comparing the predicted *F* with the observed *F*. Using data from the *CGMacros Dataset* and *HPP Cohort* and the *F* = ***K*** · ***G*** equation, individual ***K*** values were estimated from CGM-recorded glucose dynamics and dietary carbohydrate intake. For each annotated meal, CGM values were extracted within a three-hour postprandial window starting at the recorded mealtime. These free-living profiles included real-world complexities, such as periods of exercise, mixed (non-glucose-load) meals, and discrepancies between actual and logged mealtimes. Rather than excluding these sources of variability, analyses were conducted to test whether the simple formulation was informative for predicting IMT and hepatic status despite these challenges. The model parameters (*K* or ***K***) were estimated by non-linear least-squares minimization of the residuals across all available input conditions, with all components constrained to be greater than zero. This estimation was conducted using SciPy 1.10.1^57^.

For predictive modeling in the *Shanghai Study*, two types of CGM-derived representations were compared. First, the summary features were used as a baseline representation, including mean glucose level, standard deviation of glucose, and glucose autocorrelation-based index (AC_Mean). Because standardized glucose-input information (*F*) was not available in the Shanghai Study, ***K*** was not estimated directly. Instead, mean glucose, glucose standard deviation, and AC_Mean were selected as simple glucose-only descriptors of overall exposure, variability, and temporal structure, respectively, motivated by their conceptual correspondence to the components of ***G***. In the simulation analysis, the temporal-distortion component of ***G*** was also significantly associated with AC_Mean (**Extended Data Fig. 5b**). Second, GluFormer embeddings were used as a deep-learning-based representation of CGM dynamics. Because pretrained GluFormer model weights were not publicly available at the time of analysis and unresolved GitHub repository issues limited reproducible extraction of embeddings from additional datasets, we restricted GluFormer-based analyses to the *Shanghai Study*, for which precomputed embeddings were publicly available. The publicly available GluFormer embeddings (https://github.com/Guylu/GluFormer) were matched to participants using patient identifiers and merged with the clinical dataset. For binary outcomes, including diabetic complications, logistic regression models were trained. All input variables were standardized within each training fold, and class imbalance was addressed using balanced class weights. Model performance was evaluated using stratified five-fold cross-validation with a fixed random seed. Receiver operating characteristic curves were generated for each fold, and the area under the curve (AUC) was calculated. For continuous outcomes, including two-hour postprandial plasma glucose levels, ridge regression models were trained using five-fold cross-validation. Input variables were standardized within each training fold. Predictive performance was assessed using the Pearson correlation coefficient between measured and predicted values. Predictions from all test folds were pooled to visualize the association between measured and predicted values.

### Statistical analyses

Group comparisons were performed using a Welch’s *t*-test. Associations between variables were assessed using either Spearman’s correlation coefficients or linear regression models. Multiple comparisons were adjusted using the Benjamini-Hochberg procedure. Confidence intervals were estimated using bootstrap resampling with 10,000 iterations. Statistical significance was defined as *Q* < 0.05 or 95% confidence intervals excluding zero. Prior to the linear regression analyses, all data were standardized using z-score normalization. The Akaike Information Criterion (AIC) was used to compare statistical models containing different numbers of input variables. Because goodness-of-fit measures such as the log-likelihood inevitably improve as variables are added, relying on fit alone tends to favor more complex models and increases the risk of overfitting. The AIC addresses this problem by jointly accounting for model fit and model complexity. These analyses were performed using statsmodels 0.13.2 (https://www.statsmodels.org/stable/index.html).

Hierarchical clustering analyses and principal component analyses were performed after z-score normalization of the input features. Euclidean distance metrics and Ward linkage were used for clustering. The quality of the hierarchical clustering analysis was assessed using silhouette analysis^59^. These analyses were performed using scikit-learn 1.0.2 (https://scikit-learn.org/stable/). Exploratory factor analysis was used to explore the latent structure among correlated measures, following the previously reported approach^60–63^ with some modifications. Variables with absolute factor loadings of at least 0.35 were used for interpretation. Oblimin rotation was used to improve interpretation. To evaluate the applicability of the factor analysis, the Kaiser-Meyer-Olkin and Bartlett’s spherical test were performed. Prior to the analysis, all data were standardized using z-score normalization. These analyses were performed using factor-analyzer 0.5.1 (https://pypi.org/project/factor-analyzer/).

## Data Availability

The data supporting the findings of this study originate from the Kobe University Study25, the Kobe University Study26, the HuMet Repository48, the PREDICT1 Study49, the porcine IVGTT Study50, the University of Tokyo Study32, the San Francisco Bay Area Study133, the San Francisco Bay Area Study216, the CGMacros Dataset36, the Human Phenotype Project (HPP) Cohort18,34,35, and the Shanghai Study38. Data from the Kobe University Study are available at https://journals.plos.org/plosone/article?id=10.1371/journal.pone.0143880#pone.0143880.ref03152. Data from the Kobe University Study2 are available at https://www.nature.com/articles/s43856-025-00819-56. Data from the HuMet Repository are available at https://humet.helmholtz-munich.de/p/app/humet. Data from the PREDICT1 Study are available at https://www.nature.com/articles/s41591-020-0934-049. Data from the porcine IVGTT Study are available at https://journals.plos.org/plosone/article?id=10.1371/journal.pone.0148896. Data from the University of Tokyo Study are available at https://www.nature.com/articles/s41540-019-0108-1#Sec932. Data from the San Francisco Bay Area Study1 are available at https://pmc.ncbi.nlm.nih.gov/articles/PMC6057684/#sec01133. Data from the San Francisco Bay Area Study2 are available at https://www.nature.com/articles/s41551-024-01311-6#Sec1316. Data from the CGMacros Dataset are available at https://physionet.org/content/cgmacros/1.0.0/. The HPP Cohort dataset used in this study is governed by strict privacy and ethical protocols, as approved by the relevant Institutional Review Boards. Due to data protection and participant confidentiality considerations, data access is limited and is granted only for approved research proposals18,34,35. Data from the Shanghai Study are available at https://www.nature.com/articles/s41597-023-01940-738. The code that simulates blood glucose, insulin, and C-peptide levels is available at our GitHub repository (https://github.com/HikaruSugimoto/Glucose_simulation).

## Acknowledgments

We thank Nancy R. Gough (BioSerendipity, LLC) for critical reading and editorial assistance. We thank the Human Phenotype Project (HPP) and Pheno.AI for providing access to the cohort data used in this study. The images in this study are from TogoTV (© 2016 DBCLS TogoTV, CC-BY-4.0) and NIH BioArt (https://bioart.niaid.nih.gov/).

## Funding

Japan Society for the Promotion of Science (JSPS) KAKENHI (JP21H04759) (SK)

CREST, the Japan Science and Technology Agency (JST) (JPMJCR2123) (SK)

The Uehara Memorial Foundation (SK)

The Takeda Science Foundation (HS)

## Data and code availability

The data supporting the findings of this study originate from the *Kobe University Study*^25^, the *Kobe University Study2*^6^, the *HuMet Repository*^48^, the *PREDICT1 Study*^49^, the *porcine IVGTT Study*^50^, the *University of Tokyo Study*^32^, the *San Francisco Bay Area Study1*^33^, the *San Francisco Bay Area Study2*^16^, the *CGMacros Dataset*^36^, the *Human Phenotype Project (HPP) Cohort*^18,34,35^, and the *Shanghai Study*^38^. Data from the *Kobe University Study* are available at https://journals.plos.org/plosone/article?id=10.1371/journal.pone.0143880#pone.0143880.ref031^52^. Data from the *Kobe University Study2* are available at https://www.nature.com/articles/s43856-025-00819-5^6^. Data from the *HuMet Repository* are available at https://humet.helmholtz-munich.de/p/app/humet. Data from the *PREDICT1 Study* are available at https://www.nature.com/articles/s41591-020-0934-0^49^. Data from the *porcine IVGTT Study* are available at https://journals.plos.org/plosone/article?id=10.1371/journal.pone.0148896. Data from the *University of Tokyo Study* are available at https://www.nature.com/articles/s41540-019-0108-1#Sec9^32^. Data from the *San Francisco Bay Area Study1* are available at https://pmc.ncbi.nlm.nih.gov/articles/PMC6057684/#sec011^33^. Data from the *San Francisco Bay Area Study2* are available at https://www.nature.com/articles/s41551-024-01311-6#Sec13^16^. Data from the *CGMacros Dataset* are available at https://physionet.org/content/cgmacros/1.0.0/. The *HPP Cohort* dataset used in this study is governed by strict privacy and ethical protocols, as approved by the relevant Institutional Review Boards. Due to data protection and participant confidentiality considerations, data access is limited and is granted only for approved research proposals^18,34,35^. Data from the *Shanghai Study* are available at https://www.nature.com/articles/s41597-023-01940-7^38^. The code that simulates blood glucose, insulin, and C-peptide levels is available at our GitHub repository (https://github.com/HikaruSugimoto/Glucose_simulation).

## Author contributions

Conceptualization: HS, SK; Methodology: HS; Investigation: HS; Visualization: HS; Funding acquisition: SK; Project administration: WO, SK; Supervision: WO, SK; Writing – original draft: HS; Writing – review & editing: KH, TY, KS, WO, SK

## Supplementary information

Supplementary Notes 1 to 14

Extended Data Figures 1 to 5

Supplementary References

## References

1. Sun, H. et al. IDF Diabetes Atlas: Global, regional and country-level diabetes prevalence estimates for 2021 and projections for 2045. Diabetes Res. Clin. Pract. 183, 109119 (2022).

2. GBD 2021 Diabetes Collaborators. Global, regional, and national burden of diabetes from 1990 to 2021, with projections of prevalence to 2050: a systematic analysis for the Global Burden of Disease Study 2021. Lancet 402, 203–234 (2023).

3. Hulman, A. et al. Glucose patterns during an oral glucose tolerance test and associations with future diabetes, cardiovascular disease and all-cause mortality rate. Diabetologia 61, 101–107 (2018).

4. Su, G. et al. Impact of admission glycemic variability, glucose, and glycosylated hemoglobin on major adverse cardiac events after acute myocardial infarction. Diabetes Care 36, 1026–1032 (2013).

5. Sugimoto, H. et al. Association between continuous glucose monitoring-derived metrics and coronary plaque vulnerability: A retrospective exploratory analysis. eLife, 14:RP102860 (2026).

6. Sugimoto, H. et al. Improved detection of decreased glucose handling capacities via continuous glucose monitoring-derived indices. Commun. Med. (Lond.) 5, 103 (2025).

7. Mita, T. et al. Continuous glucose monitoring-derived time in range and CV are associated with altered tissue characteristics of the carotid artery wall in people with type 2 diabetes. Diabetologia 66, 2356–2367 (2023).

8. Zhong, H. et al. Two-week continuous glucose monitoring-derived metrics and degree of hepatic steatosis: a cross-sectional study among Chinese middle-aged and elderly participants. Cardiovasc. Diabetol. 23, 322 (2024).

9. Sugimoto, H., Sapir, G., Keshet, A. & Kuroda, S. Use of continuous glucose monitoring to stratify individuals without diabetes. Commun. Med. (Lond.) 6 (2026).

10. DeFronzo, R. A., Tobin, J. D. & Andres, R. Glucose clamp technique: a method for quantifying insulin secretion and resistance. Am. J. Physiol. 237, E214–23 (1979).

11. Ader, M., Pacini, G., Yang, Y. J. & Bergman, R. N. Importance of glucose per se to intravenous glucose tolerance. Comparison of the minimal-model prediction with direct measurements. Diabetes 34, 1092–1103 (1985).

12. Steil, G. M. et al. Evaluation of insulin sensitivity and beta-cell function indexes obtained from minimal model analysis of a meal tolerance test. Diabetes 53, 1201–1207 (2004).

13. Cobelli, C. et al. The oral minimal model method. Diabetes 63, 1203–1213 (2014).

14. Ajmera, I., Swat, M., Laibe, C., Le Novère, N. & Chelliah, V. The impact of mathematical modeling on the understanding of diabetes and related complications. CPT Pharmacometrics Syst Pharmacol 2, e54 (2013).

15. Kahn, S. E. et al. The β Cell in Diabetes: Integrating Biomarkers With Functional Measures. Endocr. Rev. 42, 528–583 (2021).

16. Metwally, A. A. et al. Prediction of metabolic subphenotypes of type 2 diabetes via continuous glucose monitoring and machine learning. *Nat*. Biomed. Eng. 1–18 (2024).

17. Lu, Y. et al. A pretrained transformer model for decoding individual glucose dynamics from continuous glucose monitoring data. Natl. Sci. Rev. 12, nwaf039 (2025).

18. Reicher, L. et al. Deep phenotyping of health-disease continuum in the Human Phenotype Project. Nat. Med. 1–13 (2025).

19. Carletti, M. et al. Multimodal AI correlates of glucose spikes in people with normal glucose regulation, pre-diabetes and type 2 diabetes. Nat. Med., 1–7 (2025).

20. Kodama, K. et al. Ethnic differences in the relationship between insulin sensitivity and insulin response: a systematic review and meta-analysis. Diabetes Care 36, 1789–1796 (2013).

21. Karin, O., Swisa, A., Glaser, B., Dor, Y. & Alon, U. Dynamical compensation in physiological circuits. Mol. Syst. Biol. 12, 886 (2016).

22. Kahn, S. E. et al. Quantification of the relationship between insulin sensitivity and beta-cell function in human subjects. Evidence for a hyperbolic function. Diabetes 42, 1663–1672 (1993).

23. Sergazinov, R., et al. GlucoBench: Curated list of continuous glucose monitoring datasets with prediction benchmarks. *arXiv [q-bio.QM]* (2024).

24. Aoki, S. et al. A universal biomolecular integral feedback controller for robust perfect adaptation. Nature 570, 533–537 (2019).

25. Okuno, Y. et al. Postprandial serum C-peptide to plasma glucose concentration ratio correlates with oral glucose tolerance test-and glucose clamp-based disposition indexes. Metabolism 62, 1470–1476 (2013).

26. Watson, E. M., Chappell, M. J., Ducrozet, F., Poucher, S. M. & Yates, J. W. T. A new general glucose homeostatic model using a proportional-integral-derivative controller. Comput. Methods Programs Biomed. 102, 119–129 (2011).

27. van Veen, L., Morra, J., Palanica, A. & Fossat, Y. Homeostasis as a proportional–integral control system. npj Digital Medicine 3, 1–7 (2020).

28. Bocci, F., Zhou, P. & Nie, Q. spliceJAC: transition genes and state-specific gene regulation from single-cell transcriptome data. Mol. Syst. Biol. 18, e11176 (2022).

29. Gibson, G. Decanalization and the origin of complex disease. Nat. Rev. Genet. 10, 134–140 (2009).

30. Mingrone, G. et al. Insulin sensitivity depends on the route of glucose administration. Diabetologia 63, 1382–1395 (2020).

31. Dalla Man, C., Rizza, R. A. & Cobelli, C. Meal simulation model of the glucose-insulin system. IEEE Trans. Biomed. Eng. 54, 1740–1749 (2007).

32. Fujii, M. et al. Logical design of oral glucose ingestion pattern minimizing blood glucose in humans. NPJ Syst Biol Appl 5, 31 (2019).

33. Hall, H. et al. Glucotypes reveal new patterns of glucose dysregulation. PLoS Biol. 16, e2005143 (2018).

34. Keshet, A. et al. CGMap: Characterizing continuous glucose monitor data in thousands of non-diabetic individuals. Cell Metab. 35, 758–769.e3 (2023).

35. Shilo, S. et al. 10 K: a large-scale prospective longitudinal study in Israel. Eur. J. Epidemiol. 36, 1187–1194 (2021).

36. Das, A. et al. CGMacros: a pilot scientific dataset for personalized nutrition and diet monitoring. Sci. Data 12, 1557 (2025).

37. Le Chatelier, E. et al. Richness of human gut microbiome correlates with metabolic markers. Nature 500, 541–546 (2013).

38. Zhao, Q. et al. Chinese diabetes datasets for data-driven machine learning. Sci Data 10, 35 (2023).

39. Lutsker, G. et al. A foundation model for continuous glucose monitoring data. Nature 650, 978–986 (2026).

40. Bergman, R. N., Ader, M., Huecking, K. & Van Citters, G. Accurate assessment of beta-cell function: the hyperbolic correction. Diabetes 51 Suppl 1, S212–20 (2002).

41. Bergman, R. N., Phillips, L. S. & Cobelli, C. Physiologic evaluation of factors controlling glucose tolerance in man: measurement of insulin sensitivity and beta-cell glucose sensitivity from the response to intravenous glucose. J. Clin. Invest. 68, 1456–1467 (12 1981).

42. Lorenzo, C. et al. Relationship of insulin sensitivity, insulin secretion, and adiposity with insulin clearance in a multiethnic population: the insulin Resistance Atherosclerosis study. Diabetes Care 36, 101–103 (2013).

43. Goodarzi, M. O. et al. Insulin sensitivity and insulin clearance are heritable and have strong genetic correlation in Mexican Americans. Obesity 22, 1157–1164 (2014).

44. Ahlqvist, E. et al. Novel subgroups of adult-onset diabetes and their association with outcomes: a data-driven cluster analysis of six variables. Lancet Diabetes Endocrinol. 6, 361–369 (2018).

45. Lutsker, G., et al. From glucose patterns to health outcomes: A generalizable foundation model for continuous glucose monitor data analysis. *arXiv [q-bio.QM]* (2024).

46. Lorenzo, C. et al. Cross-sectional and longitudinal changes of glucose effectiveness in relation to glucose tolerance: the insulin resistance atherosclerosis study. Diabetes Care 34, 1959–1964 (2011).

47. Bergman, R. N. Origins and History of the Minimal Model of Glucose Regulation. Front. Endocrinol. 11, 583016 (2020).

48. Weinisch, P. et al. The HuMet Repository: Watching human metabolism at work. *bioRxiv* 10.1101/2023.08.08.550079 (2023) doi:10.1101/2023.08.08.550079.

49. Berry, S. E. et al. Human postprandial responses to food and potential for precision nutrition. Nat. Med. 26, 964–973 (2020).

50. Manell, E., Hedenqvist, P., Svensson, A. & Jensen-Waern, M. Establishment of a refined oral glucose tolerance test in pigs, and assessment of insulin, glucagon and glucagon-like peptide-1 responses. PLoS One 11, e0148896 (2016).

51. Sugimoto, H. et al. DI/cle, a Measure Consisting of Insulin Sensitivity, Secretion, and Clearance, Captures Diabetic States. J. Clin. Endocrinol. Metab. 10.1210/clinem/dgad392 (2023) doi:10.1210/clinem/dgad392.

52. Ohashi, K. et al. Glucose Homeostatic Law: Insulin Clearance Predicts the Progression of Glucose Intolerance in Humans. PLoS One 10, e0143880 (2015).

53. Matsuda, M. & DeFronzo, R. A. Insulin sensitivity indices obtained from oral glucose tolerance testing: comparison with the euglycemic insulin clamp. Diabetes Care 22, 1462– 1470 (1999).

54. Dietrich, J. W. et al. SPINA Carb: a simple mathematical model supporting fast in-vivo estimation of insulin sensitivity and beta cell function. Sci. Rep. 12, 17659 (2022).

55. Dietrich, J. W. et al. A novel simple disposition index (SPINA-DI) from fasting insulin and glucose concentration as a robust measure of carbohydrate homeostasis. J. Diabetes 16, e13525 (2024).

56. Yamada, T., et al. Mathematical models of the effect of glucagon on glycemia in individuals with type 2 diabetes treated with dapagliflozin. J. Endocr. Soc. 8, bvae067 (2024).

57. Virtanen, P. et al. SciPy 1.0: fundamental algorithms for scientific computing in Python. Nat. Methods 17, 261–272 (2020).

58. Basu, R. et al. Effects of age and sex on postprandial glucose metabolism: differences in glucose turnover, insulin secretion, insulin action, and hepatic insulin extraction. Diabetes 55, 2001–2014 (2006).

59. Rousseeuw, P. J. Silhouettes: A graphical aid to the interpretation and validation of cluster analysis. J. Comput. Appl. Math. 20, 53–65 (1987).

60. Cappelleri, J. C., Gerber, R. A., Kourides, I. A. & Gelfand, R. A. Development and factor analysis of a questionnaire to measure patient satisfaction with injected and inhaled insulin for type 1 diabetes. Diabetes Care 23, 1799–1803 (2000).

61. Lakka, H.-M. et al. The metabolic syndrome and total and cardiovascular disease mortality in middle-aged men. JAMA 288, 2709–2716 (2002).

62. Oh, J.-Y., Hong, Y. S., Sung, Y.-A. & Barrett-Connor, E. Prevalence and factor analysis of metabolic syndrome in an urban Korean population. Diabetes Care 27, 2027–2032 (2004).

63. Guo, W., Zhou, Q., Jia, Y. & Xu, J. Cluster and Factor Analysis of Elements in Serum and Urine of Diabetic Patients with Peripheral Neuropathy and Healthy People. Biol. Trace Elem. Res. 194, 48–57 (2020).

