## Supplementary file for "A law of human glucose homeostasis for inferring glucose regulatory capacity from glucose time series"

### Supplementary Note

1. Estimation of insulin secretion and clearance from hyperglycemic and hyperinsulinemic-euglycemic clamp tests

We developed a mathematical model for estimating insulin secretion and insulin clearance from paired hyperglycemic and hyperinsulinemic-euglycemic clamp data. During the hyperglycemic clamp, plasma glucose approached a steady state by 60 min and remained stable at 75 and 90 min (**Fig. 1d**). In contrast, the insulin/glucose and C-peptide/glucose ratios continued to rise (**Extended Data Fig. 1a, b**), and both insulin and C-peptide concentrations significantly increased with the initial rise in glucose (**Extended Data Fig. 1c, d**). To model these observations, we first formulated insulin and C-peptide kinetics using the following expressions:

$$\dot{I} = k_{\mathrm{r}} k_{\mathrm{Sp}} G + k_{\mathrm{r}} k_{\mathrm{Si}} (G - G_{\mathrm{b}})_{+} t + k_{\mathrm{r}} k_{\mathrm{Sd}} \dot{G}_{+} - k_{\mathrm{Ic}} I + f_{\mathrm{I}}, \qquad \text{Eq. (1)}$$

$$\dot{C} = k_{\mathrm{Sp}} G + k_{\mathrm{Si}} (G - G_{\mathrm{b}})_{+} t + k_{\mathrm{Sd}} \dot{G}_{+} - k_{\mathrm{Cc}} C. \qquad \text{Eq. (2)}$$

Here, $k_{\mathrm{Sp}}$, $k_{\mathrm{Si}}$, and $k_{\mathrm{Sd}}$ represent the static, potentiated, and first-phase components of insulin secretion, respectively. Parameters $1 - k_{\mathrm{r}}$, $k_{\mathrm{Ic}}$, $k_{\mathrm{Cc}}$, and $f_{\mathrm{I}}$ represent hepatic insulin clearance, peripheral insulin clearance, C-peptide clearance, and insulin infusion rate, respectively. $G_{\mathrm{b}}$ is the basal glucose concentration. The first-phase term uses $\dot{G}_{+}$, which equals $\dot{G}$when glucose is rising and zero otherwise, and the potentiation term is evaluated as $k_{\mathrm{Si}} \cdot \max(G - G_{\mathrm{b}}, 0) \cdot t$, so that it contributes only while glucose exceeds its basal value. Both terms are therefore non-negative at all times, and neither can drive insulin secretion below its basal rate, including during the hyperinsulinaemic–euglycaemic phase, in which plasma glucose was clamped at the fasting concentration, and during any post-prandial undershoot. These three components of insulin secretion ($k_{\mathrm{Sp}}$, $k_{\mathrm{Si}}$, and $k_{\mathrm{Sd}}$) align with the previous studies indicating that insulin secretion has three components: static component, dynamic component, and potentiation factor[1]. They are also conceptually similar to a proportional-integral-derivative (PID) control structure[2].

The PID-like structure was not introduced solely by analogy to engineering control systems. Instead, it was used as a minimal phenomenological representation of the insulin and C-peptide dynamics observed experimentally. During the hyperglycemic clamp, plasma glucose reached a near steady state, whereas insulin/glucose and C-peptide/glucose ratios continued to increase, indicating that insulin secretion could not be explained by a component proportional only to the current glucose level. In addition, the rapid increase in insulin and C-peptide during the initial rise in glucose required a component sensitive to increasing glucose. We therefore represented insulin secretion using three components: a glucose-proportional component, a potentiation component during sustained hyperglycemia, and a first-phase component associated with the rate of glucose increase. These components correspond conceptually to the proportional, integral-like, and derivative-like elements of a PID controller and are consistent with previous descriptions of insulin secretion as comprising static, potentiation, and dynamic components. Thus, the PID-like formulation provides an empirically motivated structure, rather than an ad hoc assumption. It is also important to note that the values of the parameters in the above equations are not solely determined by pancreatic insulin secretion or hepatic insulin clearance. They are also influenced by the distribution volumes of insulin and C-peptide throughout the body. Although it is possible to estimate and correct for these distribution volumes using physiological indices such as body weight[3,4], we do not apply such corrections here. This is both for simplicity and because such estimation of distribution volume involves some degree of uncertainty and introduces error. Consequently, the parameters in the above equations are defined to include individual differences in distribution volumes.

Assuming steady-state conditions ($\dot{I} = 0$, $\dot{C} = 0$) at baseline (0 min), Eqs. (1) and (2) were simplified as follows:

$$0 = k_{\mathrm{r}} k_{\mathrm{Sp}} G_0 - k_{\mathrm{Ic}} I_0, \qquad \text{Eq. (3)}$$

$$0 = k_{\mathrm{Sp}} G_0 - k_{\mathrm{Cc}} C_0. \qquad \text{Eq. (4)}$$

The subscripts denote the time (min) during a hyperglycemic and a hyperinsulinemic-euglycemic clamp test. At 0 min, we approximated $k_{\mathrm{r}} k_{\mathrm{Sp}} G = k_{\mathrm{Ic}} I$ and $k_{\mathrm{Sp}} G = k_{\mathrm{Cc}} C$, which reduces Eqs. (1) and (2) to:

$$\dot{I} = k_{\mathrm{r}} k_{\mathrm{Sd}} \dot{G}_+, \qquad \text{Eq. (5)}$$

$$\dot{C} = k_{\mathrm{Sd}} \dot{G}_+. \qquad \text{Eq. (6)}$$

During the first 0-5 min of the clamp test, Eqs. (5) and (6) were approximated as follows:

$$\frac{I_5 - I_0}{5} = k_{\mathrm{r}} k_{\mathrm{Sd}} \frac{G_5 - G_0}{5}, \qquad \text{Eq. (7)}$$

$$\frac{C_5 - C_0}{5} = k_{\mathrm{Sd}} \frac{G_5 - G_0}{5}. \qquad \text{Eq. (8)}$$

By approximating the first- and second-order derivatives as zero ($\dot{G} = 0$, $\ddot{C} = 0$) between 60–90 min during the clamp tests (**Fig. 1d**), Eq. (2) can be written as follows:

$$0 = k_{\mathrm{Si}}(G - G_{\mathrm{b}}) - k_{\mathrm{Cc}} \dot{C}. \qquad \text{Eq. (9)}$$

Eq. (9) was approximated as follows:

$$\frac{k_{\mathrm{Si}}}{k_{\mathrm{Cc}}} = \frac{1}{2}\left(\frac{C_{75} - C_{60}}{15 \times (G_{60} - G_0)} + \frac{C_{90} - C_{75}}{15 \times (G_{75} - G_0)}\right). \qquad \text{Eq. (10)}$$

By approximating the first-order derivative as zero and $k_{\mathrm{r}} k_{\mathrm{Si}}(G - G_{\mathrm{b}})t = 0$ at 190–220 min during the clamp tests, Eqs. (1) and (2) can be written as follows:

$$0 = k_{\mathrm{r}} k_{\mathrm{Sp}} G - k_{\mathrm{Ic}} I + f_{\mathrm{I}}, \qquad \text{Eq. (11)}$$

$$0 = k_{\mathrm{Sp}} G - k_{\mathrm{Cc}} C. \qquad \text{Eq. (12)}$$

Eqs. (3), (4), (11), and (12) can be transformed as follows:

$$k_{\mathrm{Ic}} = \frac{1}{2}\left(\frac{f_{190}}{I_{190} - I_0 \frac{C_{190}}{C_0}} + \frac{f_{220}}{I_{220} - I_0 \frac{C_{220}}{C_0}}\right). \qquad \text{Eq. (13)}$$

Consequently, the parameters of Eqs. (1) and (2) were estimated as follows:

$$k_{\mathrm{Sp}} = \frac{k_{\mathrm{Ic}} I_0}{k_{\mathrm{r}} G_0};\ k_{\mathrm{Si}} = \frac{k_{\mathrm{Cc}}}{2}\left(\frac{C_{75} - C_{60}}{15 \times (G_{60} - G_0)} + \frac{C_{90} - C_{75}}{15 \times (G_{75} - G_0)}\right);\ k_{\mathrm{Sd}} = \frac{C_5 - C_0}{G_5 - G_0};$$

$$k_{\mathrm{r}} = \frac{I_5 - I_0}{C_5 - C_0};\ k_{\mathrm{Ic}} = \frac{1}{2}\left(\frac{f_{190}}{I_{190} - I_0 \frac{C_{190}}{C_0}} + \frac{f_{220}}{I_{220} - I_0 \frac{C_{220}}{C_0}}\right);\ k_{\mathrm{Cc}} = \frac{k_{\mathrm{Ic}} I_0}{k_{\mathrm{r}} C_0}.$$

It should be noted that if the glucose level remains above $G_{\mathrm{b}}$ indefinitely, $k_{\mathrm{Si}}(G - G_{\mathrm{b}})t$ becomes infinite, which is physiologically unrealistic. However, in this study, $\frac{C_{75} - C_{60}}{15 \times (G_{60} - G_0)}$ and $\frac{C_{90} - C_{75}}{15 \times (G_{75} - G_0)}$ were almost identical (**Extended Data Fig. 1e**), suggesting that the linear approximation is acceptable in this timescale. Furthermore, as glucose quickly reaches steady-state levels during a hyperglycemic clamp, $k_{\mathrm{Si}} \int (G - G_{\mathrm{b}})\, dt$ can be approximated by $k_{\mathrm{Si}}(G - G_{\mathrm{b}})t$. Accordingly, we used $k_{\mathrm{Si}}(G - G_{\mathrm{b}})t$ for analytical parameter estimation in the clamp tests.

We then compared Eqs. (1) and (2) with alternative models for C-peptide kinetics. The kinetics of C-peptide have traditionally been described using a two-compartment model[5,6], which is defined as follows:

$$\dot{C} = k_{\mathrm{Sp}}G + k_{\mathrm{Si}}(G - G_{\mathrm{b}})_{+}t + k_{\mathrm{Sd}}\dot{G}_{+} - k_{\mathrm{Cc}}C - k_{\mathrm{CY}}C + k_{\mathrm{YC}}Y, \qquad \text{Eq. (14)}$$

$$\dot{Y} = k_{\mathrm{CY}}C - k_{\mathrm{YC}}Y, \qquad \text{Eq. (15)}$$

where $C$ and $Y$ represent the C-peptide concentrations in accessible and non-accessible compartments, respectively. The parameters $k_{\mathrm{CY}}$ and $k_{\mathrm{YC}}$ are rate parameters describing C-peptide exchange kinetics. According to the previous study [6], Eqs (14) and (15) can be transformed as follows:

$$k_{\mathrm{Sp}}G + k_{\mathrm{Si}}(G - G_{\mathrm{b}})_{+}t + k_{\mathrm{Sd}}\dot{G}_{+} = -k_{\mathrm{CY}}C_0 e^{-k_{\mathrm{YC}}t} - k_{\mathrm{CY}}k_{\mathrm{YC}} \int_0^t C(s) e^{-k_{\mathrm{YC}}(t-s)} ds + \dot{C} + (k_{\mathrm{CY}} + k_{\mathrm{Cc}})C. \qquad \text{Eq. (16)}$$

The previous work[6] has shown that $k_{\mathrm{CY}}$, $k_{\mathrm{YC}}$ and $k_{\mathrm{Cc}}$ can be estimated from physiological variables such as obesity and age without the need for glucose or C-peptide concentration data. These estimates have been shown to accurately simulate C-peptide dynamics. However, this two-compartment model includes six parameters, and in our study C-peptide data were available at the following time points: 0, 5, 10, 15, 60, 75, 90, 100, 190 and 220 min. Of these, the measurements at 0, 190 and 220 minutes were almost identical, and the value at 75 minutes was well approximated by the values at 60 and 90 minutes. Even with this redundancy, data were available from at least six different informative time points. The number of informative time points was not smaller than the number of fitted parameters. Moreover, in the previous study[6], the correlation between the long half-life of C-peptide estimated from the actual measurements and that estimated from individual characteristics using this method was relatively low ($R$ = 0.28).

We therefore compared the mean absolute error (MAE) of the following three models:
One: a model simulated using Eqs. (1) and (2),
Two (previous): a two-compartment model simulated using Eqs. (14) and (15) with parameters derived from the previous study[6], and
Two (estimate): a two-compartment model with parameters fitted directly to our clamp test data.

Given that equations describing insulin dynamics in the one-compartment model and the two-compartment model are the same, we only compared MAE of C-peptide dynamics. The Two (estimate) model had a significantly lower MAE than the other two models (**Extended Data Fig. 1f**), suggesting that fitting the two-compartment model directly to experimental clamp data provides a more accurate representation of C-peptide kinetics than relying on either the simpler single-compartment model or parameter estimates derived from external studies. Consequently, the two-compartment model was adopted for subsequent simulations and analyses in this study.

Since hepatic insulin clearance has been reported to be suppressed under hyperglycemic conditions[7], we next investigated the temporal dynamics of hepatic insulin clearance during the clamp tests. Specifically, we analyzed how hepatic insulin clearance changed over time. By combining Eq. (1) and Eq. (16), we derived an expression for the hepatic insulin clearance rate constant $k_\mathrm{r}$ during the first 100 minutes, as follows:

$$k_\mathrm{r} = \frac{\dot{I} + k_\mathrm{Ic} I}{-k_\mathrm{CY} C_0 e^{-k_\mathrm{YC} t} - k_\mathrm{CY} k_\mathrm{YC} \int_0^t C(s) e^{-k_\mathrm{YC}(t-s)} ds + \dot{C} + (k_\mathrm{CY} + k_\mathrm{Cc}) C}. \quad \text{Eq. (17)}$$

This formulation allowed the estimation of $k_\mathrm{r}$ at different time points during the clamp experiment. Although both plasma glucose and serum insulin levels were elevated during the hyperglycemic clamp, we found that hepatic insulin clearance, represented by $1 - k_\mathrm{r}$, calculated at 60 and 75 minutes was not significantly different from that obtained at 0 minutes (**Extended Data Fig. 1g**). Therefore, to simplify the model and reduce parameter complexity, we assumed that $k_\mathrm{r}$ remained constant throughout the analysis in this study.

We then investigated the time delay in insulin secretion. By approximating the second-order derivative as zero during the hyperglycemic clamp test, Eqs. (1), (14) and (15) can be written as follows:

$$0 = k_\mathrm{r} k_\mathrm{Sp} \dot{G} + k_\mathrm{r} k_\mathrm{Si} (G - G_\mathrm{b}) - k_\mathrm{Ic} \dot{I}, \quad \text{Eq. (18)}$$

$$0 = k_\mathrm{Sp} \dot{G} + k_\mathrm{Si} (G - G_\mathrm{b}) - k_\mathrm{Cc} \dot{C}. \quad \text{Eq. (19)}$$

At 90 minutes during the clamp tests, Eqs. (18) and (19) can be written as follows:

$$0 = k_\mathrm{r} k_\mathrm{Sp} \dot{G}_{90-} + k_\mathrm{r} k_\mathrm{Si} (G_{90} - G_\mathrm{b}) - k_\mathrm{Ic} \dot{I}_{90-}, \quad \text{Eq. (20)}$$

$$0 = k_\mathrm{Sp} \dot{G}_{90-} + k_\mathrm{Si} (G_{90} - G_\mathrm{b}) - k_\mathrm{Cc} \dot{C}_{90-}, \quad \text{Eq. (21)}$$

$$0 = k_\mathrm{r} k_\mathrm{Sp} \dot{G}_{90+} + k_\mathrm{r} k_\mathrm{Si} (G_{90} - G_\mathrm{b}) - k_\mathrm{Ic} \dot{I}_{90+}, \quad \text{Eq. (22)}$$

$$0 = k_\mathrm{Sp} \dot{G}_{90+} + k_\mathrm{Si} (G_{90} - G_\mathrm{b}) - k_\mathrm{Cc} \dot{C}_{90+}, \quad \text{Eq. (23)}$$

By approximating $\dot{G}_{90-} = 0$, Eqs. (20), (21), (22) and (23) can be written as follows:

$$0 = k_\mathrm{r} k_\mathrm{Sp} \dot{G}_{90+} + k_\mathrm{Ic} \dot{I}_{90-} - k_\mathrm{Ic} \dot{I}_{90+}, \quad \text{Eq. (24)}$$

$$0 = k_\mathrm{Sp} \dot{G}_{90+} + k_\mathrm{Cc} \dot{C}_{90-} - k_\mathrm{Cc} \dot{C}_{90+}. \quad \text{Eq. (25)}$$

We can expect that the more time delay there is in insulin secretion, the smaller the right-hand sides of these equations would be. The values (denoted as Res) were not significantly smaller than 0 (**Extended Data Fig. 1h**). Although it is possible that there is a delay in insulin secretion, the delay did not appear significant in data examined with 10-minute intervals or greater. Of note, a previous study indicated that the length of the past duration whose plasma glucose levels influence the current insulin secretion was approximately 14.9 minutes[8], which also indicates that the effect of time delay in insulin secretion can be considered small in the present data, which were examined at more than 5-15 minute intervals.

Moreover, we show that $k_\mathrm{Si}(G - G_\mathrm{b})t$ in Eq. (14) quantitatively corresponds to the time delay employed in previous studies[9,10]. In the previous studies, insulin secretion rate (ISR) was written as follows:

$$\mathrm{ISR} = \mathrm{ISR}_\mathrm{b}(t) + \phi_\mathrm{d} \dot{G}_+ + \mathrm{ISR}_\mathrm{s}(t), \quad \text{Eq. (26)}$$

$$\dot{\mathrm{ISR}}_\mathrm{s}(t) = -\alpha(\mathrm{ISR}_\mathrm{s}(t) - \beta(G(t) - h)), \quad \text{Eq. (27)}$$

where $\mathrm{ISR}_\mathrm{b}$ is basal insulin secretion, and α ($\mathrm{min}^{-1}$) is a rate constant governing the time delay. When G is constant during clamp tests, the $\mathrm{ISR}_\mathrm{s}$ can be written as follows:

$$\mathrm{ISR}_\mathrm{s}(t) = \beta(G(t) - h) - \beta(G(t) - h)e^{-\alpha t} = \alpha\beta(G(t) - h)t + O(t^2). \quad \text{Eq. (28)}$$

When G increases linearly, as in the initial minutes of hyperglycemic clamp tests ($G(t) = \mathrm{G}_\mathrm{s} t + h$), $\mathrm{ISR}_\mathrm{s}$ can be written as follows:

$$\mathrm{ISR_s}(t) = -\frac{\beta}{\alpha}\mathrm{G_s} + \beta \mathrm{G_s} t + \frac{\beta}{\alpha}\mathrm{G_s} e^{-\alpha t} = \frac{\alpha\beta}{2}(G(t) - h)t + O(t^3). \qquad \text{Eq. (29)}$$

$k_{\mathrm{Si}}(G - G_{\mathrm{b}})t$ in Eq. (14) can be considered to include a rate constant reflecting time delay. Collectively, we mainly investigated Eqs. (1), (14) and (15) in this study. Although the manipulations from Eq. (3) to Eq. (29) involve several approximations that are not strictly rigorous, they are provided to show parameter identifiability and intuitive model behavior. In practice, we estimated the parameters by fitting Eqs. (1), (14), and (15) to the experimental data using a meta-evolutionary programming algorithm combined with nonlinear least-squares optimization. The resulting residuals were small (**Fig. 1e**), indicating the agreement with the data. Therefore, these heuristic approximations do not affect the main analysis or its conclusions.

When plasma glucose remains above the basal level $G_b$, a pure integral formulation, $\int (G - G_{\mathrm{b}})dt$, continues to accumulate with time and can therefore generate a progressively increasing and ultimately divergent secretion component. Such behavior is physiologically unrealistic over sufficiently long timescales. In addition, the accumulated integral does not automatically decay when glucose returns to $G_{\mathrm{b}}$; consequently, an insulin-secretion component driven solely by the integral may remain above baseline even after the glucose stimulus has returned to baseline. A physiologically complete integral-based formulation would therefore require an additional elimination, decay, or saturation mechanism.

One possible alternative is a first-order dynamic formulation such as $\dot{Y} = -\alpha\{Y - \beta(G - h)\}$, so that $Y(t)$ approaches a glucose-dependent steady state rather than accumulating indefinitely and can decay after removal of the glucose stimulus. However, introducing such a formulation requires estimation of at least one additional kinetic parameter, such as the decay constant $\alpha$. Reliable identification of this parameter would require data that adequately characterize the relevant decay timescale. The datasets analyzed in the present study were not designed specifically to identify this additional timescale; therefore, adding a decay parameter would increase model complexity without clear evidence that it could be robustly estimated. For these reasons, we used $k_{\mathrm{Si}}(G - G_{\mathrm{b}})t$ rather than a pure integral term. Importantly, this expression is not assumed to be mathematically equivalent to $\int (G - G_{\mathrm{b}})dt$ for arbitrary glucose trajectories. Instead, it is used as a parsimonious finite-time approximation of the time-dependent potentiation associated with elevated plasma glucose. This formulation has a practical advantage over the pure integral form: when $G$ returns to $G_{\mathrm{b}}$, $k_{\mathrm{Si}}(G - G_{\mathrm{b}})t$ also becomes zero, allowing the corresponding insulin-secretion component to return to baseline without introducing an additional elimination parameter. Moreover, as shown below, this formulation already provided a good empirical fit to the experimental data. It also preserves the intended relationship to the integral representation of cumulative glucose-dependent potentiation and maintains conceptual continuity with previously proposed formulations. We therefore selected $k_{\mathrm{Si}}(G - G_{\mathrm{b}})t$ as a practical compromise among return-to-baseline behavior, empirical adequacy, model parsimony, and correspondence with both the integral-like interpretation and previous modeling approaches.

We refer to this term as integral-like rather than as a true integral. The relationship between the two formulations can be stated precisely. By the definition of the running time average, the cumulative glucose signal can be written as $\int_0^t (G - G_b)\, ds = (\overline{G}(t) - G_b)t$, where $\overline{G}(t)$ is the running time average of glucose over [0, $t$]. Thus, the cumulative integral and the present formulation, $(G - G_b)t$, share the same basic structure, glucose elevation multiplied by elapsed time, and differ only in whether glucose elevation is represented by its running average or by its instantaneous value. The two expressions are identical when glucose is maintained at a constant elevated level, as in the hyperglycemic clamp experiments from which the secretion parameters were estimated, and differ only when the glucose trajectory is transient.

This approximation is particularly appropriate for the relatively short experimental windows examined here. The hyperglycemic and hyperinsulinemic-euglycemic clamp experiments covered 0–220 min, with plasma glucose reaching a near-steady level within approximately 30 min. OGTT analyses covered 0–120 or 0–180 min, during which glucose typically peaked within 30–60 min and returned toward baseline by 120–180 min. Mixed-meal tolerance tests and CGM-based postprandial analyses extended to 240 min and showed comparable temporal profiles. Within these bounded observation windows, the cumulative effect represented by an integral remains finite, while the instantaneous term $(G - G_b)t$ was empirically well behaved and naturally vanished as glucose returned to baseline. Thus, over the experimentally relevant timescales considered here, this formulation provides a parsimonious and empirically adequate representation of the time-dependent potentiation of insulin secretion during elevated plasma glucose, while avoiding the residual effect inherent to a pure integral term.

Two consequences of this formulation should nevertheless be made explicit. First, the derived feature $G_{\mathrm{Distortion}} = \int \max(G - G_{\mathrm{b}}, 0)t\,dt$ weights glucose elevation occurring later in the observation window more strongly than elevation occurring earlier. In contrast, if a true cumulative integral were itself integrated over the same window, earlier glucose elevation would receive greater weight because its effect would persist for a longer period. $G_{\mathrm{Distortion}}$ therefore, reflects the temporal placement of glucose exposure rather than simply its cumulative magnitude. Equivalently, $\frac{G_{Distortion}}{G_{AUC}}$ represents the temporal center of mass of the glucose excursion. Second, because $t$ is defined relative to the onset of the glucose challenge, the term is not invariant to translation of the time origin. The analysis window must therefore be anchored to the start of glucose administration. An error $\delta$ in the recorded start time propagates to an error of approximately $\delta G_{\mathrm{AUC}}$ in $G_{\mathrm{Distortion}}$, which is particularly relevant to free-living data in which meal timing is self-reported. We therefore retained this formulation because of its simplicity and empirical adequacy within the timescales studied here. It should not, however, be interpreted as a physiologically unbounded process or as a general replacement for an integral model over arbitrary timescales. For sustained chronic hyperglycemia over multi-hour to multi-day periods, a model incorporating explicit decay or saturation would be required, together with re-estimation of the corresponding kinetic parameters using data that adequately capture those longer-term dynamics.

We then provide a coarse-grained physiological interpretation of the proportional, integral, and derivative-like terms in the insulin secretion equation ($\mathrm{k_{Sp}G} + \mathrm{k_{Si}}(G - G_{\mathrm{b}})t + \mathrm{k_{Sd}}\dot{\mathrm{G}}_{+}$). The proportional term ($\mathrm{k_{Sp}G}$) represents the local static gain of glucose-stimulated insulin secretion near the operating range of the experiment. Mechanistically, glucose affects secretion through glucose uptake and metabolism, changes in ATP/ADP, closure of ATP-sensitive potassium channels, membrane depolarization, voltage-gated calcium entry, calcium microdomain signaling near exocytotic sites, and fusion of insulin granules[11–15]. Glucose also generates amplifying signals that enhance the efficacy of calcium on exocytosis without necessarily causing a further rise in cytosolic calcium. Detailed beta-cell models explicitly represent parts of this cascade, including glycolysis, ATP/ADP dynamics, calcium handling, and exocytotic granule-pool transitions[13–16]. A compact way to express this is to assume a nonlinear steady or quasi-steady secretion function: $\mathrm{S_{static}} = \mathrm{S_{static}}(\mathrm{G}, \mathrm{N})$, where N denotes slowly varying metabolic, granule-pool, amplifying, incretin, and paracrine states. Around the basal or local operating point, this function can be linearized: $\mathrm{S_{static}}(\mathrm{G}, \mathrm{N}) \approx \mathrm{S_b} + \left.\frac{\partial \mathrm{S_{static}}}{\partial \mathrm{G}}\right|_{\mathrm{G_b},\mathrm{N_b}} \Delta\mathrm{G}$. Thus, $\mathrm{k_{Sp}}$ should be described as an aggregate local glucose sensitivity rather than as a direct rate constant for a single biochemical reaction. A schematic chain-rule view is: $\frac{\partial \mathrm{S_{static}}}{\partial \mathrm{G}} \sim \frac{\partial \mathrm{S}}{\partial \mathrm{C_\mu}} \frac{\partial \mathrm{C_\mu}}{\partial \mathrm{V_m}} \frac{\partial \mathrm{V_m}}{\partial \mathrm{K_{ATP}}} \frac{\partial \mathrm{K_{ATP}}}{\partial \mathrm{ATP/ADP}} \frac{\partial \mathrm{ATP/ADP}}{\partial \mathrm{G}} + \frac{\partial \mathrm{S}}{\partial \mathrm{A_G}} \frac{\partial \mathrm{A_G}}{\partial \mathrm{G}}$. Here, $\mathrm{C_\mu}$ denotes local calcium microdomain signaling, $\mathrm{V_m}$ denotes membrane potential, $\mathrm{K_{ATP}}$ denotes KATP-channel activity, and $\mathrm{A_G}$ denotes an effective glucose-dependent amplifying signal. This expression should be presented as a conceptual decomposition, not as a uniquely identifiable biochemical formula. It states that the fitted proportional gain summarizes the net local effect of triggering and amplifying mechanisms. This interpretation is supported by mechanistic and systems biology models of glucose sensing. Fridly and Philipson developed a computational systems analysis in which rising glucose increases oxidative metabolism and ATP/ADP, leading to KATP-channel closure, depolarization, calcium influx, and insulin secretion[13]. More recent SBML-based pathway modeling of glucose-stimulated insulin secretion has integrated data across multiple experimental systems and linked glycolysis, ATP/ADP dynamics, and biphasic insulin secretion[16]. These models support the interpretation of the proportional term as a local linear approximation to a complex nonlinear glucose-secretion relation.

The potentiation term ($\mathrm{k_{Si}}(G - G_\mathrm{b})t$) should be interpreted cautiously. It should not be described as if beta cells literally compute an indefinite mathematical integral of glucose. A more physiologically accurate interpretation is that the integral term approximates slow, finite-memory potentiation over the analyzed postprandial window. Candidate processes include granule mobilization, docking, priming, refilling of release-competent pools, metabolic amplification, incretin/cAMP-dependent potentiation, and delayed changes in beta-cell secretory competence[1,11,12,14–18]. A generic potentiation state can be written as: $\tau_\mathrm{Q}\dot{\mathrm{Q}}(\mathrm{t}) = \mathrm{a_Q}\left(\Delta\mathrm{G}(\mathrm{t})\right)_+ - \mathrm{Q}(\mathrm{t})$ with solution $\mathrm{Q}(\mathrm{t}) = \mathrm{Q}(\mathrm{t_0})e^{-(\mathrm{t}-\mathrm{t_0})/\tau_\mathrm{Q}} + \frac{\mathrm{a_Q}}{\tau_\mathrm{Q}}\int_{\mathrm{t_0}}^{\mathrm{t}} e^{-(\mathrm{t}-\mathrm{s})/\tau_\mathrm{Q}}\left(\Delta\mathrm{G}(\mathrm{s})\right)_+ \mathrm{ds}$. The empirical model uses the simpler approximation. This approximation is reasonable only within a defined OGTT or postprandial interval and should not be extrapolated indefinitely. It is most defensible when the relevant slow state changes over the same several-hour time scale as the experiment, when saturation and depletion are not dominant, and when the model is used descriptively rather than mechanistically outside the fitted time window. This interpretation aligns with the potentiation component of the Mari/Tura/Ferrannini model, in which the glucose dose-response is modulated by a time-varying potentiation factor during meal or OGTT data[1,17]. It also aligns with granule-pool models in which slowly evolving mobilization, priming, and refilling processes shape second-phase secretion[14,15]. The coefficient $\mathrm{k_{Si}}$ should therefore be described as an effective time-dependent potentiation gain.

The derivative-like term represents early secretion that is preferentially recruited during rising glucose. Several biological mechanisms can generate a rate-sensitive transient after reduction to a low-dimensional input-output form. One mechanistic route is fast secretory drive followed by slower adaptation or depletion. Let a rapid fusion drive be: $F(t) = F_G\big(G(t)\big)$, and let a slower adaptive or depletion state follow that drive: $\tau_A \dot{A}(t) = F(t) - A(t)$. The transient component is: $S_{tr}(t) = F(t) - A(t)$. When the input changes smoothly relative to the adaptive time scale, this gives the local approximation: $S_{tr}(t) \approx \tau_A \frac{dF}{dt} = \tau_A F_G'\big(G(t)\big)\frac{dG(t)}{dt}$. Over a limited glucose range, $F_G'\big(G(t)\big)$ can be absorbed into an effective gain, yielding a derivative-like regressor. The use of $\dot{G}_+$ rather than the fully signed derivative is an empirical rectification that focuses on upward glucose transitions, where early recruitment of release-competent granules and first-phase secretion are most prominent.

A second route is the threshold-distribution view of insulin packets. Grodsky proposed that labile insulin is stored in packets with a distribution of glucose thresholds and that packets respond rapidly once their thresholds are reached[19]. If releasable packets are indexed by threshold θ, with density ρ(θ) and labile content L(θ, t), then rising glucose recruits newly crossed threshold classes. Locally, this contribution can be written as: $S_{thr}(t) = q\rho\big(G(t)\big)L(G(t), t)\dot{G}_+$. If ρ(G)L(G,t) changes slowly over the local range, it reduces to: $S_{thr}(t) \approx k_{Sd}\dot{G}_+$. This provides a direct rationale for the positive derivative-like term as an upward-threshold-crossing or recruitment term. However, the approximation is local. During large glucose excursions, repeated stimulation, depletion, or refilling, the effective coefficient multiplying $\dot{G}_+$ should be expected to depend on glucose level and prior secretory history.

A third route is granule-pool dynamics. A simplified release-competent pool model can be written as: $\dot{R}(t) = m(G, C, N) - f\big(C_\mu\big)R(t), S(t) = f\big(C_\mu\big)R(t)$, where R(t) denotes a release-competent pool, m denotes mobilization, docking, priming, or refilling, C denotes cytosolic calcium, and $C_\mu$ denotes calcium microdomain signaling. Kinetic models of beta-cell exocytosis show how fast fusion near calcium microdomains and slower resupply or priming can coexist in the same system[14]. Models fitted to experimental calcium and secretion data show that calcium dynamics and granule-pool dynamics jointly shape biphasic insulin secretion and glucose amplification[15]. After reduction, fast recruitment contributes to $\dot{G}_+$, and local steady glucose sensitivity contributes to ΔG(t).

Autonomic regulation may also contribute to early postprandial insulin dynamics. Vagal input can provide insulin-promoting cholinergic effects during cephalic and early postprandial phases. It can be driven by sensory, gastrointestinal, and central inputs. Moreover, vagal effects can include inhibitory components. Recent experiments indicate that vagal activation can inhibit insulin release through neuronal nitric oxide synthase, highlighting that vagal regulation can include both stimulatory and suppressive branches depending on physiological state[20]. A reduced representation is: $S_V(t) = g_A A(t) - g_R R(t), \tau_A \dot{A}(t) = u_V(t) - A(t), \tau_R \dot{R}(t) = u_V(t) - R(t)$, where $u_V(t)$ is an unobserved meal-related vagal drive, A(t) is a relatively fast stimulatory branch, and R(t) is a slower inhibitory or counter-regulatory branch. If $u_V(t)$ is temporally aligned with early glucose appearance or rising glucose in a meal test, part of its net effect may be absorbed into the derivative-like regressor $\dot{G}(t)$. However, this should be stated as a possible unobserved contributor, not as an exclusive explanation of the derivative-like term. With glucose and insulin data alone, autonomic input is not identifiable separately from beta-cell rate sensitivity, incretin effects, or early glucose appearance.

<u>2. Estimation of glucose effectiveness and insulin sensitivity from hyperglycemic and hyperinsulinemic-euglycemic clamp tests</u>

This supplementary note describes a mathematical model used to estimate glucose effectiveness and insulin sensitivity from hyperglycemic and hyperinsulinemic-euglycemic clamp data. We first examined a basic model characterizing glucose kinetics, as follows:

$$\dot{G} = -k_{\mathrm{Glu}}(G - G_{\mathrm{b}}) - k_{\mathrm{Sen}}G(I - I_{\mathrm{b}}) + f_{\mathrm{G}}, \qquad \text{Eq. (30)}$$

where $k_{\mathrm{Glu}}$ and $k_{\mathrm{Sen}}$ represent glucose effectiveness and insulin sensitivity, respectively. $f_{\mathrm{G}}$ denotes the glucose infusion rate. Under steady-state conditions during the clamp tests, Eq. (30) can be analytically transformed into:

$$k_{\mathrm{Glu}} = \frac{-f_{\mathrm{Gg}}G_{\mathrm{i}}(I_0 - I_{\mathrm{i}}) + f_{\mathrm{Gi}}G_{\mathrm{g}}(I_0 - I_{\mathrm{g}})}{G_{\mathrm{i}}(G_0 - G_{\mathrm{g}})(I_0 - I_{\mathrm{i}}) - G_{\mathrm{g}}(G_0 - G_{\mathrm{i}})(I_0 - I_{\mathrm{g}})}, \qquad \text{Eq. (31)}$$

$$k_{\mathrm{Sen}} = \frac{f_{\mathrm{Gg}}(G_0 - G_{\mathrm{i}}) - f_{\mathrm{Gi}}(G_0 - G_{\mathrm{g}})}{G_{\mathrm{i}}(G_0 - G_{\mathrm{g}})(I_0 - I_{\mathrm{i}}) - G_{\mathrm{g}}(G_0 - G_{\mathrm{i}})(I_0 - I_{\mathrm{g}})}, \qquad \text{Eq. (32)}$$

where $G_{\mathrm{g}}$ and $G_{\mathrm{i}}$ represent steady-state plasma glucose concentrations during hyperglycemic and hyperinsulinemic-euglycemic clamps, respectively; $I_{\mathrm{g}}$ and $I_{\mathrm{i}}$ correspond to steady-state serum insulin levels during the respective clamp tests; and $f_{\mathrm{Gg}}$ and $f_{\mathrm{Gi}}$ denote the steady-state amounts of glucose infused during hyperglycemic and hyperinsulinemic-euglycemic clamps, respectively. Theoretically, the parameters $k_{\mathrm{Glu}}$ and $k_{\mathrm{Sen}}$ are unaffected by delays in insulin action, provided that the steady-state period is sufficiently prolonged, and thus this model is consistent with the minimal model of glucose regulation at the steady state[21]. Application of this model to individuals with normal glucose tolerance (NGT) yielded negative mean values for $k_{\mathrm{Glu}}$ (**Extended Data Fig. 1i**), which is a physiologically implausible result.

We then investigated a model with a first-order approximation around the equilibrium point:

$$\dot{G} = -k_{\mathrm{Glu}}(G - G_{\mathrm{b}}) - k_{\mathrm{Sen}}(I - I_{\mathrm{b}}) + f_{\mathrm{G}}. \qquad \text{Eq. (33)}$$

It is important to note that this model does not exclude the existence of nonlinear interactions between $G$ and $I$. Rather, our aim was to evaluate whether a model based on a first-order approximation around the equilibrium point could adequately characterize glucose dynamics even in the presence of potential nonlinear interactions at the extreme glucose and insulin concentrations encountered during clamp tests. At the steady state during the clamp tests, Eq. (33) can be transformed as follows:

$$k_{\mathrm{Glu}} = \frac{-f_{\mathrm{Gg}}(I_0 - I_{\mathrm{i}}) + f_{\mathrm{Gi}}(I_0 - I_{\mathrm{g}})}{(G_0 - G_{\mathrm{g}})(I_0 - I_{\mathrm{i}}) - (G_0 - G_{\mathrm{i}})(I_0 - I_{\mathrm{g}})}, \qquad \text{Eq. (34)}$$

$$k_{\mathrm{Sen}} = \frac{f_{\mathrm{Gg}}(G_0 - G_{\mathrm{i}}) - f_{\mathrm{Gi}}(G_0 - G_{\mathrm{g}})}{(G_0 - G_{\mathrm{g}})(I_0 - I_{\mathrm{i}}) - (G_0 - G_{\mathrm{i}})(I_0 - I_{\mathrm{g}})}. \qquad \text{Eq. (35)}$$

The mean value of $k_{\mathrm{Glu}}$ in NGT individuals was positive (**Extended Data Fig. 1j**). Based on these results, this first-order approximation model was selected for use in subsequent analyses.

To quantify the potential errors generated by Eq. (33) during blood glucose fluctuations, we examined the first 5 minutes of the hyperglycemic clamp test:

$$G_5 - G_0 = -k_{\mathrm{Glu}} \int_0^5 (G - G_{\mathrm{b}}) dt - k_{\mathrm{Sen}} \int_0^5 (I - I_{\mathrm{b}}) dt + \int_0^5 f_{\mathrm{G}} dt \qquad \text{Eq. (36)}$$

The discrepancy between the right and left sides of Eq. (36) (denoted as Res) was significantly different from zero (**Extended Data Fig. 1k**), indicating the presence of model error.

To overcome this limitation, we modified Eq. (33) based on the established two-compartment model of glucose kinetics[22], as follows:

$$\dot{G} = -k_{\mathrm{Glub}}(G - G_{\mathrm{b}}) - k_{\mathrm{GluG}} G + k_{\mathrm{GluQ}} Q - k_{\mathrm{Sen}}(I - I_{\mathrm{b}}) + f_{\mathrm{G}}, \qquad \text{Eq. (37)}$$

$$\dot{Q} = k_{\mathrm{GluG}} G - k_{\mathrm{GluQ}} Q. \qquad \text{Eq. (38)}$$

Here, $G$ denotes the plasma glucose concentration, whereas $Q$ denotes a glucose-equivalent amount in the returnable remote pool normalized by the same reference glucose distribution volume used for $G$. The parameters $k_{\mathrm{GluG}}$ and $k_{\mathrm{GluQ}}$ are rate parameters describing glucose exchange kinetics. At the steady state during the clamp tests, $k_{\mathrm{Glub}}$ and $k_{\mathrm{Sen}}$ can be estimated with Eqs. (34) and (35), respectively. During the first 5 minutes of the hyperglycemic clamp test, Eqs. (37) and (38) can be transformed as follows:

$$Q_5 - Q_0 = -k_{\mathrm{Glub}} \int_0^5 (G - G_{\mathrm{b}}) dt - k_{\mathrm{Sen}} \int_0^5 (I - I_{\mathrm{b}}) dt + \int_0^5 f_{\mathrm{G}} dt - (G_5 - G_0). \qquad \text{Eq. (39)}$$

Given that $Q_5$ is larger than $Q_0$ and the right-hand side of Eq. (39) (equivalent to Res) was positive, this two-compartment model (Eqs. (37) and (38)) would correct the error observed in the single-compartment model (Eq. (33)). By approximating the second-order derivative as zero, Eq. (38) was simplified to:

$$\frac{k_{\mathrm{GluG}}}{k_{\mathrm{GluQ}}} = \frac{Q_5 - Q_0}{G_5 - G_0}. \qquad \text{Eq. (40)}$$

Collectively, the parameters $k_{\mathrm{Glub}}$, $k_{\mathrm{Sen}}$, and $k_{\mathrm{GluG}}/k_{\mathrm{GluQ}}$ were estimated as follows:

$$k_{\mathrm{Glub}} = \frac{-f_{\mathrm{Gg}}(I_0 - I_{\mathrm{i}}) + f_{\mathrm{Gi}}(I_0 - I_{\mathrm{g}})}{(G_0 - G_{\mathrm{g}})(I_0 - I_{\mathrm{i}}) - (G_0 - G_{\mathrm{i}})(I_0 - I_{\mathrm{g}})}, \qquad \text{Eq. (41)}$$

$$k_{\mathrm{Sen}} = \frac{f_{\mathrm{Gg}}(G_0 - G_{\mathrm{i}}) - f_{\mathrm{Gi}}(G_0 - G_{\mathrm{g}})}{(G_0 - G_{\mathrm{g}})(I_0 - I_{\mathrm{i}}) - (G_0 - G_{\mathrm{i}})(I_0 - I_{\mathrm{g}})}, \qquad \text{Eq. (42)}$$

$$\frac{k_{\mathrm{GluG}}}{k_{\mathrm{GluQ}}} = -\left(1 + \frac{5k_{\mathrm{Glub}}}{2}\right) - \frac{5k_{\mathrm{Sen}}(I_5 - I_0)}{2(G_5 - G_0)} + \frac{F_{\mathrm{G0-G5}}}{G_5 - G_0}, \qquad \text{Eq. (43)}$$

where $F_{\mathrm{G0-G5}}$ represents the total glucose infused during the first 5 minutes of the hyperglycemic clamp. Of note, the $k_{\mathrm{GluG}}/k_{\mathrm{GluQ}}$ value of NGT from our clamp experiments was comparable to previously reported values ($k_{21}/k_{12}$) for NGT[22] (**Extended Data Fig. 1l**).

We then compared Eqs. (37) and (38) with alternative models incorporating an insulin action time delay[21] as follows:

$$\dot{G} = -k_{\mathrm{Glub}}(G - G_{\mathrm{b}}) - k_{\mathrm{GluG}} G + k_{\mathrm{GluQ}} Q - X + f_{\mathrm{G}}, \qquad \text{Eq. (44)}$$

$$\dot{Q} = k_{\mathrm{GluG}} G - k_{\mathrm{GluQ}} Q, \qquad \text{Eq. (45)}$$

$$\dot{X} = k_{\mathrm{Sen}} k_{\mathrm{SenX}}(I - I_{\mathrm{b}}) - k_{\mathrm{SenX}} X, \qquad \text{Eq. (46)}$$

where $X$ denotes the insulin action in the remote compartment. The MAE between the measured and simulated glucose levels using a meta-evolutionary programming algorithm combined with nonlinear least-squares optimization was lower with this time delay model than with the model without time delay (**Extended Data Fig. 1m**). Based on these analyses, we adopted Eqs. (44), (45) and (46) for our subsequent investigations.

To visualize how distinct components of glucose effectiveness ($k_{\mathrm{Glub}}$ and $k_{\mathrm{GluG}}/k_{\mathrm{GluQ}}$) shaped plasma glucose dynamics, we simulated intravenous glucose tolerance tests while independently varying $k_{\mathrm{Glub}}$ and $k_{\mathrm{GluG}}/k_{\mathrm{GluQ}}$. Glucose was administered as a constant intravenous input of 302 mg/kg over the first minute, followed by no exogenous glucose input from 1 to 240 min, and all remaining parameters were fixed at the mean values estimated in NGT participants. In this framework, $k_{\mathrm{Glub}}$ represents net irreversible removal of glucose from the accessible compartment, including renal glucose excretion and metabolic conversion into products that do not immediately re-enter the circulating glucose pool in tissues such as brain, adipose tissue, and skeletal muscle. By contrast, $k_{\mathrm{GluG}}$ and $k_{\mathrm{GluQ}}$ describe reversible glucose dynamics, including bidirectional exchange between plasma and interstitial compartments and conversion of glucose into hepatic intermediates that can later be reconverted to glucose as plasma glucose concentrations decline. These two parameters produced qualitatively distinct effects on plasma glucose trajectories. Increasing $k_{\mathrm{Glub}}$ lowered plasma glucose throughout the simulated IVGTT, indicating that this parameter primarily determines the overall magnitude of glucose disposal. In contrast, increasing $k_{\mathrm{GluG}}/k_{\mathrm{GluQ}}$ produced a biphasic effect: it suppressed plasma glucose during the early phase of the excursion but increased glucose concentrations during the later recovery phase (**Extended Data Fig. 1n**). We further evaluated the identifiability of these parameters using profile residual sum of squares (RSS) analysis, following previous studies[23–25]. For each parameter, the parameter of interest was fixed over a range from 0.1- to 10-fold of its best-fit value, while all remaining parameters were re-optimized. In both cases, ΔRSS increased as the fixed value deviated from the optimum, indicating that $k_{\mathrm{Glub}}$ and $k_{\mathrm{GluG}}/k_{\mathrm{GluQ}}$ are identifiable from the model fitting procedure (**Extended Data Fig. 1o**). Thus, irreversible glucose disposal and reversible glucose exchange contribute differently to glucose effectiveness. These simulations indicate that glucose effectiveness is not adequately represented by a single scalar clearance term, but instead consists of separable processes that differentially control the amplitude and temporal shape of glucose excursions. We note that incorporating this reversible plasma–remote exchange into glucose effectiveness extends the conventional usage, in which glucose effectiveness denotes net insulin-independent glucose disposal; we adopt the broader definition because the reversible flux materially shapes the glucose waveform and is identifiable from glucose data through the $k_{\mathrm{GluG}}/k_{\mathrm{GluQ}}$.

We then provide a coarse-grained physiological interpretation of the insulin sensitivity and glucose effectiveness. The delayed insulin action ($\dot{X} + k_{\mathrm{SenX}}X = k_{\mathrm{Sen}}k_{\mathrm{SenX}}(I - I_{\mathrm{b}})$) has a direct molecular interpretation. Insulin does not lower glucose simply by appearing in plasma. It must be transported to insulin-sensitive tissues, bind to insulin receptors, activate receptor and IRS phosphorylation, propagate PI3K-Akt signaling, regulate AS160/TBC1D4 and related trafficking machinery, and promote GLUT4 translocation in skeletal muscle and adipose tissue[26,27]. In the liver, insulin signaling through Akt and FoxO1-related mechanisms suppresses gluconeogenic programs and modulates glycogen metabolism, while insulin can also suppress hepatic glucose production indirectly through extrahepatic pathways[28,29]. These steps necessarily introduce delay and filtering between plasma insulin and the effective glucose-lowering response. Therefore, in the present model, $X$ should be interpreted as delayed insulin action or insulin-mediated glucose-lowering flux, rather than as insulin concentration itself. This interpretation is also consistent with the original rationale for the remote compartment in the minimal model, where a remote insulin-action variable was needed to reproduce the temporal delay between measured plasma insulin and observed acceleration of glucose disappearance during IVGTT[21,30].

The term $-k_{\mathrm{Glub}}(G - G_{\mathrm{b}})$ represents net irreversible removal of glucose from the accessible glucose pool. This term is called irreversible because it reduces the total amount of glucose-equivalent material represented by the model, rather than merely transferring glucose between compartments. Mechanistically, it summarizes processes such as glucose oxidation, glycolytic conversion to downstream metabolites that do not rapidly return to plasma glucose, incorporation into longer-lived metabolic pools, and renal glucose excretion when glycemia exceeds the renal handling capacity.

The pair of terms $G \underset{k_{\mathrm{GluQ}}}{\overset{k_{\mathrm{GluG}}}{\rightleftarrows}} Q$ represents reversible glucose redistribution between the accessible plasma compartment and a returnable glucose-equivalent pool. The state $Q$ should not be interpreted as a single molecule. Rather, it is a lumped pool that includes glucose temporarily residing outside the measured plasma compartment and glucose equivalents that can later contribute back to circulating glucose. There are at least two physiological bases for this term. First, glucose distributes between plasma and interstitial fluid. Plasma-to-interstitium glucose kinetics have been described by two-compartment linear models, and tracer/microdialysis studies have demonstrated measurable equilibration times between plasma and interstitial fluid[31]. Second, the liver can transiently convert incoming glucose into glucose-6-phosphate, glycogen, and related intermediates. Although some of this glucose is irreversibly oxidized or converted into non-returning products, a portion can later re-enter circulating glucose through glycogenolysis or changes in endogenous glucose production as plasma glucose declines. Setting the reversible subsystem to quasi-equilibrium yields $\dot{Q} = 0 \Rightarrow k_{\mathrm{GluG}}G = k_{\mathrm{GluQ}}Q$, which gives $\frac{Q}{G} = \frac{k_{\mathrm{GluG}}}{k_{\mathrm{GluQ}}}$. Thus, the ratio $k_{\mathrm{GluG}}/k_{\mathrm{GluQ}}$ has a mechanistic interpretation as the equilibrium partitioning of glucose equivalents into the returnable pool relative to the accessible plasma pool. A larger ratio means that a larger fraction of the glucose load is transiently stored or redistributed into a pool that can later return to plasma glucose.

<u>3. Validation of the simplified glucose-insulin model using *HuMet Repository* OGTT data</u>

To test whether the simplified glucose–insulin model developed in this study generalized beyond clamp experiments, we evaluated its ability to reproduce oral glucose tolerance test (OGTT) dynamics using an external dataset from the *HuMet Repository*[32] (**Extended Data Fig. 2a**). The *HuMet Repository* includes data from an investigation of dynamic human metabolism in 15 healthy men (mean age 27.8 ± 2.9 years; mean body mass index 23.1 ± 1.8 kg/m$^2$) who were not taking medication and had no metabolic abnormalities. An OGTT was administered on day 3 of the study protocol. After an overnight fast, each participant ingested 300 mL of a glucose solution containing mono- and oligosaccharides that yielded 75 g of glucose equivalents after enzymatic cleavage, and consumed it within 5 min. Blood samples were collected at 0, 15, 30, 45, 60, 90, 120, 180, and 240 min after ingestion. For the present analysis, all 15 participants with complete plasma glucose and serum insulin profiles were included.

We compared the model developed in this study with two previously published models: the insulin secretion and sensitivity model of Ha et al. (Previous study 1)[3,33] and the meal simulation model of Dalla Man et al. (Previous study 2)[34]. To isolate differences in glucose–insulin regulatory structure from differences in oral glucose appearance, the glucose input function in our model was set to the Ha et al. formulation when compared with the Ha et al. model, and to the Dalla Man et al. formulation when compared with the Dalla Man et al. model. Because only glucose and insulin trajectories were fitted in this analysis, parameter estimation was restricted to $k_{\mathrm{Glub}}$, $k_{\mathrm{GluG}}/k_{\mathrm{GluQ}}$, $k_{\mathrm{Sen}}$, $k_{\mathrm{Sp}}$, $k_{\mathrm{Si}}$, $k_{\mathrm{Sd}}$, $k_{\mathrm{Ic}}$.

All three models captured the overall temporal profiles of glucose and insulin during the OGTT. However, the simplified model developed in this study showed lower normalized mean squared error (MSE) than the Ha et al. model for both glucose and insulin trajectories. Its performance was comparable to that of the Dalla Man meal simulation model, despite its reduced structural complexity. Thus, the simplified glucose–insulin model reproduced OGTT glucose and insulin dynamics in an independent human dataset with accuracy comparable to a detailed meal simulation model.

##### 4. Validation using data from the *University of Tokyo Study*

We next tested model performance using data from the *University of Tokyo Study*[35], in which plasma glucose and serum insulin responses were measured after bolus ingestion of 25, 50, or 75 g of glucose (**Extended Data Fig. 2b**). This dataset provided an additional validation setting because the model was required to reproduce glucose–insulin dynamics across multiple oral glucose doses rather than under a single standardized OGTT condition. The *University of Tokyo Study* enrolled three healthy volunteers with HbA1c values of 4.9–5.4% and no diagnosis of diabetes. After a 10-h overnight fast, each participant underwent glucose administration at doses of 25, 50, and 75 g under two ingestion protocols: rapid bolus ingestion completed within 1 min and continuous ingestion over 2 h. Continuous ingestion was delivered through tubing by a non-contact micro-dispensing robot (Mr. MJ; MECT Corporation). All glucose solutions (TRELAN-G75; Ajinomoto) were diluted with distilled water to a fixed total volume of 225 mL to standardize the ingested volume. Each dose-by-protocol condition was tested in the same participants in separate sessions held at least 1 month apart. Blood samples were collected from a superficial forearm vein at 0 min and every 10 min until 240 min. For the present validation, we analyzed the three bolus-ingestion dose conditions: 25, 50, and 75 g. The model was fitted to the glucose and insulin trajectories for each condition, as described for the *HuMet Repository* analysis, and accuracy was quantified using normalized mean squared error and compared with that of the Ha et al. and Dalla Man et al. models.

All three models captured the principal features of the observed glucose and insulin responses across glucose doses. The simplified model developed in this study reproduced the dose-dependent glucose excursions and the major temporal features of the insulin response, including the early post-load rise and subsequent decline toward baseline. When fitting accuracy was quantified by normalized mean squared error, the performance of the simplified model was comparable to that of the Ha et al. and Dalla Man et al. models. For glucose, paired differences in normalized mean squared error between the simplified model and each previous model were distributed around zero, indicating similar accuracy in reproducing glucose trajectories. Similar results were observed for insulin dynamics. Thus, the simplified glucose–insulin model reproduced oral glucose and insulin responses across multiple glucose-dose conditions in this validation dataset with accuracy comparable to that of previously published models.

5. Validation using data from the *Kobe University Study*

We further evaluated the model using data from the *Kobe University Study*, which included 121 individuals with normal glucose tolerance (NGT), impaired glucose tolerance (IGT), or type 2 diabetes mellitus (T2DM). This dataset allowed us to test whether the simplified glucose–insulin model could reproduce oral glucose tolerance test (OGTT) dynamics across a broad spectrum of glucose tolerance. During the 75-g OGTT, measured plasma glucose trajectories differed markedly across glucose tolerance groups, with progressively larger and more sustained glucose excursions from NGT to IGT and T2DM (**Extended Data Fig. 2c**).

The model developed in this study reproduced this between-group separation of glucose trajectories and captured the overall insulin response profiles across the three groups. When fitting accuracy was quantified using normalized mean squared error (NMSE), the simplified model performed similarly to the Dalla Man et al. meal simulation model and showed lower NMSE than the Ha et al. model in most comparisons. For glucose, paired differences in NMSE between the simplified model and the Ha et al. model were negative when calculated as NMSE_simplified − NMSE_Ha, indicating lower error for the simplified model. In contrast, the differences relative to the Dalla Man et al. model were distributed around zero, indicating comparable accuracy. Similar trends were observed for insulin dynamics, for which the simplified model showed lower or comparable error relative to the Ha et al. model and comparable performance relative to the Dalla Man et al. model. Together with the *HuMet Repository* and *University of Tokyo Study* analyses, these results suggest that the simplified glucose–insulin model reproduces OGTT glucose and insulin dynamics across independent datasets, multiple glucose doses, and different glucose tolerance states with accuracy comparable to previously published models.

6. Validation of the mathematical model using *PREDICT1* mixed-meal tolerance test data

We next asked whether the simplified glucose–insulin model could reproduce glucose–insulin dynamics under a nutritional challenge more complex than isolated glucose administration. To this end, we analyzed mixed-meal tolerance test (MMTT) data from the *PREDICT1 Study*[36] (**Extended Data Fig. 3a**). The *PREDICT1 Study* was a multinational clinical study of postprandial metabolic responses to standardized meals. Participants attended an in-clinic visit at 08:30 after an overnight fast beginning at 21:00 on the previous evening. Before the visit, participants refrained from exercise, limited fat, fiber, and alcohol intake during the preceding 24 h, and avoided caffeine after 18:00 on the previous evening. After an antecubital vein was cannulated and a baseline fasting sample was collected, each participant consumed a standardized breakfast consisting of muffins and a milkshake, providing 86 g of carbohydrate and 53 g of fat, within 10 min. Venous blood samples were collected at 0 min, corresponding to the fasting baseline, and at 15, 30, 60, 120, 180, and 240 min after the meal. For the present analysis, we used the postprandial plasma glucose and serum insulin time courses after breakfast from 903 participants with complete data. Participant-specific parameters were fitted by minimizing the normalized residuals between measured and simulated concentrations.

Despite the additional physiological complexity of mixed meals, which may reflect nutrient inputs other than glucose as well as meal-dependent differences in absorption and hormonal responses, the model captured the overall temporal profiles of both glucose and insulin. Across all participants and sampling time points, simulated glucose concentrations correlated well with measured glucose concentrations ($R = 0.85$, $P < 0.01$), and simulated insulin concentrations correlated well with measured insulin concentrations ($R = 0.91$, $P < 0.01$). These results suggest that the model can reproduce glucose–insulin dynamics beyond glucose-only challenges.

7. Validation of the mathematical model using porcine intravenous glucose tolerance test data

To test whether the simplified glucose–insulin model could reproduce glucose regulatory dynamics beyond human datasets, we applied it to porcine intravenous glucose tolerance test (IVGTT) data[37] (**Extended Data Fig. 3b**). The porcine IVGTT dataset included 18 pigs from two litters bred at the Swedish Livestock Research Centre (Lövsta): litter 1 (n = 8; 4 females and 4 males; Yorkshire × Swedish Landrace; mean body weight, 17.1 ± 0.4 kg at approximately 2 months of age) and litter 2 (n = 10; 5 females and 5 males; Yorkshire × Swedish Landrace × Hampshire; mean body weight, 32.7 ± 0.8 kg at approximately 3 months of age). After a 2-week acclimatization period that included training for scale weighing and bottle feeding, an indwelling silicone catheter was implanted into the jugular vein under aseptic conditions and general anesthesia. The catheter was tunneled subcutaneously to an exit point between the scapulae, and catheters were flushed twice daily with heparinized saline. Food was withdrawn 1 h before the test. For the IVGTT, glucose (Glucose Fresenius Kabi, 500 mg/mL) was administered intravenously at 0.5 g/kg body weight over 1 min. Blood samples were collected before infusion and at 5, 10, 20, 30, 45, 60, 90, 120, and 180 min after infusion. Blood glucose was measured immediately using Accu-Chek test strips (Roche) validated for porcine blood, and plasma insulin was measured using a porcine-specific enzyme-linked immunosorbent assay (10-1200-01; Mercodia), with all samples run in duplicate and duplicate measurements showing a coefficient of variation <10%. For the present analysis, animal-specific parameters were fitted to each pig's glucose and insulin trajectories.

The fitted model reproduced the principal features of the porcine IVGTT response, including the rapid decline in plasma glucose after intravenous loading and the accompanying glucose-stimulated insulin response. Across pooled observations from 18 pigs and 10 sampling time points, simulated glucose concentrations were strongly correlated with measured glucose concentrations ($R = 0.95$, $P < 0.01$). Simulated insulin concentrations also showed a strong correlation with measured insulin concentrations ($R = 0.98$, $P < 0.01$). These results indicate that the simplified model captures both glucose disappearance and insulin secretory dynamics during IVGTT in pigs, supporting the generalizability of the model beyond human glucose challenge datasets.

8. *In silico* validation of the simplified glucose–insulin model against IVGTT simulations generated by the Sorensen model

We next tested whether the simplified glucose–insulin model could approximate the dynamics generated by a more detailed physiological model under intravenous glucose perturbation. To this end, we generated reference intravenous glucose tolerance test (IVGTT) trajectories using the revised Sorensen model[38] and fitted the simplified model to the simulated glucose and insulin time courses (**Extended Data Fig. 3c**). The revised Sorensen model is a detailed physiological representation of the glucose–insulin system, comprising 22 coupled ordinary differential equations and approximately 135 parameters. It explicitly describes glucose and insulin concentrations across brain, heart–lung, liver, gut, kidney, and peripheral compartments, and incorporates a pancreatic submodel of insulin secretion with first- and second-phase components, as well as renal glucose excretion. In its original description, this revised model was calibrated and validated against published glycemic and insulinemic responses to glucose perturbations.

Reference IVGTT trajectories were generated using the publicly available implementation of the revised Sorensen model with the default parameter set reported in the original study. The Sorensen model was first integrated until numerical steady state was reached to obtain physiologically consistent initial conditions. At steady state, the model had a basal venous plasma glucose concentration of 89.0 mg/dL and a basal plasma insulin concentration of 12.8 mU/L, with the simulated body weight set to 70 kg. IVGTT simulations were then performed at three glucose doses: 0.4, 0.5, and 0.6 g/kg. For each dose, the intravenous glucose challenge was modeled as a constant infusion over the first 3 min. Simulated venous plasma glucose and plasma insulin concentrations were sampled at 0, 5, 10, 15, 20, 25, 30, 35, 40, 45, 50, 55, 60, 65, 70, 75, 80, 90, 100, 110, 120, 140, 160, 180, and 200 min after the start of glucose infusion and used as target data. The simplified glucose–insulin model developed in this study was then fitted to the three Sorensen-generated reference trajectories using the same parameter estimation procedure as in the human clamp analysis. A single common parameter set was estimated simultaneously across the three glucose doses, with the exogenous glucose infusion schedule provided as model input. Parameter values were optimized by minimizing the normalized residual sum of squares between the Sorensen-generated reference trajectories and the trajectories simulated by the simplified model, using an evolutionary programming algorithm combined with nonlinear least-squares optimization implemented in SciPy 1.10.1.

Despite its reduced structure, the simplified model closely reproduced the glucose and insulin dynamics generated by the detailed physiological model across all glucose doses. Across pooled dose–time observations, fitted glucose concentrations closely matched the Sorensen model reference values, with a fitted-versus-reference correlation of $R = 0.99$ ($P < 0.01$). Insulin dynamics were also closely reproduced, with a pooled fitted-versus-reference correlation of $R = 0.99$ ($P < 0.01$). These results indicate that the simplified model can capture the dominant glucose–insulin dynamics of a detailed physiological simulator under graded intravenous glucose challenges using a single common parameter set across doses.

9. *In silico* validation of C-peptide dynamics under graded glucose infusion

We next tested whether the C-peptide component of the simplified glucose–insulin model could reproduce C-peptide dynamics generated by an established model of β-cell secretion and C-peptide kinetics[10]. Although the insulin secretion terms in the Toffolo model and in our model can be analytically related under the approximations described in Eqs. 26–29, we asked whether the two formulations also produced similar dynamic behavior in numerical simulations. To this end, we performed an *in silico* model-comparison analysis using the graded up-and-down glucose infusion protocol of Toffolo et al.[10] as a reference system (**Extended Data Fig. 3d**). In the original study, this graded up-and-down infusion protocol was applied to nondiabetic individuals and was designed to characterize the β-cell glucose dose–response relationship across a wide range of glucose infusion rates, thereby providing a useful benchmark for the secretion model.

The reference C-peptide model was the M1 model of Toffolo et al.[10], which couples a two-compartment description of C-peptide kinetics, with a secretion model controlled by both static and dynamic components of plasma glucose:

$$\frac{d\mathrm{CP}_1}{dt} = -(k_{01} + k_{21})\mathrm{CP}_1(t) + k_{12}\mathrm{CP}_2(t) + \mathrm{SR}(t), \qquad \mathrm{CP}_1(0) = 0,$$

$$\frac{d\mathrm{CP}_2}{dt} = k_{21}\mathrm{CP}_1(t) - k_{12}\mathrm{CP}_2(t), \qquad \mathrm{CP}_2(0) = 0,$$

$$\mathrm{SR}(t) = \mathrm{SR}_\mathrm{s}(t) + \mathrm{SR}_\mathrm{d}(t),$$

$$\mathrm{SR}_\mathrm{s}(t) = Y(t),$$

$$\frac{dY}{dt} = -\alpha\{Y(t) - \beta[G(t) - G_\mathrm{b}]\}, \qquad Y(0) = 0,$$

$$\mathrm{SR}_\mathrm{d}(t) = \begin{cases} k_\mathrm{d}\dfrac{dG}{dt}, & \text{if } \dfrac{dG}{dt} > 0 \text{ and } G(t) > G_\mathrm{b}, \\ 0, & \text{otherwise.} \end{cases}$$

Here, $\mathrm{CP}_1$ and $\mathrm{CP}_2$ (pmol/L) denote C-peptide concentrations above basal in the accessible and peripheral compartments; $k_{01}$, $k_{21}$, and $k_{12}$ ($\mathrm{min}^{-1}$) are kinetic rate parameters; Y(t) is the provision of new insulin to beta cells; α ($\mathrm{min}^{-1}$) is the rate constant of provision; β ($\mathrm{min}^{-1}$) is the static beta-cell sensitivity to glucose; and $k_\mathrm{d}$ describes the dynamic control of glucose on insulin secretion.

The simulated protocol consisted of seven consecutive 40-min glucose infusion periods at 0, 4, 8, 16, 8, 4, and 0 mg $\mathrm{kg}^{-1}$ $\mathrm{min}^{-1}$. The resulting C-peptide trajectories were used as target data. The values of α, β, and $k_\mathrm{d}$ were taken from the individual estimates of model M1 reported by Toffolo et al. Because numerical values for individual plasma glucose time courses were not available, the mean plasma glucose trajectory reported in the original study was extracted from the published figure using WebPlotDigitizer[39] and linearly interpolated between sampled time points to define the glucose input G(t). To assess the accuracy of this digitization procedure, we applied the same extraction workflow to the *Kobe University Study* data for which the original numerical values were available; the reconstruction error was less than 1%. Although this digitized glucose input did not provide individual-specific glucose trajectories, the objective of this analysis was not to refit individual experimental data, but rather to test whether the simplified C-peptide subsystem developed in this study could reproduce the dynamic behavior of an established mathematical model. We therefore considered the reconstructed mean glucose input sufficient for this *in silico* model-comparison analysis. Reference C-peptide trajectories were obtained by simulating the Toffolo model equations for each individual using individual parameter estimates and the common reconstructed mean glucose input. The C-peptide subsystem developed in this study was then fitted separately to each reference trajectory by minimizing the normalized residual sum of squares using an evolutionary programming algorithm combined with nonlinear least-squares optimization. Goodness of fit was evaluated by inspecting normalized residuals over the full graded glucose infusion protocol.

The fitted model closely reproduced the reference C-peptide dynamics over the entire 0- to 240-min protocol, including the gradual rise during increasing glucose infusion, the peak during the maximal glucose input period, and the subsequent decline as glucose infusion decreased. Across all eight individuals, normalized residuals were centered near zero throughout the protocol, indicating little systematic deviation from the reference model. Thus, the C-peptide formulation used in this study not only approximates the previously established model analytically but also recapitulates its dynamic behavior under graded glucose stimulation.

10. *In silico* validation of glucose dynamics using IVGTT data

We next asked whether the glucose subsystem of our model could reproduce glucose trajectories generated by an established IVGTT model[22]. Because the glucose equation in our model represents a first-order approximation around equilibrium, whereas the established two-compartment minimal model (2CMM) includes nonlinear insulin-dependent effects, this analysis tested whether the simplified formulation could approximate 2CMM-generated glucose trajectories during IVGTT-induced hyperglycemic excursions. We used the 2CMM as a reference system and treated the glucose trajectories generated by this model as target data for fitting.

The reference glucose model was the 2CMM developed by Cobelli et al.[22] which extends the classical single-compartment minimal model of Bergman et al. by appending a non-accessible glucose compartment. The model is described by the following equations:

$$\frac{dQ_1}{dt} = -[p_1 + k_{21} + X(t)]Q_1(t) + k_{12}Q_2(t) + p_1 Q_{1\mathrm{b}}, \qquad Q_1(0) = Q_{1\mathrm{b}} + D$$

$$\frac{dQ_2}{dt} = k_{21}Q_1(t) - k_{12}Q_2(t), \qquad Q_2(0) = Q_{2\mathrm{b}}$$

$$\frac{dX}{dt} = -p_2 X(t) + p_3[I(t) - I_\mathrm{b}], \qquad X(0) = 0$$

$$G(t) = \frac{Q_1(t)}{V_1}$$

Here, $Q_1$ and $Q_2$ (mg/kg) denote glucose masses in the accessible and non-accessible compartments, respectively; X(t) is the insulin action variable in a compartment remote from plasma; D is the intravenous glucose dose (mg/kg); $V_1$ is the volume of the accessible compartment; $k_{21}$ and $k_{12}$ are exchange rate parameters between compartments; $p_1$ is the glucose effectiveness rate constant; and $p_2$ and $p_3$ parameterize the dynamics of insulin action. Reference simulations were generated using the Bayesian parameter estimates for 22 normal human individuals reported by Cobelli et al. The IVGTT glucose dose was set to the reported mean value of 302 mg/kg and administered as a 1-min rectangular intravenous infusion, consistent with the original study. The mean plasma insulin time course reported in the previous study[40] was used as the model input I(t), and $Q_1(0)$ and $Q_2(0)$ were initialized to the corresponding steady-state values for the basal plasma glucose concentration. The 2CMM equations were integrated numerically to generate reference plasma glucose trajectories, which were used as target data. The glucose subsystem of the model developed in this study was then fitted to the reference trajectories by minimizing the normalized residual sum of squares using an evolutionary programming algorithm combined with nonlinear least-squares optimization. Goodness of fit was evaluated by comparing the fitted and reference trajectories and by examining normalized residuals.

The glucose subsystem developed in this study closely reproduced the 2CMM-generated reference glucose trajectories (**Extended Data Fig. 3e**). The fitted curves captured both the rapid early decline after glucose administration and the slower return toward baseline during the later phase. Normalized residuals remained close to zero throughout the simulation period, including the early post-bolus interval, when two-compartment exchange is expected to contribute substantially to the glucose curve. Thus, under the simulated IVGTT condition examined here, the first-order glucose formulation approximated the behavior of a nonlinear two-compartment IVGTT model.

11. *In silico* validation of insulin and C-peptide dynamics using insulin-modified IVGTT data

We then evaluated whether the insulin and C-peptide subsystems of the model could reproduce trajectories generated by an established insulin-modified intravenous glucose tolerance test (IM-IVGTT) model[41]. Reference simulations were based on the protocol of Toffolo et al.[41], in which healthy participants received an intravenous glucose bolus of 0.3 g/kg body weight, followed by a square-wave insulin infusion of 0.02 U/kg body weight from 20 to 25 min. The insulin and C-peptide trajectories generated by the models were used as target data for fitting.

The reference insulin-C-peptide models were those of Toffolo et al.[41], in which a C-peptide minimal model of secretion and kinetics is coupled to a single-compartment model of insulin delivery rate and kinetics. The C-peptide subsystem is described by the following equations:

$$\frac{d\mathrm{CP}_1}{dt} = -(k_{01} + k_{21})\mathrm{CP}_1(t) + k_{12}\mathrm{CP}_2(t) + \mathrm{isr}(t),$$

$$\frac{d\mathrm{CP}_2}{dt} = k_{21}\mathrm{CP}_1(t) - k_{12}\mathrm{CP}_2(t),$$

$$\mathrm{isr}(t) = mX(t),$$

$$\frac{dX}{dt} = -mX(t) + Y(t), \qquad X(0) = X_0,$$

$$\frac{dY}{dt} = -\alpha\{Y(t) - \beta[G(t) - h]\}, \qquad Y(0) = 0.$$

Here, $X_0$ accounts for first-phase secretion in response to the glucose bolus, $Y(t)$ represents the slower provision of new insulin to beta cells, $m$ is the rate constant of secretion, $\alpha$ is the rate constant of provision, $\beta$ is the static responsivity of provision to glucose above the threshold $h$, and $\mathrm{isr}(t)$ is the C-peptide secretion rate above basal, normalized to the accessible distribution volume. The insulin subsystem is described by the following equations:

$$\frac{dI}{dt} = -nI(t) + \mathrm{idr}(t) + \frac{U(t)}{V_\mathrm{I}}, \qquad I(0) = 0,$$

$$\mathrm{idr}(t) = m^{\mathrm{IDR}}X^{\mathrm{IDR}}(t),$$

$$\frac{dX^{\mathrm{IDR}}}{dt} = -m^{\mathrm{IDR}}X^{\mathrm{IDR}}(t) + Y^{\mathrm{IDR}}(t), \qquad X^{\mathrm{IDR}}(0) = X_0^{\mathrm{IDR}},$$

$$\frac{dY^{\mathrm{IDR}}}{dt} = -\alpha^{\mathrm{IDR}}\{Y^{\mathrm{IDR}}(t) - \beta^{\mathrm{IDR}}[G(t) - h]\}, \qquad Y^{\mathrm{IDR}}(0) = 0.$$

Here, $I(t)$ is the plasma insulin concentration above basal, $n$ is the rate constant of insulin disappearance, $V_\mathrm{I}$ is the insulin distribution volume, $U(t)$ is the exogenous insulin input, and $\mathrm{idr}(t)$ is the posthepatic insulin delivery rate above basal, normalized to $V_\mathrm{I}$. The exogenous insulin input $U(t)$ was nonzero only during the 20- to 25-min square-wave infusion of 0.02 U/kg. The secretory parameters $m^{\mathrm{IDR}}$, $\alpha^{\mathrm{IDR}}$, $\beta^{\mathrm{IDR}}$, and $X_0^{\mathrm{IDR}}$ have the same structural meaning as in the C-peptide subsystem but additionally absorb the effects of hepatic insulin extraction. The threshold parameter $h$ was constrained to take the same value in both subsystems.

Reference simulations were generated using the parameter values reported by Toffolo et al.[41] for 20 healthy individuals, with C-peptide kinetic parameters fixed to the population values of Van Cauter et al. [6]. Because individual glucose time courses were not available in numerical form, the mean glucose time course reported by Toffolo et al. during the IM-IVGTT was extracted from the published figure using WebPlotDigitizer, sampled at the original measurement times, and used as the glucose input $G(t)$. The resulting C-peptide and insulin trajectories were used as target data. The C-peptide and insulin subsystems of the model developed in this study were then fitted simultaneously to these target trajectories by minimizing the normalized residual sum of squares using a meta-evolutionary programming algorithm combined with nonlinear least-squares optimization. The exogenous insulin input $U(t)$ was incorporated explicitly in the fitted model with the same square-wave profile as in the reference simulation.

The model developed in this study reproduced the principal features of both simulated insulin and C-peptide dynamics (**Extended Data Fig. 3f**). For C-peptide, the fitted model captured the early glucose-stimulated rise, the subsequent decline, and the gradual return toward baseline. For insulin, the model reproduced both the endogenous glucose-stimulated response and the transient increase caused by exogenous insulin administration during the IM-IVGTT. These results indicate that the insulin–C-peptide subsystem recapitulates established model behavior under a perturbation that combines endogenous beta-cell stimulation with exogenous insulin input.

12. Derivation of an equation relating blood glucose levels to the parameters governing glucose homeostasis

This section derives a relationship between blood-glucose dynamics and parameters that govern glucose regulation under physiological conditions, *i.e.*, no exogenous insulin administration ($f_{\mathrm{I}} = 0$). By integrating Eqs. (1), (44), (45), and (46) from one steady state to another, we obtained the following equations:

$$0 = -k_{\mathrm{Glub}} \int (G - G_{\mathrm{b}})dt - k_{\mathrm{Sen}} \int (I - I_{\mathrm{b}})dt + F, \qquad \text{Eq. (47)}$$

$$0 = k_{\mathrm{r}} k_{\mathrm{Sp}} \int G dt + k_{\mathrm{r}} k_{\mathrm{Si}} \int (G - G_{\mathrm{b}})_{+} t dt + k_{\mathrm{r}} k_{\mathrm{Sd}} \int \dot{G}_{+} dt - k_{\mathrm{Ic}} \int I dt\,, \qquad \text{Eq. (48)}$$

where $F$ denotes the time integral of the exogenous glucose input rate. Eqs (47) and (48) can be transformed as follows:

$$F = \left(k_{\mathrm{Glub}} + \frac{k_{\mathrm{Sen}} k_{\mathrm{r}} k_{\mathrm{Sp}}}{k_{\mathrm{Ic}}}\right) \int (G - G_{\mathrm{b}})dt + \frac{k_{\mathrm{Sen}} k_{\mathrm{r}} k_{\mathrm{Si}}}{k_{\mathrm{Ic}}} \int (G - G_{\mathrm{b}})_{+} t dt + \frac{k_{\mathrm{Sen}} k_{\mathrm{r}} k_{\mathrm{Sd}}}{k_{\mathrm{Ic}}} \int \dot{G}_{+} dt\,. \qquad \text{Eq. (49)}$$

If blood glucose rises once and does not rise again, Eq. (49) can be written as follows:

$$F = \boldsymbol{K} \cdot \boldsymbol{G}, \qquad \text{Eq. (50)}$$

where

$$F = (\text{integrated glucose input}),$$

$$\boldsymbol{K} = \begin{pmatrix} k_{\mathrm{Glub}} + \dfrac{k_{\mathrm{Sen}} k_{\mathrm{r}} k_{\mathrm{Sp}}}{k_{\mathrm{Ic}}} \\ \dfrac{k_{\mathrm{Sen}} k_{\mathrm{r}} k_{\mathrm{Si}}}{k_{\mathrm{Ic}}} \\ \dfrac{k_{\mathrm{Sen}} k_{\mathrm{r}} k_{\mathrm{Sd}}}{k_{\mathrm{Ic}}} \end{pmatrix},$$

$$\boldsymbol{G} = \begin{pmatrix} \int (G - G_{\mathrm{b}}) dt \\ \int (G - G_{\mathrm{b}})_{+} t dt \\ G_{\mathrm{max}} - G_{\mathrm{min}} \end{pmatrix}.$$

For readability, we denote the last term as $G_{\mathrm{max}} - G_{\mathrm{min}}$; however, if blood glucose first decreases and then increases again, $G_{\mathrm{max}} - G_{\mathrm{min}}$ should be replaced by $\int \dot{G_{+}} dt$. The values of the parameters in the above equations are not determined solely by pancreatic insulin secretion or glucose effectiveness. Rather, they are also influenced by the volumes in which glucose and insulin are distributed throughout the body. For example, individuals with a larger insulin distribution volume may have lower blood insulin concentrations, despite having high levels of insulin secretion from the pancreas. In principle, it is possible to estimate and correct for differences in distribution volume using measurable physiological indices, such as body weight [4,31]. However, for the sake of simplicity and because any estimation of distribution volume involves some degree of uncertainty, we choose not to perform such corrections in this derivation. Consequently, the parameters in Eqs. (47)-(50) are defined to include the effects of distribution volume.

Because $G_{\mathrm{Distortion}} = \int (G - G_{\mathrm{b}})_{+} t dt$ is an integral of cumulative exposure, it contains information that partly overlaps with $G_{\mathrm{AUC}} = \int (G - G_{\mathrm{b}}) dt$. Therefore, when glucose-dynamics features are used as predictors in regression or classification models, an AUC-normalized temporal feature may be considered to reduce the contribution of total exposure: $G_{\mathrm{Distortion}}^{\mathrm{norm}} = \frac{G_{\mathrm{Distortion}}}{G_{\mathrm{AUC}}}$. This normalized quantity reflects the temporal placement of glucose exposure relative to total exposure. When a dimensionless measure is desired, it can be further scaled by the length of the analysis window: $\tilde{G}_{\mathrm{Distortion}} = \frac{G_{\mathrm{Distortion}}}{(T - t_0) G_{\mathrm{AUC}}}$. Analyses using normalized distortion should account for cases in which $G_{\mathrm{AUC}}$ is close to zero, for example by excluding such challenges according to a prespecified threshold or by using an equivalent regularized denominator.

Eq. (50) provides a theoretical basis for estimating glucose-regulatory capacity (***K***) from measured glucose concentration profiles (***G***) together with the total administered glucose (F). Advances in CGM now enable continuous, minimally invasive monitoring of interstitial glucose. Furthermore, previous studies[4,31] have shown that plasma/blood glucose can be inferred from CGM using a first-order compartment model:

$$\dot{\mathrm{CGM}}(t) = -\frac{1}{\tau} \mathrm{CGM}(t) + \frac{1}{\tau} G(t), \qquad \text{Eq. (51)}$$

$$\mathrm{CGM}(0) = G_0, \qquad \text{Eq. (52)}$$

Here, τ represents the time constant for glucose equilibration between the intravascular and interstitial compartments. Collectively, these models enable the estimation of glucose regulation capacity without the need for blood sampling or insulin measurements, providing a less invasive approach to metabolic assessment.

Previous studies have also evaluated glucose-regulatory capacity using glucose data alone, without insulin measurements[3,4]. Two approaches have been proposed. One defines the disposition index (DI) as the mean glucose level[3], while the other defines DI as the product of insulin sensitivity and insulin secretion (β in Eq. (27)) [4]. Consequently, the latter approach qualitatively corresponds to intermediate forms between $\mathrm{DI}_{/\mathrm{clep}}$ and $\mathrm{DI}_{/\mathrm{clei}}$ (**Supplementary Note 1**). In the latter approach, additional assumptions were made (*e.g.*, setting the $k_{\mathrm{Sd}}$ term to zero). While these methods aim to reduce DI to a single scalar value, our results suggest that $\mathrm{DI}_{/\mathrm{clep}}$, $\mathrm{DI}_{/\mathrm{clei}}$, and $\mathrm{DI}_{/\mathrm{cled}}$ exhibit weak correlations and cannot be adequately condensed into a single latent dimension (**Figs. 1g–j** and **4c–e**). Accordingly, the three indices should be treated as distinct. Moreover, these three indices correspond to different features of the glucose waveform: mean, temporal distortion, and amplitude[42–46]. The distribution of individuals in the three-dimensional space spanned by these indices (**Fig. 1h–j**) provides a theoretical rationale for prior observations that mean, variability, and autocorrelation of glucose are independently associated with glucose-regulatory capacity and diabetes-related complications[42–46]. Finally, unlike DI definitions based solely on the product of insulin secretion and sensitivity, our framework integrates glucose dynamics, total glucose administration, and regulatory capacity into a single formulation that also considers glucose effectiveness.

When estimating the glucose regulation capacity ($\boldsymbol{K}$) using this method, the following four considerations should be noted. First, in our formulation $\boldsymbol{G}$ and $\boldsymbol{K}$ are three-dimensional vectors. In principle, glucose infusion data from at least three linearly independent administration patterns suffice to estimate $\boldsymbol{K}$ from $\boldsymbol{G}$ and F. In practice, however, collinearity among input patterns leads to ill-conditioned estimation and degrades stability and accuracy. More robust inference is obtained when using CGM data acquired under diverse glucose-administration conditions. In this study, we observed significant associations between estimated $\boldsymbol{K}$ and indices of glucose regulation and diabetes complications when CGM data were collected after >3 meals with distinct macronutrient compositions (**Fig. 2k–q**).

Second, the current model assumes that insulin sensitivity remains constant over time despite known fluctuations due to circadian rhythms[34]. To ensure consistency and comparability, CGM data should be collected at the same time of day (*e.g.*, morning) over several consecutive days. Additionally, meal intake prior to data collection and exercise may affect subsequent glucose dynamics and should be considered when designing data acquisition protocols.

Third, this mathematical model is based on a first-order approximation around an equilibrium point. Although the model has been validated under clamp conditions, where glucose and insulin levels increase markedly, the approximation may incur bias under conditions involving large-amplitude or rapid glucose excursions.

Fourth, estimates of $\boldsymbol{K}$ derived from clamp protocols may differ from those obtained during oral glucose tolerance tests or mixed-meal tolerance tests due to physiological factors, such as incretin effects and parasympathetic/autonomic activation. Nevertheless, the parameters, including $k_{\mathrm{Sd}}$ and $k_{\mathrm{Si}}$, are mathematically and qualitatively consistent with established models for oral glucose administration[4,34] (**Supplementary Note 1, Fig. 2a–c**), albeit with different absolute scales. Furthermore, disposition indices (DIs) estimated from OGTTs strongly correlated with DIs derived from clamp data (**Fig. 1p, q**). While a formal mathematical proof has yet to be established, the results in **Figure 2** provide empirical support for the model's predictive utility in assessing diabetes-related outcomes. Because ingested carbohydrate is not identical to systemic glucose appearance, the resulting CGM-derived $\boldsymbol{K}$ should be interpreted as an effective model-derived index that may also reflect gastric emptying, intestinal absorption, hepatic handling, and meal composition. Future work should directly compare CGM-based estimates of glucose-regulatory capacity with clamp-based measurements within the same individuals under harmonized protocols.

Equation $F = \boldsymbol{K} \cdot \boldsymbol{G}$ is an exact consequence of integrating the model between two steady states and is therefore a mathematical property of the model rather than an independent empirical hypothesis. Its empirical content lies in three separable claims, which we address in turn: (i) that the underlying model is an adequate description of glucose–insulin–C-peptide dynamics (**Supplementary Notes 3–11**); (ii) that $\boldsymbol{K}$ can in practice be recovered from $F$ and $\boldsymbol{G}$ alone, an inverse problem whose solvability does not follow from the derivation (**Fig. 2c, g**); and (iii) that the recovered $\boldsymbol{K}$ carries physiological and clinical information beyond conventional indices (**Figs. 1k–o, 2n–q**). Two of these analyses are informative precisely because they do not use the present model as the data-generating process. In the *in silico* analysis (**Fig. 2c**), the reference trajectories were generated by an independent meal simulation model of different structure; recovering a quantity that tracks the ground-truth VmxK of that model therefore tests whether the reduction retains the information it is intended to carry and cannot be satisfied by internal consistency alone. In the University of Tokyo Study (**Fig. 2g**), a single $\boldsymbol{K}$ was estimated per participant and held fixed across six glucose-challenge conditions differing in dose and ingestion rate; because the number of conditions exceeds the number of free components of $\boldsymbol{K}$, recovery of the administered dose is an overdetermined fit. The residual analysis in **Figure 2j** is of a different kind and is presented as a consistency check rather than as an independent test. It quantifies the cumulative magnitude of the approximations entering the derivation (the first-order approximation around equilibrium, the finite-window potentiation term, incomplete return to baseline, and the mapping from ingested to systemically appearing glucose) under conditions in which the total glucose appearance was constrained to the administered dose during model fitting.

#### 13. PID-like insulin secretion and multidimensional glucose dynamics

We next examined whether the insulin secretion component of the model required proportional, integral-like, and derivative-like terms. We compared the full model with reduced models using serum insulin trajectories measured during the hyperglycemic clamp in the *Kobe University Study*. In the full model, glucose-stimulated insulin secretion was represented as a PID-like process comprising proportional, integral-like, and derivative-like components, $\dot{I} = k_{\mathrm{r}} k_{\mathrm{Sp}} G + k_{\mathrm{r}} k_{\mathrm{Si}} (G - G_{\mathrm{b}})_{+} t + k_{\mathrm{r}} k_{\mathrm{Sd}} \dot{G}_{+} - k_{\mathrm{Ic}} I$, where the derivative-like term was asymmetric and was activated only when plasma glucose was increasing. We fitted the full model containing all three components and an asymmetric derivative input (PID+), as well as reduced models lacking the proportional, integral-like, or derivative-like component, and a symmetric derivative model in which $\dot{G}_{+}$ was replaced by the signed derivative $\dot{G}$.

The full PID+ model reproduced the measured insulin trajectory during the clamp, whereas model performance deteriorated when any of the three components was removed (**Extended Data Fig. 4a, b**). Removing the proportional term impaired prediction of the early insulin response, removing the integral-like term worsened prediction during the later clamp phase, and removing the derivative-like term reduced accuracy after the initial glucose elevation. Replacing the asymmetric derivative term with a signed derivative also reduced model performance, particularly during the phase around 100 min when plasma glucose declined and insulin concentrations decreased. Thus, insulin dynamics during the hyperglycemic clamp were best captured by a PID-like secretion model that included all three components and an asymmetric derivative term.

We then asked whether glucose input could be inferred from a one-dimensional representation based only on glucose area under the curve, $F = K_{\mathrm{AUC}} \cdot G_{\mathrm{AUC}}$, or whether a multidimensional description of glucose dynamics ($\boldsymbol{G}$) was required. Using the mathematical model developed in this study and mean parameter values estimated from NGT participants in the *Kobe University Study*, we simulated glucose and insulin dynamics while holding the individual-level regulatory capacity and total glucose input constant but varying only the duration of glucose delivery (**Extended Data Fig. 4c, d**). Shorter input durations produced sharper glucose excursions, whereas longer input durations produced lower and more prolonged profiles. Despite identical total glucose input ($F$) in the same simulated individual ($K$), $G_{\mathrm{AUC}}$ changed with input duration, causing the apparent scalar capacity $F/G_{\mathrm{AUC}}$ to vary across conditions (**Extended Data Fig. 4e, f**). Thus, glucose AUC alone did not provide an invariant measure of glucose-regulatory capacity.

Consistent with this limitation, prediction of administered glucose from the mean scalar capacity $F/G_{\mathrm{AUC}}$ produced variable estimates across glucose waveforms, whereas the multidimensional equation $F = \boldsymbol{K} \cdot \boldsymbol{G}$ recovered the administered glucose input from waveform features (**Extended Data Fig. 4g**). We systematically tested this difference across 40 simulated conditions combining five total glucose inputs (15, 25, 35, 45, and 55 mmol/L) and eight input durations (3, 5, 10, 20, 30, 60, 120, and 240 min). The AUC-only model predicted administered glucose with substantial error (R=0.794, AIC = 189.4), whereas the three-dimensional formulation recovered the input nearly exactly (R=1.000, AIC = −447.6) (**Extended Data Fig. 4h**).

We further tested whether $G_{\mathrm{AUC}}$ uniquely determines the shape of the glucose waveform within a physiological parameter range. In these simulations, the total administered glucose dose ($F$) and the duration of administration were held identical, and only the glucose-regulatory capacity ($\boldsymbol{K}$) was varied. With parameters sampled from within the interquartile range of NGT participants in the *Kobe University Study*, glucose waveforms with nearly identical $G_{\mathrm{AUC}}$ nonetheless differed markedly in amplitude and in temporal distortion (**Extended Data Fig. 4i–n**). Waveforms with high amplitude or high distortion were associated with lower components of $\boldsymbol{K}$, despite comparable $G_{\mathrm{AUC}}$. Thus, $G_{\mathrm{AUC}}$ does not uniquely determine either glucose waveform structure or the underlying glucose-regulatory capacity. These simulations support modeling glucose homeostasis using the three-dimensional equation $F = \boldsymbol{K} \cdot \boldsymbol{G}$, rather than a one-dimensional AUC-based formulation.

14. Estimation of insulin-independent and -dependent glucose-lowering effects

We estimated the relative contributions of the insulin-independent and insulin-dependent effects of lowering glucose under steady-state and dynamic conditions. Under steady state conditions, Eqs. (37) and (38) can be rewritten as follows:

$$f_{\mathrm{G}} = k_{\mathrm{Glub}}(G - G_{\mathrm{b}}) + k_{\mathrm{Sen}}(I - I_{\mathrm{b}}), \qquad \text{Eq. (53)}$$

where $f_{\mathrm{G}}$ denotes the glucose infusion rate between 60 and 90 minutes during the clamp tests. We defined the steady-state insulin-independent glucose-lowering effect as $k_{\mathrm{Glub}}(G - G_{\mathrm{b}})$ and the steady-state insulin-dependent effect as $k_{\mathrm{Sen}}(I - I_{\mathrm{b}})$.

Eqs. (44), (45), and (46) can be rearranged as follows:

$$F_{\mathrm{G0-G5}} = (G_5 - G_0) + k_{\mathrm{Glub}} \int (G - G_0) dt + (Q_5 - Q_0) + \int X dt, \qquad \text{Eq. (54)}$$

where $F_{\mathrm{G0-G5}}$ corresponds to the total amount of glucose infused during the first five minutes of the clamp tests. We defined the dynamic insulin-independent glucose-lowering effect as $(G_5 - G_0) + k_{\mathrm{Glub}} \int (G - G_0) dt + (Q_5 - Q_0)$ and the dynamic insulin-dependent glucose-lowering effect as $\int X dt$.

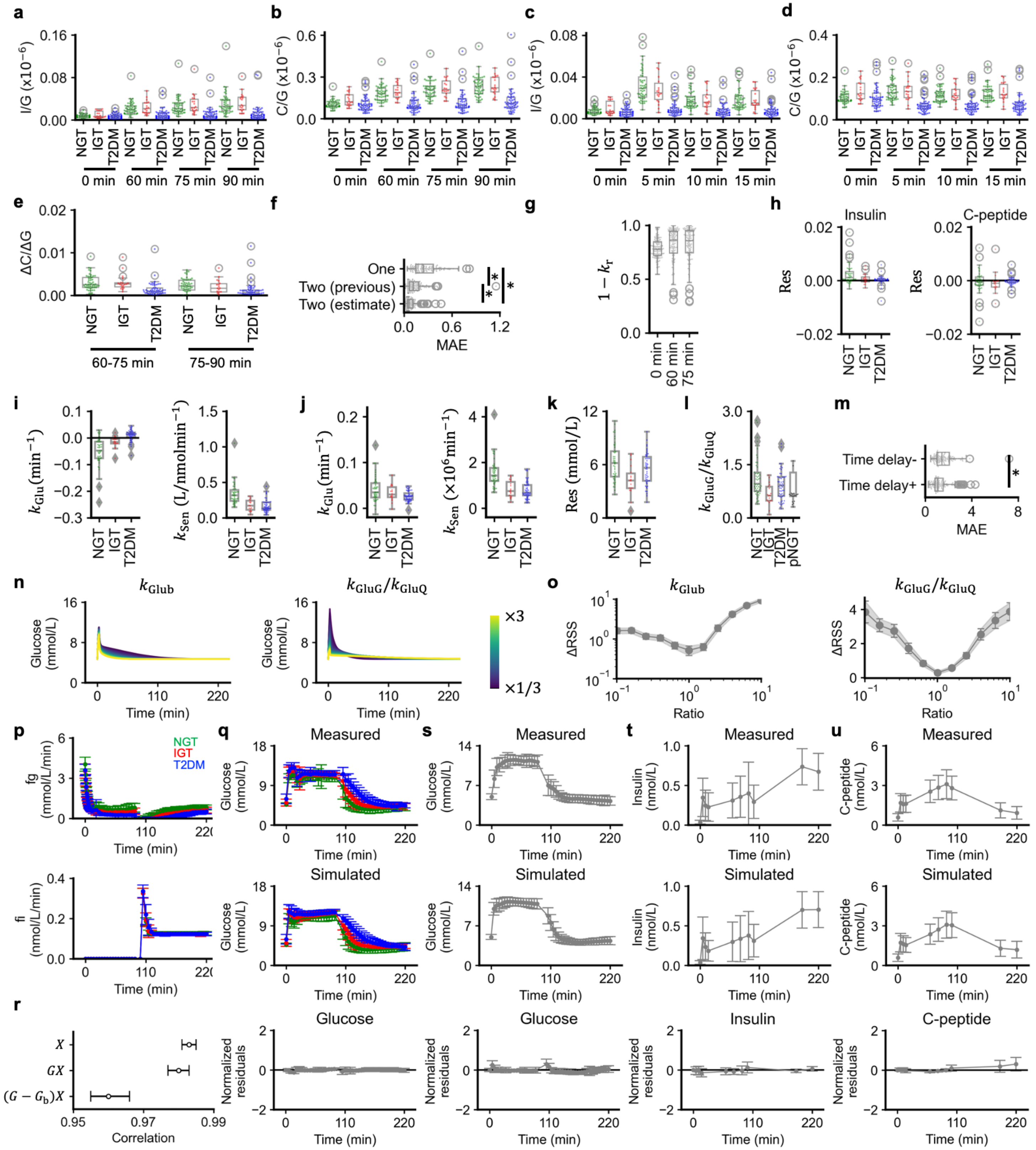

**Extended Data Fig. 1 | Estimation of glucose-insulin regulatory parameters using clamp data in NGT, IGT and T2DM.**

**a-d,** Box plots of insulin-to-glucose (I/G) and C-peptide-to-glucose (C/G) ratios at indicated time points during hyperglycemic clamp tests in individuals with normal glucose tolerance (NGT), impaired glucose tolerance (IGT), and type 2 diabetes mellitus (T2DM).
**e,** Box plots of the first-order differences (ΔC/ΔG) between 60-90 minutes.
**f,** Box plots of the mean absolute error (MAE) of three different models to simulate C-peptide dynamics: one-compartment model (One), two-compartment model with externally estimated parameters (Two (previous)) and two-compartment model with fitted parameters (Two (estimate)). $*Q < 0.05$.
**g,** Box plots of estimated hepatic insulin clearance ($1 - k_r$) at 0, 60, and 75 min during hyperglycemic clamp tests.
**h,** Box plots of the right-hand sides of Eqs. (24) and (25) in NGT, IGT, and T2DM.
**i, j,** Box plots of estimated glucose effectiveness ($k_{\text{Glu}}$) and insulin sensitivity ($k_{\text{Sen}}$) based on Eq. (30) (**i**) and Eq. (33) (**j**).
**k,** Residual differences between the left and right sides of Eq. (36) (Res).
**l,** Box plots of $k_{\text{GluG}}/k_{\text{GluQ}}$ in NGT, IGT, T2DM, and previously studied NGT (pNGT) individuals [22].
**m,** Box plots of MAE for models with or without an insulin-action delay (Time delay (+) vs Time delay (−)). $*P < 0.05$. Box plots show median (center line) and interquartile range (box).
**n,** Simulated plasma glucose trajectories generated using a two-compartment model of glucose kinetics during an intravenous glucose tolerance test. Values of $k_{\text{Glub}}$ and $k_{\text{GluG}}/k_{\text{GluQ}}$ were varied from one-third to three times the mean value in NGT individuals.
**o,** Profile residual sum of squares (RSS) analysis for assessing the identifiability of $k_{\text{Glub}}$ and $k_{\text{GluG}}/k_{\text{GluQ}}$. Each parameter was fixed between 0.1× and 10× its best-fit value while the others were re-optimized; ΔRSS relative to the minimum is plotted against the fixed-parameter ratio. Points and shaded areas, mean and 95% CI across individuals (n = 121).
**p,** Time course of the administered glucose rate (fg) and insulin rate (fi) during sequential hyperglycemic and hyperinsulinemic–euglycemic clamp tests in the *Kobe University Study* (n = 121). Data are mean ± SD for individuals with normal glucose tolerance (NGT, green), impaired glucose tolerance (IGT, red), and type 2 diabetes mellitus (T2DM, blue).
**q,** Validation of the mathematical model in the *Kobe University Study* (n = 121). Plasma-glucose time courses during the sequential clamp. Sub-panels show measured concentrations (top), concentrations simulated with individually estimated parameters (middle), and normalized residuals [(simulated − measured)/measured] (bottom). Data are mean ± SD, colored by group.
**r,** Mean and 95% CI of the correlation coefficients between measured and simulated glucose trajectories during the clamp tests. $X, GX, (G - G_b)X$ denote alternative formulations of the insulin-dependent glucose-lowering term in the model.
**s–u,** Validation of the mathematical model using an independent clamp dataset from the *Kobe University Study2* (n = 64). Time courses of plasma glucose (**s**), serum insulin (**t**), and serum C-peptide (**u**) during sequential hyperglycemic and hyperinsulinemic–euglycemic clamp tests. For each variable, the upper sub-panel shows the measured concentrations, the middle sub-panel shows the concentrations simulated with individually estimated parameters, and the lower sub-panel shows the normalized residuals. Data are mean ± SD.

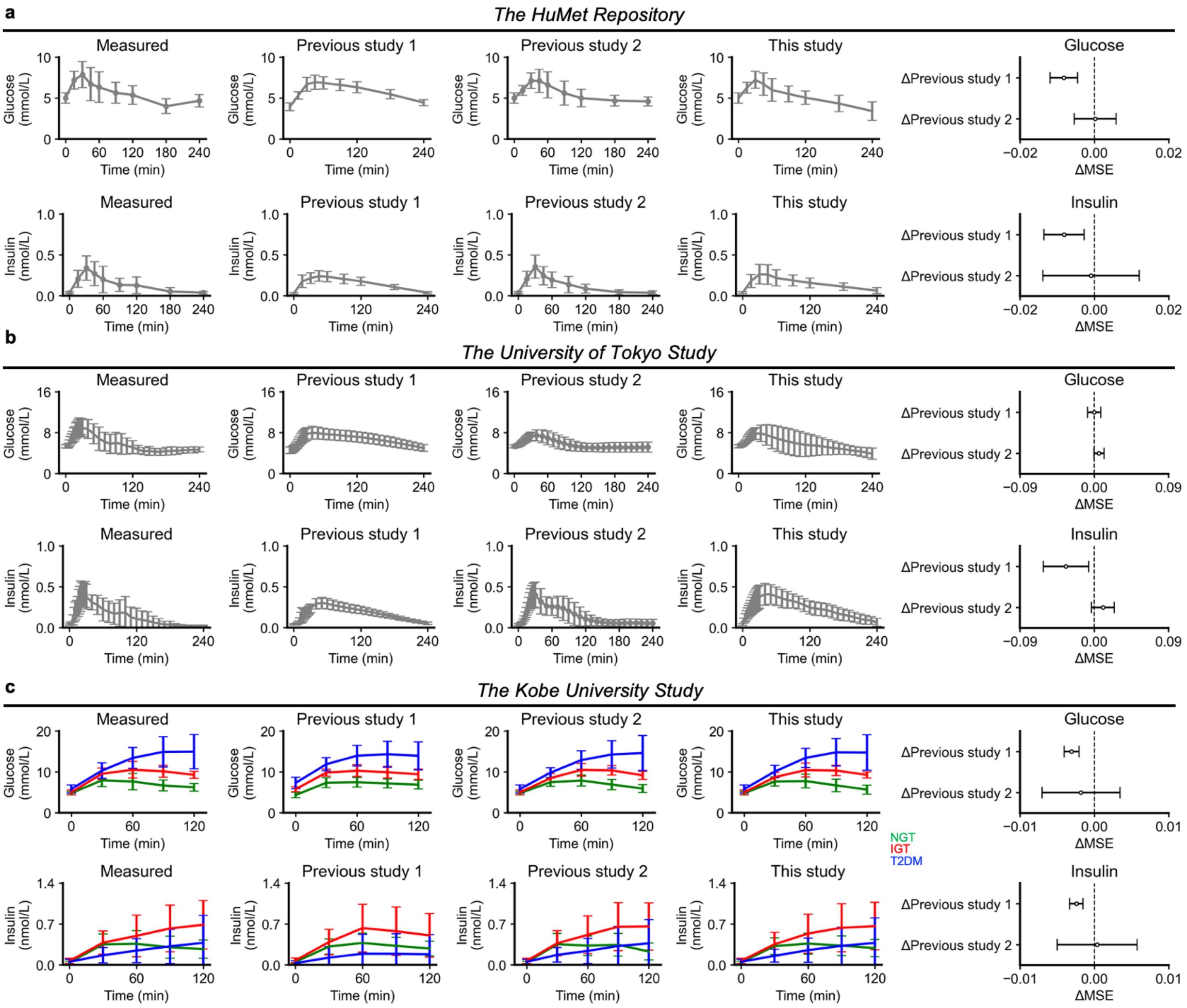


**Extended Data Fig. 2 | Validation of the mathematical model using OGTT data.**
**a,** Validation using 75 g OGTT data from the *HuMet Repository*. The upper row shows plasma glucose concentrations, and the lower row shows serum insulin concentrations. Measured data are shown together with simulations from Previous study 1, Previous study 2, and the model developed in this study. Previous study 1 corresponds to the insulin secretion and sensitivity model of Ha et al.; Previous study 2 corresponds to the Dalla Man meal simulation model; This study corresponds to the simplified glucose-insulin model developed in this study. Data are mean ± SD across participants. Right panels show paired differences in mean squared error (MSE) between This study and each previous model. ΔMSE was calculated as MSE (This study)−MSE (Previous study). Negative values indicate lower error for the model used in this study. Points indicate mean differences, and horizontal bars indicate 95% confidence intervals.
**b,** Validation using OGTT data from the *University of Tokyo Study*. Participants received 25, 50, or 75 g of glucose by bolus ingestion. The upper row shows plasma glucose concentrations, and the lower row shows serum insulin concentrations. Measured trajectories are shown together with simulations from Previous study 1,

Previous study 2, and This study. Data are mean ± SD across glucose challenge profiles. Right panels show ΔMSE for glucose and insulin.
**c,** Validation using 75 g OGTT data from the *Kobe University Study*. The analysis included 121 individuals classified as NGT, IGT, or T2DM. The upper row shows plasma glucose concentrations, and the lower row shows serum insulin concentrations. Measured and simulated trajectories are shown separately by glucose tolerance group: NGT, green; IGT, red; T2DM, blue. Data are mean ± SD within each group. Right panels show ΔMSE for glucose and insulin.

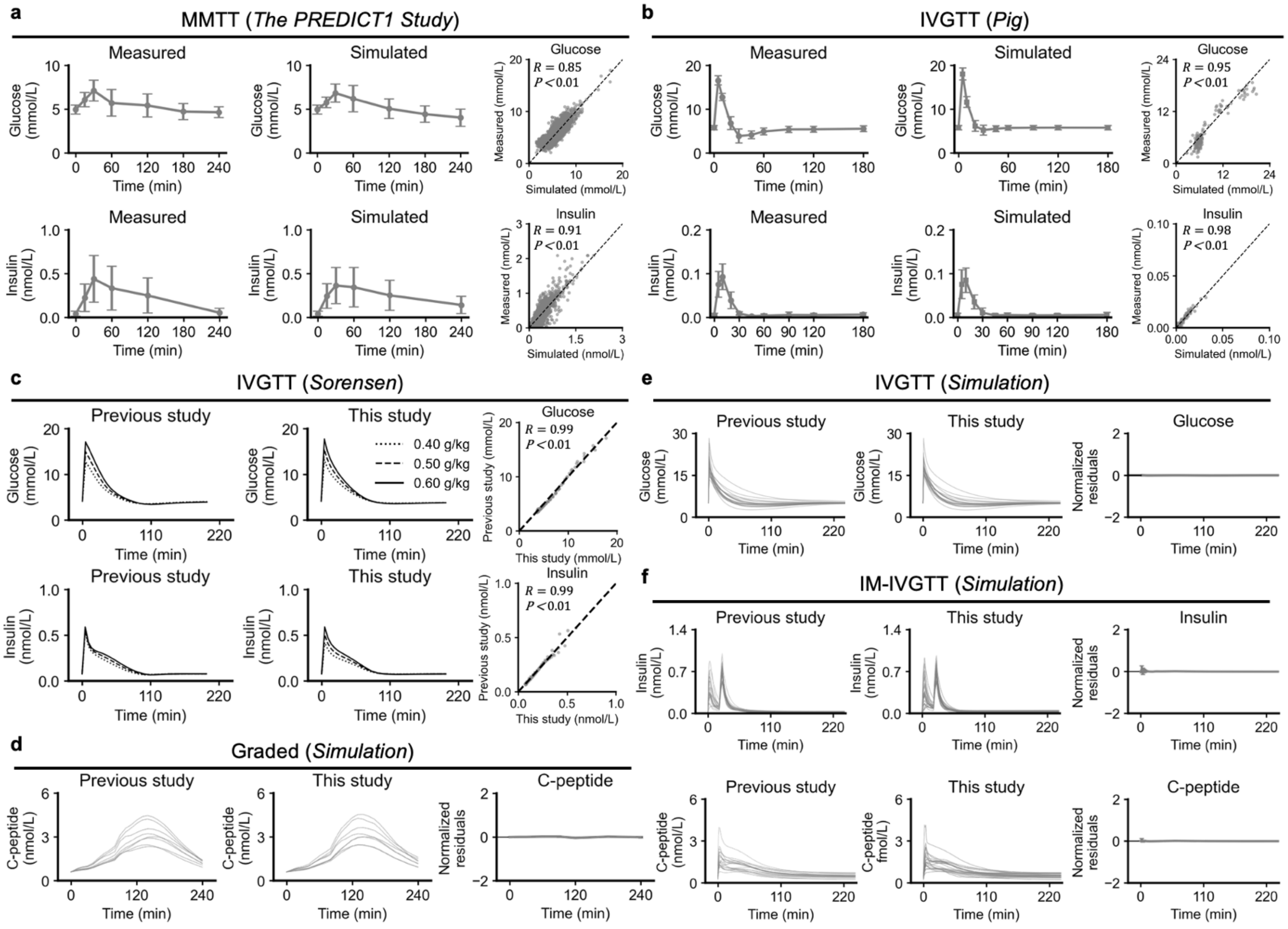


**Extended Data Fig. 3 | Validation of the mathematical model across heterogeneous experimental and simulated datasets.**

**a,** Validation using mixed-meal tolerance test (MMTT) data from the *PREDICT1 Study*. The analysis included 903 participants who underwent an MMTT with an administered glucose-equivalent carbohydrate load of 86 g. The mathematical model was fitted separately to each participant's glucose and insulin time courses by optimizing participant-specific parameters to minimize normalized residuals between measured and simulated concentrations. Time-course panels show measured and simulated plasma glucose and serum insulin concentrations. Data are mean ± SD across participants. Scatter plots compare measured and simulated concentrations across all participants and time points. Each point represents one measured–simulated pair from one participant at one sampling time point. Pearson's correlation coefficients are shown. The dashed diagonal line indicates identity.

**b,** Validation using intravenous glucose tolerance test (IVGTT) data from pigs. Eighteen pigs received intravenous glucose at 0.5 g/kg body weight. Time-course panels show measured and simulated plasma glucose and serum insulin concentrations. Data are mean ± SD across pigs. Scatter plots compare measured and simulated concentrations across all pigs and time points. Pearson's correlation coefficients are shown. The dashed diagonal line indicates identity.

**c,** Glucose and insulin trajectories generated by the Sorensen model and fitted trajectories from the simplified model developed in this study. The Sorensen model was used to simulate intravenous glucose tolerance tests with glucose doses of 0.4, 0.5, and 0.6 g/kg in a 70 kg individual. Glucose was administered as a rectangular

intravenous input over the first 3 min, and the resulting Sorensen-model trajectories were treated as reference trajectories. A single set of model parameters was fitted simultaneously to all three Sorensen-generated IVGTT trajectories by minimizing normalized glucose and insulin residuals. Line styles indicate glucose dose: dotted, 0.4 g/kg; dashed, 0.5 g/kg; solid, 0.6 g/kg. The left panels show Sorensen-model trajectories, the middle panels show trajectories simulated by the fitted simplified model, and the right panels compare fitted values with Sorensen-model reference values across all glucose doses and time points. Each point represents one dose–time pair. The dashed diagonal line indicates identity. Pearson's correlation coefficients are shown.
**d,** Validation of the C-peptide component of the mathematical model using graded glucose infusion data. Reference C-peptide trajectories were generated using the model formulation of Toffolo et al. for eight individuals. The graded glucose infusion protocol consisted of seven consecutive 40-min periods with glucose infusion rates of 0, 4, 8, 16, 8, 4, and 0 mg $kg^{-1}$ $min^{-1}$. The mean plasma glucose trajectory from the reference study was used as the glucose input. Participant-specific parameters were set to the values reported by Toffolo et al., and the two-compartment C-peptide kinetic parameters were used. The panel labeled "Previous study" shows C-peptide trajectories simulated by the Toffolo model. The panel labeled "This study" shows fitted C-peptide trajectories generated by the C-peptide model developed in this study. Model parameters were fitted separately for each individual by minimizing normalized residuals between Toffolo-generated and fitted C-peptide concentrations.
**e,** Validation of the glucose component of the mathematical model using IVGTT trajectories generated by the two-compartment model of Cobelli et al. The reference model simulated plasma glucose dynamics after a standard intravenous glucose tolerance test with a glucose dose of approximately 0.30 g/kg. The glucose bolus was modeled as a rectangular intravenous infusion, and the insulin time course was used as an external input. The panel labeled "Previous study" shows glucose trajectories simulated by the Cobelli two-compartment model. The panel labeled "This study" shows the corresponding trajectories fitted by the glucose subsystem of the model developed in this study. The residual panel shows normalized residuals; the horizontal line indicates zero.
**f,** Validation of the insulin and C-peptide components of the mathematical model using insulin-modified IVGTT (IM-IVGTT) trajectories generated by the models of Toffolo et al. The reference simulation was based on an insulin-modified intravenous glucose tolerance test in which glucose was administered intravenously at 0.3 g/kg, followed by exogenous insulin infusion from 20 to 25 min at a total dose of 0.02 U/kg. The reference model generated insulin and C-peptide trajectories using a C-peptide model of insulin secretion and kinetics and an insulin delivery and kinetics model. The panels labeled "Previous study" show insulin and C-peptide trajectories simulated by the Toffolo model. The panels labeled "This study" show trajectories fitted by the insulin–C-peptide subsystem of the model developed in this study. The residual panels show normalized residuals; horizontal lines indicate zero.

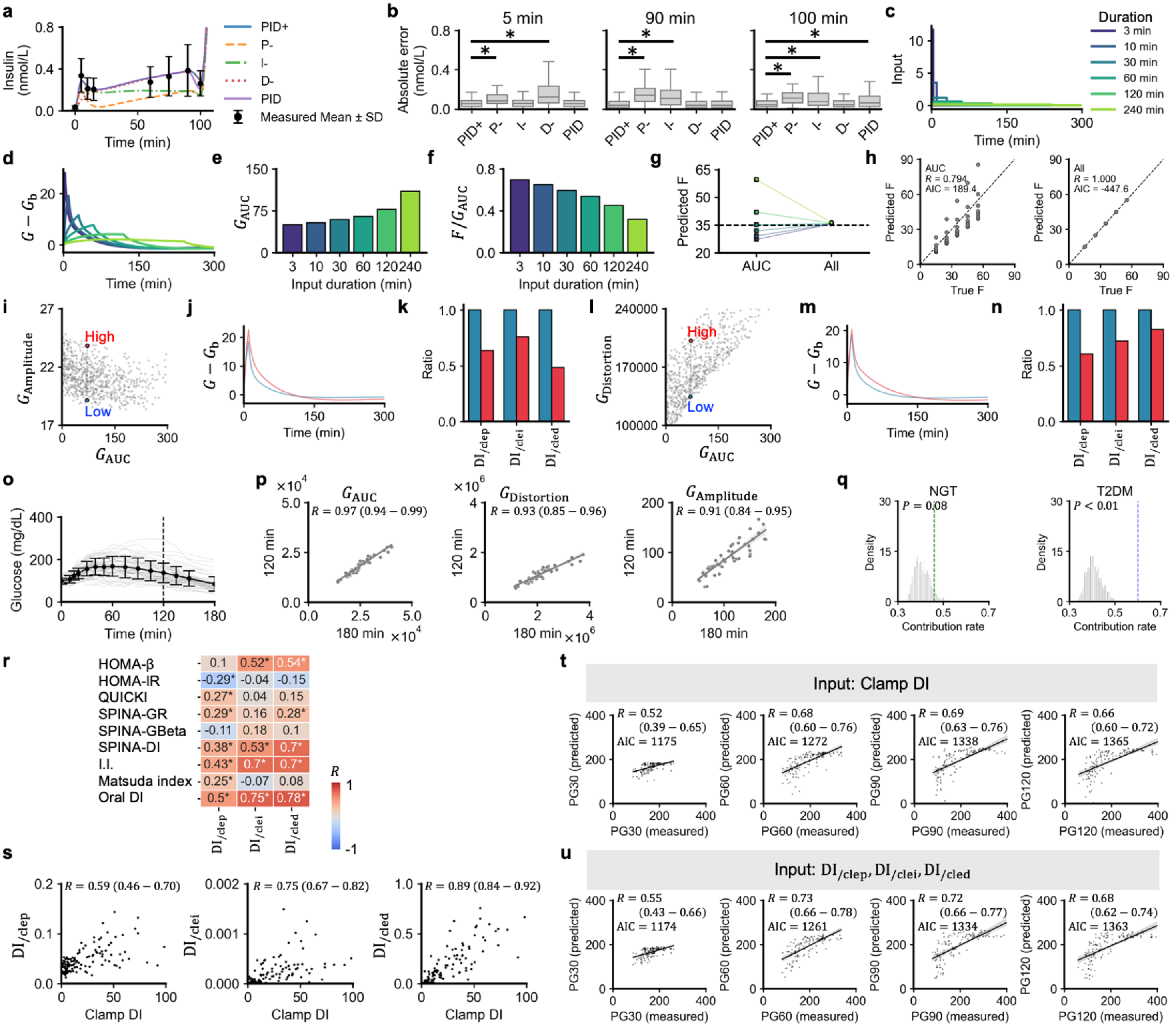


**Extended Data Fig. 4 | Model-structure validation of the three-dimensional representation of glucose control abilities.**

**a,** Simulated serum insulin trajectories during the hyperglycemic clamp in the *Kobe University Study*. Measured insulin concentrations are shown as mean ± SD. The full model (PID+) includes proportional, integral-like, and asymmetric derivative-like insulin secretion components. Reduced models remove the proportional (P−), integral-like (I−), or the derivative-like (D−) component, or replace the asymmetric derivative term with a symmetric signed derivative term (PID).

**b,** Box plots showing the absolute insulin prediction error at representative time points during the clamp test. Asterisks indicate significant differences from the full PID+ model ($Q < 0.05$).

**c,** Simulated glucose input profiles with identical total glucose input $F$ = 35 mmol/L but different input durations. Durations were 3, 10, 30, 60, 120, and 240 min.

**d,** Plasma glucose excursions ($G–G_b$) generated by the input profiles in (c). The same model parameters and the same total input $F$ were used for all curves; only the duration of glucose input was varied.

**e,** $G_{\text{AUC}}$ calculated from the simulated glucose trajectories in (d).
**f,** Apparent one-dimensional glucose-regulatory capacity $F/G_{\text{AUC}}$ for each input duration.
**g,** Predicted glucose input $F$ using either an AUC-only model ($F = K \cdot G_{\text{AUC}}$) or the full three-dimensional model ($F = \boldsymbol{K} \cdot \boldsymbol{G}$). The dashed line indicates the true input F = 35 mmol/L.
**h,** True versus predicted glucose input $F$ across 40 simulations (5 amounts: 15, 25, 35, 45, 55 mmol/L × 8 durations: 3, 5, 10, 20, 30, 60, 120, 240 min) for the AUC-only model (left) and the full three-dimensional model (right). The dashed line indicates identity.
**i,** Simulations within the interquartile range of NGT parameter values showing that $G_{\text{AUC}}$ does not uniquely determine $G_{\text{Amplitude}}$. Gray points represent 800 individuals generated by uniformly sampling model parameters within the NGT interquartile range. Colored points highlight two simulations with similar $G_{\text{AUC}}$ but different $G_{\text{Amplitude}}$.
**j,** Glucose waveforms for the low- and high-amplitude simulations highlighted in (i). The two waveforms had similar $G_{\text{AUC}}$ but different $G_{\text{Amplitude}}$.
**k,** Relative $\boldsymbol{K}$ components for the high-amplitude waveform (red) compared with the low-amplitude waveform (blue). Bars show ratios of $\text{DI}_{/\text{clep}}$, $\text{DI}_{/\text{clei}}$, and $\text{DI}_{/\text{cled}}$.
**l,** Simulations within the NGT interquartile parameter range showing that $G_{\text{AUC}}$ does not uniquely determine $G_{\text{Distortion}}$. Colored points highlight two simulations with similar $G_{\text{AUC}}$ but different $G_{\text{Distortion}}$.
**m,** Glucose waveforms for the low- and high-distortion simulations highlighted in (l). The two waveforms had similar $G_{\text{AUC}}$ but distinct $G_{\text{Distortion}}$.
**n,** Relative $\boldsymbol{K}$ components for the high-distortion waveform compared with the low-distortion waveform. Bars show ratios of $\text{DI}_{/\text{clep}}$, $\text{DI}_{/\text{clei}}$, and $\text{DI}_{/\text{cled}}$. Higher $G_{\text{Distortion}}$ despite similar $G_{\text{AUC}}$ was associated with lower estimated glucose-regulatory capacity components.
**o,** Time courses of plasma glucose concentrations after the oral administration of 75 g glucose (n = 51). Data are mean ± SD.
**p,** Scatter plots of glucose dynamics-derived indices ($G_{\text{AUC}}$, $G_{\text{Distortion}}$, and $G_{\text{Amplitude}}$) calculated from 0–120 min versus 0–180 min post-glucose load. Spearman's correlation coefficient ($R$) and its 95% confidence intervals (in parentheses) are shown.
**q,** Null distribution of the PC1 contribution rate underlying Figure 1j, for NGT and T2DM.
**r,** Heatmap of Spearman's correlations between conventional metabolic indices and DI/cle components in the *Kobe University Study*. Rows indicate HOMA-β, HOMA-IR, QUICKI, SPINA-GR, SPINA-GBeta, SPINA-DI, insulinogenic index (I.I.), Matsuda index, and oral DI. Columns indicate DI/clep, DI/clei, and DI/cled. Values indicate Spearman's correlation coefficients. Asterisks indicate Q < 0.05 after Benjamini-Hochberg correction.
**s,** Scatter plots comparing clamp DI with $\text{DI}_{/\text{clep}}$, $\text{DI}_{/\text{clei}}$, and $\text{DI}_{/\text{cled}}$.
**t,** Measured versus predicted plasma glucose concentrations from linear regression using clamp DI as a predictor.
**u,** Measured versus predicted plasma glucose concentrations from linear regression using $\text{DI}_{/\text{clep}}$, $\text{DI}_{/\text{clei}}$, and $\text{DI}_{/\text{cled}}$ as predictors. AIC denotes Akaike Information Criterion.

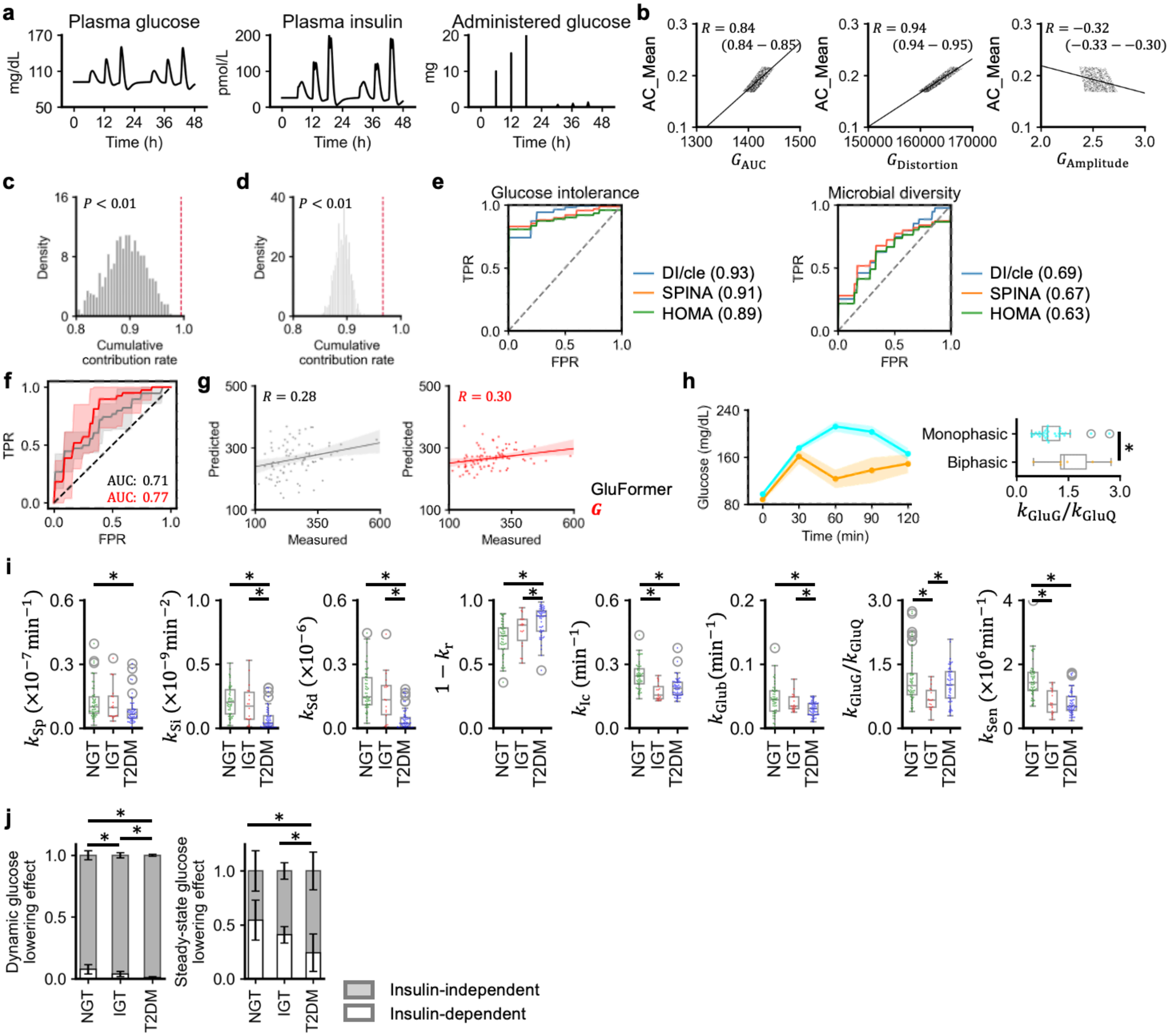


**Extended Data Fig. 5 | Additional model simulations and external validation of the glucose-control representation.**

**a,** Examples of simulated plasma glucose and insulin after different glucose loads (20, 30, 40 g) and temporal patterns (bolus, continuous).

**b,** Scatter plots of AC_Mean against $G_{\mathrm{AUC}}$, $G_{\mathrm{Distortion}}$ and $G_{\mathrm{Amplitude}}$. As in Figure 2c, 1,000 randomly subsampled individuals are displayed for clarity, while all correlation coefficients and 95% confidence intervals were computed on the full simulated dataset (n = 10,000).

**c,** Null distribution of the cumulative PC1 + PC2 contribution rate underlying Figure 2f. The statistic was computed by performing PCA independently for each of n = 3 participants on a 6-condition × 3-feature $\boldsymbol{G}/F$ matrix and averaging the per-participant PC1 + PC2 cumulative contribution rates. The gray histogram shows the null distribution, normalized to probability density, generated from 1000 synthetic datasets. Empirical value = 0.995 (crimson dashed line).

**d,** Null distribution of the cumulative PC1 + PC2 contribution rate underlying Figure 2m. The statistic was computed as in (c) but for n = 22 participants. Empirical value = 0.967 (crimson dashed line).
**e,** Receiver operating characteristic (ROC) curves for classification of glucose intolerance and active microbial diversity using logistic regression models based on $\mathrm{DI}_{/\mathrm{cle}}$ ($\mathrm{DI}_{/\mathrm{clep}}$, $\mathrm{DI}_{/\mathrm{clei}}$, and $\mathrm{DI}_{/\mathrm{cled}}$), SPINA (SPINA-GBeta, SPINA-GR, and SPINA-DI), or HOMA/QUICKI (HOMA-β, HOMA-IR, and QUICKI) feature sets. AUC values are shown in parentheses. Data are from the *CGMacros dataset* (n = 45).
**f,** ROC curves for predicting the presence of diabetic complications from continuous glucose monitoring data. Black, GluFormer; red, prediction based on glucose dynamics–derived indices ($\boldsymbol{G}$). Shaded areas indicate 95% confidence intervals. AUC, area under the ROC curve. Data are from the *Shanghai Study* (n = 100).
**g,** Scatter plots of measured versus predicted plasma glucose concentration at 2 hours after glucose administration. R, Pearson's correlation coefficient.
**h,** Time courses of plasma glucose levels during OGTT in individuals with monophasic and biphasic glucose responses. Data are mean ± SEM. Box plots comparing estimated $k_{\mathrm{GluG}}/k_{\mathrm{GluQ}}$ between individuals with monophasic and biphasic glucose responses. $^{*}P < 0.05$.
**i,** Box plots of estimated parameters for NGT, IGT, and T2DM individuals: basal insulin secretion ($k_{\mathrm{Sp}}$), potentiation factor of insulin secretion ($k_{\mathrm{Si}}$), first-phase insulin secretion ($k_{\mathrm{Sd}}$), hepatic insulin clearance ($1 - k_{\mathrm{r}}$), peripheral insulin clearance ($k_{\mathrm{Ic}}$), glucose effectiveness ($k_{\mathrm{Glub}}$), ratio of rate parameters describing glucose exchange kinetics ($k_{\mathrm{GluG}}/k_{\mathrm{GluQ}}$), and insulin sensitivity ($k_{\mathrm{Sen}}$). The boxes denote the median and upper and lower quartiles.
**j,** Relative contributions of insulin-independent versus insulin-dependent glucose-lowering. Data are mean ± SD. $^{*}Q < 0.05$. $Q$ values were calculated using Welch's *t*-tests with Benjamini–Hochberg correction.

#### Supplementary References


1. Mari, A., Tura, A., Gastaldelli, A. & Ferrannini, E. Assessing insulin secretion by modeling in multiple-meal tests: role of potentiation. *Diabetes* **51 Suppl 1**, S221-6 (2002).
2. Watson, E. M., Chappell, M. J., Ducrozet, F., Poucher, S. M. & Yates, J. W. T. A new general glucose homeostatic model using a proportional-integral-derivative controller. *Comput. Methods Programs Biomed.* **102**, 119–129 (2011).
3. Ha, J. *et al.* A mathematical model-derived disposition index without insulin validated in youth with obesity. *J. Clin. Endocrinol. Metab.* **110**, 706–714 (2025).
4. Schiavon, M., Vella, A. & Dalla Man, C. Quantitative estimation of disposition Index from postprandial glucose data across the spectrum of glucose tolerance. *Am. J. Physiol. Endocrinol. Metab.* https://doi.org/10.1152/ajpendo.00407.2024 (2025) doi:10.1152/ajpendo.00407.2024.
5. Piccinini, F. & Bergman, R. N. The Measurement of Insulin Clearance. *Diabetes Care* **43**, 2296–2302 (2020).
6. Van Cauter, E., Mestrez, F., Sturis, J. & Polonsky, K. S. Estimation of insulin secretion rates from C-peptide levels. Comparison of individual and standard kinetic parameters for C-peptide clearance. *Diabetes* **41**, 368–377 (1992).
7. Gastaldelli, A., Abdul Ghani, M. & DeFronzo, R. A. Adaptation of Insulin Clearance to Metabolic Demand Is a Key Determinant of Glucose Tolerance. *Diabetes* **70**, 377–385 (2021).
8. De Gaetano, A. & Arino, O. Mathematical modelling of the intravenous glucose tolerance test. *J. Math. Biol.* **40**, 136–168 (2000).
9. Campioni, M., Toffolo, G., Basu, R., Rizza, R. A. & Cobelli, C. Minimal model assessment of hepatic insulin extraction during an oral test from standard insulin kinetic parameters. *Am. J. Physiol. Endocrinol. Metab.* **297**, E941-8 (2009).
10. Toffolo, G. *et al.* Quantitative indexes of beta-cell function during graded up&down glucose infusion from C-peptide minimal models. *Am. J. Physiol. Endocrinol. Metab.* **280**, E2-10 (2001).
11. Henquin, J. C. Triggering and amplifying pathways of regulation of insulin secretion by glucose. *Diabetes* **49**, 1751–1760 (2000).
12. Rorsman, P. & Renström, E. Insulin granule dynamics in pancreatic beta cells. *Diabetologia* **46**, 1029–1045 (2003).
13. Fridlyand, L. E. & Philipson, L. H. Glucose sensing in the pancreatic beta cell: a computational systems analysis. *Theor. Biol. Med. Model.* **7**, 15 (2010).
14. Chen, Y., Wang, S. & Sherman, A. Identifying the targets of the amplifying pathway for insulin secretion in pancreatic beta-cells by kinetic modeling of granule exocytosis. *Biophys. J.* **95**, 2226–2241 (2008).
15. Pedersen, M. G., Tagliavini, A. & Henquin, J.-C. Calcium signaling and secretory granule pool dynamics underlie biphasic insulin secretion and its amplification by glucose: experiments and modeling. *Am. J. Physiol. Endocrinol. Metab.* **316**, E475–E486 (2019).
16. Deepa Maheshvare, M., Raha, S., König, M. & Pal, D. A pathway model of glucose-stimulated insulin secretion in the pancreatic β-cell. *Front. Endocrinol. (Lausanne)* **14**, 1185656 (2023).
17. Mari, A. *et al.* Meal and oral glucose tests for assessment of β-cell function: modeling analysis in normal subjects. *American Journal of Physiology-Endocrinology and Metabolism* **283**, E1159–E1166 (2002).

18. Takeda, Y. *et al.* Systems analysis of GLP-1 receptor signaling in pancreatic β-cells. *Am. J. Physiol. Cell Physiol.* **301**, C792-803 (2011).
19. Grodsky, G. M. A threshold distribution hypothesis for packet storage of insulin and its mathematical modeling. *J. Clin. Invest.* **51**, 2047–2059 (1972).
20. Hashiuchi, E. *et al.* Vagal activation inhibits insulin release through neuronal nitric oxide synthase in obese male mice. *Sci. Signal.* **19**, eadz8805 (2026).
21. Bergman, R. N. Origins and History of the Minimal Model of Glucose Regulation. *Front. Endocrinol.* **11**, 583016 (2020).
22. Cobelli, C., Caumo, A. & Omenetto, M. Minimal model SG overestimation and SI underestimation: improved accuracy by a Bayesian two-compartment model. *Am. J. Physiol.* **277**, E481-8 (1999).
23. Raue, A. *et al.* Structural and practical identifiability analysis of partially observed dynamical models by exploiting the profile likelihood. *Bioinformatics* **25**, 1923–1929 (2009).
24. Maiwald, T. *et al.* Driving the model to its limit: Profile likelihood based model reduction. *PLoS One* **11**, e0162366 (2016).
25. Wieland, F.-G., Hauber, A. L., Rosenblatt, M., Tönsing, C. & Timmer, J. On structural and practical identifiability. *Curr. Opin. Syst. Biol.* **25**, 60–69 (2021).
26. Sedaghat, A. R., Sherman, A. & Quon, M. J. A mathematical model of metabolic insulin signaling pathways. *Am. J. Physiol. Endocrinol. Metab.* **283**, E1084-101 (2002).
27. Van, G. J., Shun-Shion, A. S. & Fazakerley, D. J. Insulin signalling and GLUT4 trafficking in insulin resistance. *Biochemical Society Transactions* **51**, 1057–1069 (2023).
28. Titchenell, P. M. *et al.* Direct hepatocyte insulin signaling is required for lipogenesis but is dispensable for the suppression of glucose production. *Cell Metab.* **23**, 1154–1166 (2016).
29. Lewis, G. F., Carpentier, A. C., Pereira, S., Hahn, M. & Giacca, A. Direct and indirect control of hepatic glucose production by insulin. *Cell Metab.* **33**, 709–720 (2021).
30. Bergman, R. N., Ider, Y. Z., Bowden, C. R. & Cobelli, C. Quantitative estimation of insulin sensitivity. *Am. J. Physiol.* **236**, E667-77 (1979).
31. Schiavon, M. *et al.* Modeling plasma-to-interstitium glucose kinetics from multitracer plasma and microdialysis data. *Diabetes Technol. Ther.* **17**, 825–831 (2015).
32. Weinisch, P. *et al.* The HuMet Repository: Watching human metabolism at work. *bioRxiv* https://doi.org/10.1101/2023.08.08.550079 (2023) doi:10.1101/2023.08.08.550079.
33. Ha, J. *et al.* Estimating Insulin Sensitivity and Beta-Cell Function from the Oral Glucose Tolerance Test: Validation of a new Insulin Sensitivity and Secretion (ISS) Model. *Am. J. Physiol. Endocrinol. Metab.* https://doi.org/10.1152/ajpendo.00189.2023 (2023) doi:10.1152/ajpendo.00189.2023.
34. Dalla Man, C., Rizza, R. A. & Cobelli, C. Meal simulation model of the glucose-insulin system. *IEEE Trans. Biomed. Eng.* **54**, 1740–1749 (2007).
35. Fujii, M. *et al.* Logical design of oral glucose ingestion pattern minimizing blood glucose in humans. *NPJ Syst Biol Appl* **5**, 31 (2019).
36. Berry, S. E. *et al.* Human postprandial responses to food and potential for precision nutrition. *Nat. Med.* **26**, 964–973 (2020).
37. Manell, E., Hedenqvist, P., Svensson, A. & Jensen-Waern, M. Establishment of a refined oral glucose tolerance test in pigs, and assessment of insulin, glucagon and glucagon-like peptide-1 responses. *PLoS One* **11**, e0148896 (2016).

38. Panunzi, S., Pompa, M., Borri, A., Piemonte, V. & De Gaetano, A. A revised Sorensen model: Simulating glycemic and insulinemic response to oral and intra-venous glucose load. *PLoS One* **15**, e0237215 (2020).
39. Drevon, D., Fursa, S. R. & Malcolm, A. L. Intercoder reliability and validity of WebPlotDigitizer in extracting graphed data. *Behav. Modif.* **41**, 323–339 (2017).
40. Vicini, P., Caumo, A. & Cobelli, C. The hot IVGTT two-compartment minimal model: indexes of glucose effectiveness and insulin sensitivity. *Am. J. Physiol.* **273**, E1024-32 (1997).
41. Toffolo, G., Campioni, M., Basu, R., Rizza, R. A. & Cobelli, C. A minimal model of insulin secretion and kinetics to assess hepatic insulin extraction. *Am. J. Physiol. Endocrinol. Metab.* **290**, E169–E176 (2006).
42. Sugimoto, H. *et al.* Association between continuous glucose monitoring-derived metrics and coronary plaque vulnerability: A retrospective exploratory analysis. *eLife*, **14**:RP102860 (2026).
43. Mita, T. *et al.* Continuous glucose monitoring-derived time in range and CV are associated with altered tissue characteristics of the carotid artery wall in people with type 2 diabetes. *Diabetologia* **66**, 2356–2367 (2023).
44. Zhong, H. *et al.* Two-week continuous glucose monitoring-derived metrics and degree of hepatic steatosis: a cross-sectional study among Chinese middle-aged and elderly participants. *Cardiovasc. Diabetol.* **23**, 322 (2024).
45. Sugimoto, H. *et al.* Improved detection of decreased glucose handling capacities via continuous glucose monitoring-derived indices. *Commun. Med. (Lond.)* **5**, 103 (2025).
46. Sugimoto, H., Sapir, G., Keshet, A. & Kuroda, S. Use of continuous glucose monitoring to stratify individuals without diabetes. *Commun. Med. (Lond.)* **6** (2026).
